# Evaluating the Risk of Cardiovascular Adverse Events and Appendicitis After COVID-19 Diagnosis in Adults in the United States: Implications of the Start of Follow-Up

**DOI:** 10.1101/2024.09.05.24313134

**Authors:** J. Bradley Layton, Arnstein Lindaas, Stella G Muthuri, Patricia C. Lloyd, Morgan M. Richey, Joann F. Gruber, Hai Lyu, Mollie M. McKillop, Lisa S. Kowarski, Christine Bui, Shelby S. Fisher, Tainya C. Clarke, Angela S. Cheng, Zhiruo Wan, Pablo Freyria Duenas, Yangping Chen, Timothy Burrell, Minya Sheng, Azadeh Shoaibi, Yoganand Chillarige, Jeffrey Beers, Mary S. Anthony, Richard A. Forshee, Steven A. Anderson

## Abstract

**Purpose:** This study evaluated the association between coronavirus disease 2019 (COVID-19) diagnosis and adverse events (AEs), including cardiovascular AEs and appendicitis, in US adults before the introduction of COVID-19 vaccines. Real-world studies of AEs after COVID-19 suggest that diagnoses of AEs and COVID-19 frequently occur on the same day and may be a source of bias.

**Methods:** Cohort and self-controlled risk interval (SCRI) designs were used in 2 US administrative claims data sources—Merative™ MarketScan® (ages 18-64 years) and Medicare (ages ≥ 65 years). AEs included stroke (nonhemorrhagic and hemorrhagic), acute myocardial infarction, myocarditis/pericarditis, deep vein thrombosis, pulmonary embolism (PE), disseminated intravascular coagulation (DIC), unusual-site and common-site thrombosis with thrombocytopenia syndrome, and appendicitis. In cohort analyses, weighted hazard ratios (HRs) and 95% confidence intervals (CIs) compared adults with a COVID-19 diagnosis and matched comparators. In SCRI analysis, relative incidences (RIs) and 95% CIs compared risk and reference windows within individuals. Analyses were performed starting follow-up on Time 0 and Day 1.

**Results:** For cardiovascular AEs, all estimates starting follow-up on Day 1 were above 1.0 in both data sources. For cohort analyses, the strongest associations were for inpatient PE in both databases: MarketScan, HR=8.65 (95% CI, 6.06-12.35), Medicare HR=3.06 (95% CI, 2.88-3.26). For SCRI analyses, the strongest association in MarketScan was for DIC: RI=32.28 (95% CI, 17.06-61.09) and in Medicare was for myocarditis/pericarditis: RI=4.53 (95% CI, 3.89-5.27). AEs diagnosed concurrently with COVID-19 (ie, on Time 0) were common; including Time 0 in follow-up/risk windows resulted in higher RIs, as well as higher HRs for some AEs. However, some AEs (eg, stroke) were more common on Time 0 in the comparator group resulting in lower HRs.

**Conclusion:** COVID-19 diagnoses had moderate to strong associations with cardiovascular AEs and weak or inconsistent associations with appendicitis, although estimates varied by design and methodology.

## Introduction

Coronavirus disease 2019 (COVID-19) severity may range from mild to fatal and has been associated with a variety of sequelae. Previous studies from several countries have evaluated the association between COVID-19 and select adverse events (AEs) using cohort,^1–6^ case-control,^7^ and self-controlled^3,5,8–10^ study designs; these studies have generally reported increased risk of cardiovascular AEs, such as thromboembolism, pulmonary embolism, and acute myocardial infarction, and stroke. Limited evidence is available on the association of COVID-19 with appendicitis.^11,12^ Some real-world studies of the sequelae of COVID-19 have identified that AEs occurring on the same day as the COVID-19 diagnosis may have a substantial impact on effect measure estimates,^9,13^ but that effect may differ by study design, AE severity, and setting of COVID-19 diagnosis.

Understanding the risk of AEs after a COVID-19 diagnosis is important both for clinical care after COVID-19 and for better understanding the benefit-risk profile of COVID-19 vaccines, providing important context for vaccine safety surveillance findings. Additionally, understanding the implications of various study design choices on effect measure estimates may inform future research in this field. As part of its public health surveillance mandate, the US FDA Biologics Effectiveness and Safety (BEST) Initiative conducted this study to assess the association between COVID-19 and select AEs—including nonhemorrhagic stroke (NHS), hemorrhagic stroke (HS), acute myocardial infarction (AMI), myocarditis/pericarditis, deep vein thrombosis (DVT), pulmonary embolism(PE), disseminated intravascular coagulation (DIC), immune thrombocytopenia (ITP), common-site and unusual-site thrombosis with thrombocytopenia syndrome (TTS), and appendicitis—before the introduction of COVID-19 vaccines in US adults. Neurologic/immune-mediated AEs were also evaluated but are presented elsewhere. To evaluate the potential impact of various biases (eg, exposure misclassification, confounding, reverse causality, and selection bias), two study designs (ie, a cohort and a self-controlled risk interval [SCRI]) were implemented with varying specifications (eg, start of follow-up/risk windows).

## Material and Methods

### Data Sources

This study used 2 administrative health insurance claims databases participating in the FDA CBER BEST Initiative: Merative™ MarketScan® Commercial Database and US Centers for Medicare and Medicaid Services (CMS) Medicare fee-for-service. All analyses were performed separately in each data source according to a common protocol.^14^ The analyses were restricted to individuals aged 18 to 64 years in MarketScan and those aged 65 years or older in Medicare. Individuals in Medicare were required to have coverage with Part A (inpatient hospital care) and Part B (outpatient care and physician services).

### Study Designs

Two different study designs were used—an SCRI and a cohort—to evaluate each AE individually. Using data from the COVID-19 pandemic presented certain challenges, such as temporary disruptions in healthcare utilization,^15–17^ which may have different impacts on the 2 study designs. The cohort design used a matched comparator group, but because not all COVID-19 cases resulted in a medical diagnosis, exposure misclassification can be expected, and individuals with COVID-19 undiagnosed in clinical settings may be present in the comparator group, potentially resulting in estimates biased toward the null. Additionally, confounding between exposure groups may be difficult to account for if the COVID-19-diagnosed group differed from the comparator in ways that were difficult to measure (eg, adherence to preventive measures, risk tolerance, behavioral or lifestyle factors). While misclassification of the COVID-19 diagnosis and residual confounding by time-invariant factors are less of a concern in the SCRI design, the SCRI requires key assumptions that may be violated (eg, AE event rates remain constant over time; a biologically relevant risk window for an AE that can be clearly defined relative to the COVID-19 diagnosis; and that events do not influence the end of the observation period). Thus, both study designs were used, and their results are presented as complementary. In both SCRI and cohort analyses, AE-specific exclusion criteria were applied, resulting in a unique analytic data set for each AE.

#### SCRI

The SCRI design^18^ included only those diagnosed with COVID-19 who also experienced an AE of interest during the study period, and compared an individual’s risk of AEs during a risk window immediately after the COVID-19 diagnosis to reference windows before and after the diagnosis (Figure 1, Table 1).^18,19^ Individuals were eligible for the SCRI if they had a COVID-19 diagnosis between 1 June 2020 and 10 December 2020—the day before the first COVID-19 vaccine was authorized for use in the US. This period removed the potential for vaccine-associated AEs being counted as COVID-19-associated AEs. The SCRI study period began in June 2020, as pandemic-related reductions in diagnoses for a variety of conditions^15–17^ may violate underlying assumptions of the SCRI analysis that outcome rates are consistent over the time period^18^; by June 2020, many measures of healthcare utilization returned to pre-pandemic levels in healthcare claims.^15,17^

**Figure 1.**
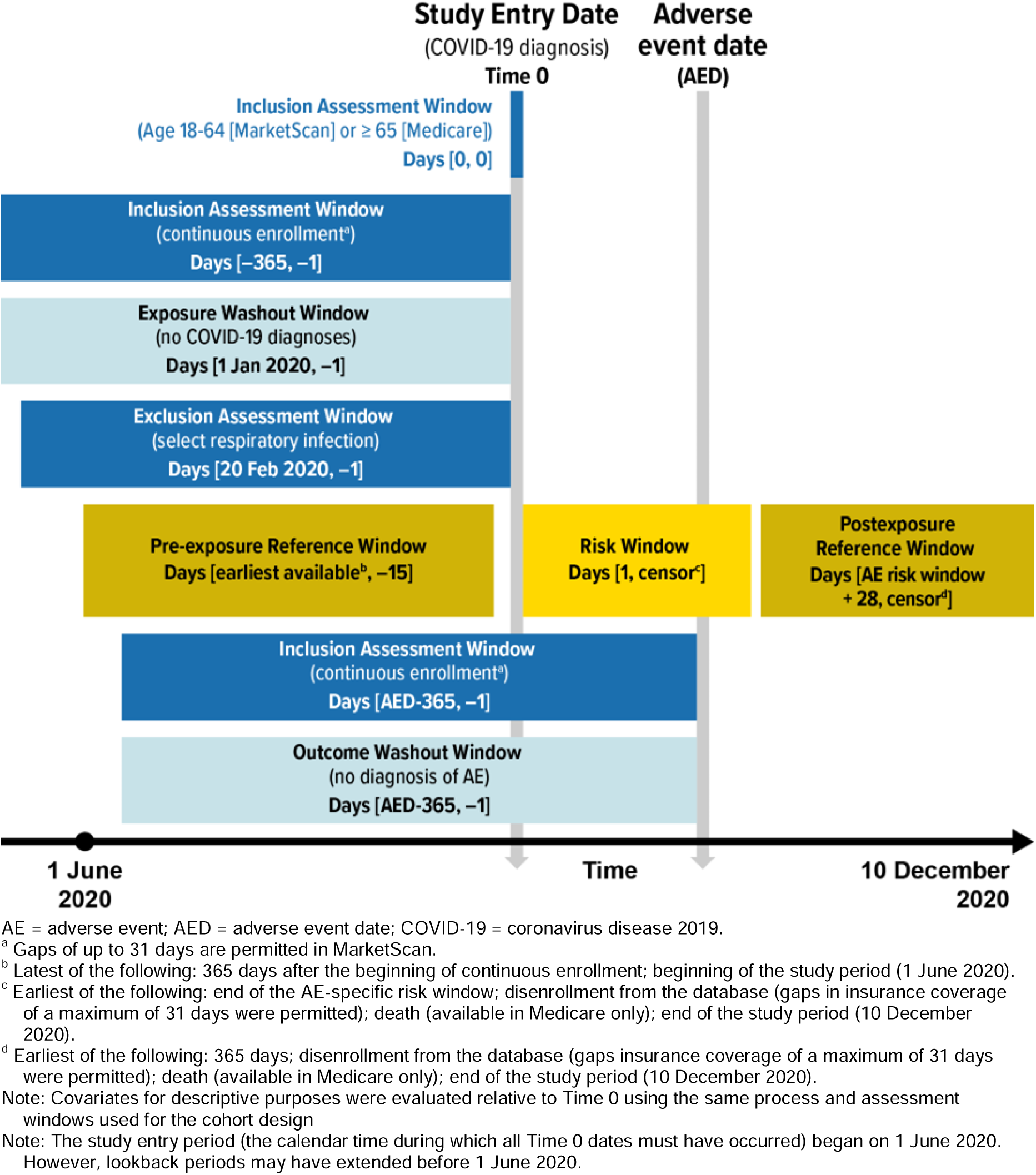
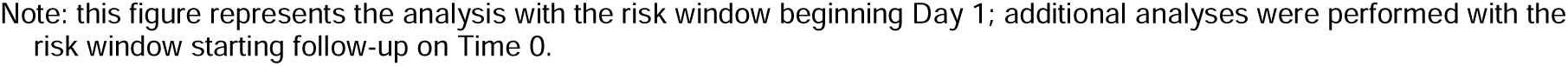
Eligibility Assessment, Covariate Assessment, Risk Windows, and Reference Windows Relative to Time 0 for the Self-Controlled Risk Interval Design.

**Table 1.** Analysis Set and Definition for Each Specified Adverse Event.

| Adverse event | Care setting | Risk window for SCRI<sup>a</sup> | Washout window (days)<sup>b</sup> |
| --- | --- | --- | --- |
| Nonhemorrhagic stroke | IP | NA | 365 |
| Hemorrhagic stroke | IP | NA | 365 |
| Acute myocardial infarction | IP | NA | 365 |
| Myocarditis/pericarditis | IP, OP/PB | Days 1-41 | 365 |
| Deep vein thrombosis | IP, OP/PB | Days 1-27 | 365 |
| Pulmonary embolism – any setting<sup>c</sup> | IP, OP/PB | Days 1-27 | 365 |
| Pulmonary embolism – inpatient only<sup>c</sup> | IP | Days 1-27 | 365 |
| Disseminated intravascular coagulation | IP, OP/PB | Days 1-27 | 365 |
| Unusual-site thrombosis with thrombocytopenia syndrome<br>▪ Unusual-site thrombosis (intracranial or intra-abdominal venous thrombosis, such as in portal, renal, and other veins) AND<br>▪ Thrombocytopenia within ≤ 14 days | Thrombosis: IP, ED<br>Thrombocytopenia: IP, OP/PB | NA | 365 |
| Common-site thrombosis with thrombocytopenia syndrome<br>▪ Common-site thrombosis (AMI, HS, NHS, DVT, PE) AND<br>▪ Thrombocytopenia within ≤ 14 days | AMI, HS, and NHS: IP<br>DVT, PE, and thrombocytopenia: IP, OP/PB | NA | 365 |
| Appendicitis | IP, OP-ED | Days 1-41 | 365 |

AMI = acute myocardial infarction; DVT = deep vein thrombosis; ED = emergency department; HS = hemorrhagic stroke; IP = inpatient; NA = not applicable; NHS = nonhemorrhagic stroke; OP/PB = outpatient or provider claims; PE = pulmonary embolism; SCRI = self-controlled risk interval.
<sup>a</sup> For the SCRI analysis, the risk window for the main analysis started on Day 1; an additional analysis included Time 0 (ie, the date of the COVID-19 diagnosis) in the risk window.
<sup>b</sup> For the cohort analysis, the washout windows is applied to the specified period before and including the date of COVID-19 diagnosis or matched date (ie, Time 0). For the SCRI, the washout window is applied to the specified period before and not including the date of the AE occurring in the risk or reference window.
<sup>c</sup> Evaluated separately in inpatient settings only, and in combined inpatient, outpatient, or provider claims settings; both outcomes were evaluated in the same analysis set.
Note: Details of outcome algorithms given in the study protocol (CBER Surveillance Program <sup>14</sup>).

Individuals included in the SCRI analyses were followed until disenrollment from the database, death (in Medicare only), or the end of the study period (Figure 2). The length of the risk window was defined separately for each AE (Figure 1). Pre-exposure and post-exposure reference windows were defined using data up to 365 days before and after the risk window (Figure 1); as much data as were available for an individual during the study period were included. Individuals were required to have at least 1 day in the pre-exposure reference window for inclusion. Buffer periods immediately before (14 days) and after (28 days) the risk window were excluded from both risk and reference windows.

**Figure 2.**
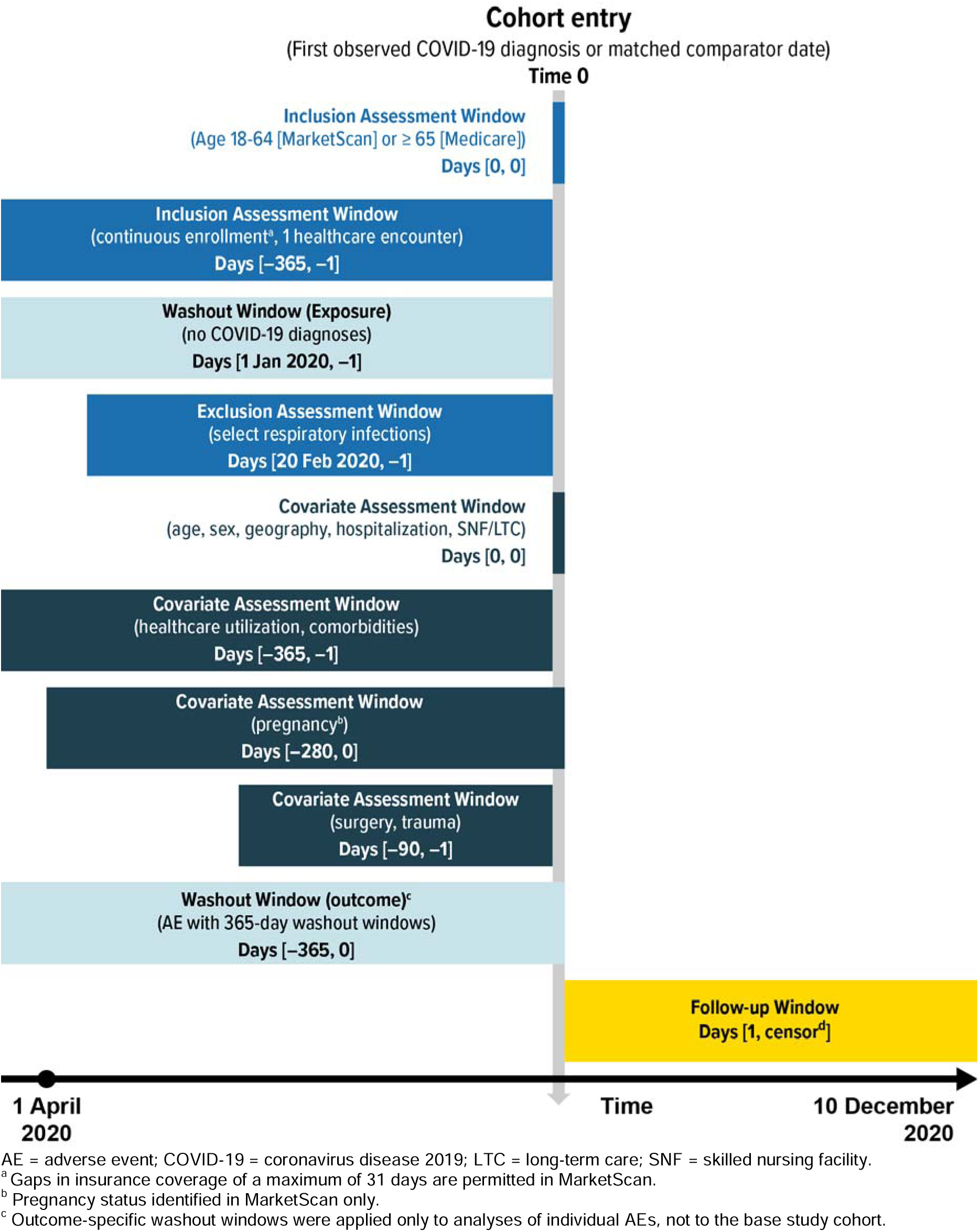
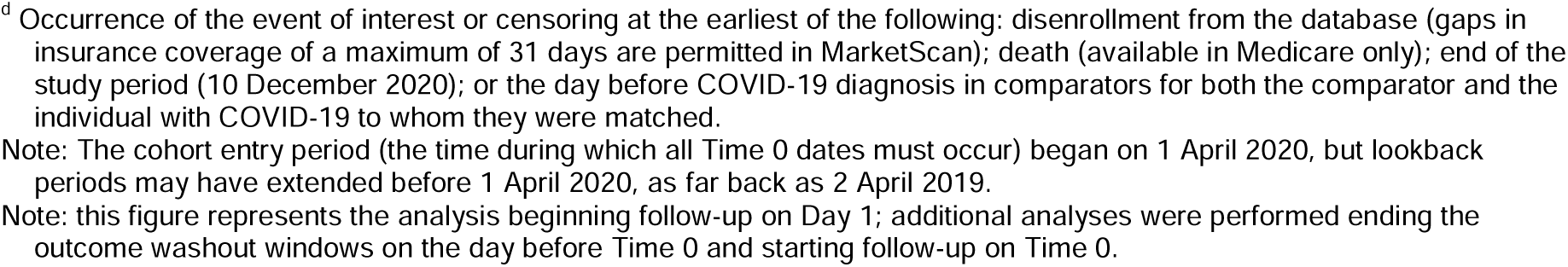
Eligibility Assessment, Covariate Assessment, and Follow-Up Windows Relative to Time 0 for the Cohort Design.

#### Cohort Design

The cohort design identified individuals with a recorded COVID-19 diagnosis and matched comparators who were not diagnosed with COVID-19 as of the matched calendar date (ie, the diagnosis date of the COVID-19 case). The study period for the cohort spanned the period between 1 April 2020—the date the *International Classification of Diseases, Tenth Revision, Clinical Modification* (ICD-10-CM) code for COVID-19 (U07.1) was introduced—to 10 December 2020.

Individuals were identified at their first COVID-19 diagnosis during the study period; the diagnosis date was used as Time 0 in the study. The calendar date of matching became Time 0 for the comparator group. Individuals in the COVID-19-diagnosed group were 1:1 exact-matched with replacement to comparators on age (5-year increments), sex, geographic region, immunocompromised status, hospitalization status on Time 0, and skilled nursing facility/long-term care residence on Time 0 (in Medicare). Hospitalization status on Time 0 was included as a matching variable as a proxy for COVID-19 disease severity, ensuring that a COVID-19 patient admitted to the hospital was matched to a comparator who was also admitted to the hospital on that same day (in Medicare) or who was already in the hospital (in MarketScan). Matching on hospitalization was also intended to address potential surveillance bias, present if the COVID-19-diagnosed group was more likely to have healthcare interactions than the comparator group, as typical patterns of healthcare utilization and diagnosis were temporarily altered during the early phase of the pandemic.^16,20^ Comparators who were matched to a COVID-19 case remained eligible to enter the COVID-19-diagnosed group or act as a comparator to an additional COVID-19 case as long as they remained eligible at a given calendar date. Individuals were followed until the end of the study period, disenrollment from the database, death (in Medicare only), the day before COVID-19 diagnosis in the matched comparator, or the occurrence of an outcome (Figure 2).

### Variables

Recorded diagnoses were identified with ICD-10-CM diagnosis codes; additionally, Diagnosis Related Groups (DRGs) were also used to identify medical conditions. Pharmacy dispensing of influenza vaccines was identified with National Drug Codes in MarketScan only. Procedures were identified with Current Procedural Terminology (CPT), Healthcare Common Procedure Coding System (HCPCS), or *ICD-10 Procedure Coding System* (ICD-10-PCS) or DRG codes.

#### Exposure

In both study designs, the primary exposure of interest was a COVID-19 diagnosis (ICD-10-CM code U07.1) identified in claims data from inpatient, emergency department (ED), outpatient, or professional service claims in any coding position (in Medicare, admitting diagnosis codes were not considered). The diagnosis date was defined as the index date for the exposure group in each data source (eg, the date of admission for facility-level diagnoses, the service date for professional service-level diagnoses).

#### Outcomes

The outcomes of interest for this study included AEs that have previously been included in safety surveillance studies of COVID-19 vaccines, identified using ICD-10-CM diagnosis codes. AE-specific washout windows were employed in both study designs to restrict outcomes to new-onset AEs (Table 1). Washout windows for the cohort study were applied to the 12-month period before the date of COVID-19 diagnosis or the matched index date for comparators (Figure 1). The washout windows for the SCRI analysis were applied to a 12-month period before the date of the AE (Figure 2).

#### Covariates

Covariates were measured from claims, enrollment, and assessment information in each data source and included demographic and clinical characteristics, comorbidities, and healthcare utilization. Covariates were used to describe the characteristics of each study sample in both study designs and to estimate propensity scores in the cohort analysis (details of AE-specific covariates are given in the supplemental information, sTable 1).

#### Follow-Up

For both study designs, analyses were performed both starting the follow-up/risk period on Time 0 (ie, the day of COVID-19 diagnosis or matched comparator date) and starting the follow-up/risk period on Day 1 (ie, the day after Time 0). Additionally, to evaluate the potential impact of AE cases on Time 0, risk estimates on Time 0 alone were evaluated.

### SCRI Statistical Methods

The baseline descriptive characteristics of the included individuals with COVID-19 were described with means and standard deviations (SD) and medians and first and third quartiles (Q1, Q3) for continuous variables, and counts and percentages for categorical variables. Within each outcome-specific analysis set, the relative incidence (RI) and 95% confidence interval (CI)^19,21^ for the association between the AE and a COVID-19 diagnosis was estimated using conditional Poisson regression accounting for the variable length of follow-up in the risk and reference windows.^13^ Because some outcomes may increase the risk of mortality, the assumption of having no event-dependent censoring may be violated^13^ and an extended Poisson model accounting for event-dependent observation windows was used^22^ to estimate RI and 95% CI. AEs with particularly high case fatality rates (approximately 10% or greater, such as acute myocardial infarction, nonhemorrhagic and hemorrhagic stroke, and unusual-site and common-site TTS) were not evaluated with the SCRI design. Some AEs (i.e., pulmonary embolism, disseminated intravascular coagulation) were evaluated with the SCRI in MarketScan but not in Medicare due to higher case fatality rates in the older Medicare population compared to the younger commercially insured population in MarketScan. Additionally, models were further adjusted for calendar month to account for potential seasonality and time trends. To contextualize the RI observed in the SCRI analysis, attributable risk (ie, the number of excess cases of the AE observed because of COVID-19 diagnosis) was estimated; a formula adapted from Yih and colleagues^23^ and Ammann and colleagues￼ estimated the AR from the RI, the number of AE cases in the risk window, and the total number of eligible COVID-19 cases.

### Cohort Study Statistical Methods

Patient characteristics were described in the overall matched cohort before application of the washout period (Table 1). The relative balance of characteristics between the COVID-19-diagnosed and matched comparator groups were described with absolute standardized differences (ASDs).^25^ For each AE, after outcome-specific exclusion requirements were applied, an AE-specific propensity score model was estimated using a priori identified covariates, including potential confounders for each AE.^26,27^ The full list of covariates that were selected for inclusion in outcome-specific propensity score models, as appropriate for each outcome, are detailed in the supplemental information (sTable 1). The distributions of propensity scores were evaluated by exposure groups. The propensity scores were utilized to estimate stabilized inverse probability of treatment (sIPT) weights for each AE analysis. sIPT weights were truncated below the 1st and above the 99th percentiles to minimize the impact of extreme weights. Covariate balance between exposure groups was evaluated for both the crude and weighted cohorts using ASDs.^25^

For the sIPT-weighted cohorts, Kaplan-Meier estimates of survival were generated,^28^ and robust variance estimators were utilized to estimate 95% CIs to account for re-use of comparators.^29^ The daily cumulative incidence estimates were calculated for each follow-up day by subtracting the Kaplan-Meier survival estimate and 95% CI from 1.0. Cox proportional hazard models were utilized to estimate crude and sIPT-weighted hazard ratios (HRs), and 95% CIs were estimated using robust sandwich variance estimators. Subgroup analyses were performed by hospitalization status at Time 0.

Analyses were conducted with SAS version 9.4 (SAS Institute Inc., Cary, NC, USA) and R versions 4.1.2 and 4.1.3 (R Core Team 2021). This surveillance activity was conducted as part of the FDA public health surveillance mandate and was not subject to Institutional Review Board oversight.

## Results

### SCRI

For the SCRI analyses, we identified 330,799 eligible individuals with a COVID-19 diagnosis in MarketScan (mean [SD] age, 41.4 [13.7] years, 54.7% female), and 855,065 in Medicare (mean [SD] age, 77.4 [8.4] years, 56.9% female) (Table 2, sTable 2).

**Table 2.**
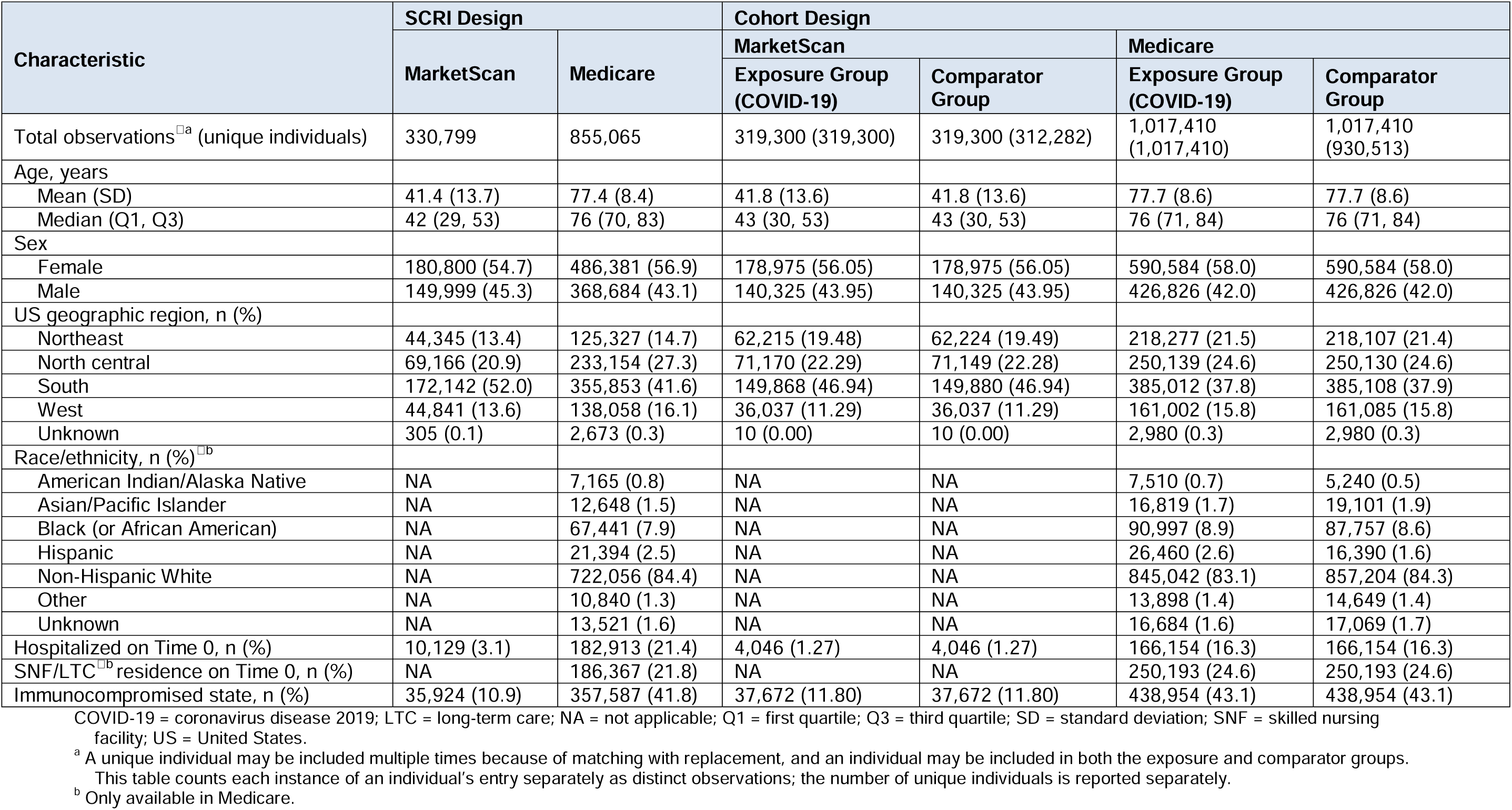
Selected Characteristics of Included Individuals for SCRI and Cohort Analyses.

| Characteristic | SCRI Design |  | Cohort Design |  |  |  |
| --- | --- | --- | --- | --- | --- | --- |
|  | MarketScan | Medicare | MarketScan |  | Medicare |  |
|  |  |  | Exposure Group (COVID-19) | Comparator Group | Exposure Group (COVID-19) | Comparator Group |
| Total observations<sup>□a</sup> (unique individuals) | 330,799 | 855,065 | 319,300 (319,300) | 319,300 (312,282) | 1,017,410 (1,017,410) | 1,017,410 (930,513) |
| Age, years |  |  |  |  |  |  |
| Mean (SD) | 41.4 (13.7) | 77.4 (8.4) | 41.8 (13.6) | 41.8 (13.6) | 77.7 (8.6) | 77.7 (8.6) |
| Median (Q1, Q3) | 42 (29, 53) | 76 (70, 83) | 43 (30, 53) | 43 (30, 53) | 76 (71, 84) | 76 (71, 84) |
| Sex |  |  |  |  |  |  |
| Female | 180,800 (54.7) | 486,381 (56.9) | 178,975 (56.05) | 178,975 (56.05) | 590,584 (58.0) | 590,584 (58.0) |
| Male | 149,999 (45.3) | 368,684 (43.1) | 140,325 (43.95) | 140,325 (43.95) | 426,826 (42.0) | 426,826 (42.0) |
| US geographic region, n (%) |  |  |  |  |  |  |
| Northeast | 44,345 (13.4) | 125,327 (14.7) | 62,215 (19.48) | 62,224 (19.49) | 218,277 (21.5) | 218,107 (21.4) |
| North central | 69,166 (20.9) | 233,154 (27.3) | 71,170 (22.29) | 71,149 (22.28) | 250,139 (24.6) | 250,130 (24.6) |
| South | 172,142 (52.0) | 355,853 (41.6) | 149,868 (46.94) | 149,880 (46.94) | 385,012 (37.8) | 385,108 (37.9) |
| West | 44,841 (13.6) | 138,058 (16.1) | 36,037 (11.29) | 36,037 (11.29) | 161,002 (15.8) | 161,085 (15.8) |
| Unknown | 305 (0.1) | 2,673 (0.3) | 10 (0.00) | 10 (0.00) | 2,980 (0.3) | 2,980 (0.3) |
| Race/ethnicity, n (%)<sup>□b</sup> |  |  |  |  |  |  |
| American Indian/Alaska Native | NA | 7,165 (0.8) | NA | NA | 7,510 (0.7) | 5,240 (0.5) |
| Asian/Pacific Islander | NA | 12,648 (1.5) | NA | NA | 16,819 (1.7) | 19,101 (1.9) |
| Black (or African American) | NA | 67,441 (7.9) | NA | NA | 90,997 (8.9) | 87,757 (8.6) |
| Hispanic | NA | 21,394 (2.5) | NA | NA | 26,460 (2.6) | 16,390 (1.6) |
| Non-Hispanic White | NA | 722,056 (84.4) | NA | NA | 845,042 (83.1) | 857,204 (84.3) |
| Other | NA | 10,840 (1.3) | NA | NA | 13,898 (1.4) | 14,649 (1.4) |
| Unknown | NA | 13,521 (1.6) | NA | NA | 16,684 (1.6) | 17,069 (1.7) |
| Hospitalized on Time 0, n (%) | 10,129 (3.1) | 182,913 (21.4) | 4,046 (1.27) | 4,046 (1.27) | 166,154 (16.3) | 166,154 (16.3) |
| SNF/LTC<sup>□b</sup> residence on Time 0, n (%) | NA | 186,367 (21.8) | NA | NA | 250,193 (24.6) | 250,193 (24.6) |
| Immunocompromised state, n (%) | 35,924 (10.9) | 357,587 (41.8) | 37,672 (11.80) | 37,672 (11.80) | 438,954 (43.1) | 438,954 (43.1) |

COVID-19 = coronavirus disease 2019; LTC = long-term care; NA = not applicable; Q1 = first quartile; Q3 = third quartile; SD = standard deviation; SNF = skilled nursing facility; US = United States.
<sup>a</sup> A unique individual may be included multiple times because of matching with replacement, and an individual may be included in both the exposure and comparator groups.
This table counts each instance of an individual's entry separately as distinct observations; the number of unique individuals is reported separately.
<sup>b</sup> Only available in Medicare.

In the analyses starting the risk window on Day 1, elevated RI estimates (ie, RI > 3) were observed across both data sources for myocarditis/pericarditis and DVT (Table 3). Pulmonary embolism and DIC were only evaluated with the SCRI in MarketScan, and the resulting RI estimates were very high; the largest RI estimate observed was for DIC in MarketScan (RI = 32.28; 95% CI, 17.06-61.09). RI estimates for appendicitis were generally close to null (MarketScan RI = 1.38; 95% CI, 1.00-1.90; Medicare RI = 0.98; 95% CI, 0.70-1.36). The attributable risk estimates suggested large absolute numbers of AE cases associated with COVID-19 diagnosis (Table 3), with the largest estimate in MarketScan being for pulmonary embolism (177.23 cases per 100,000 COVID-19 cases) and for DVT in Medicare (437.75 cases per 100,000 COVID-19 cases).

**Table 3.**
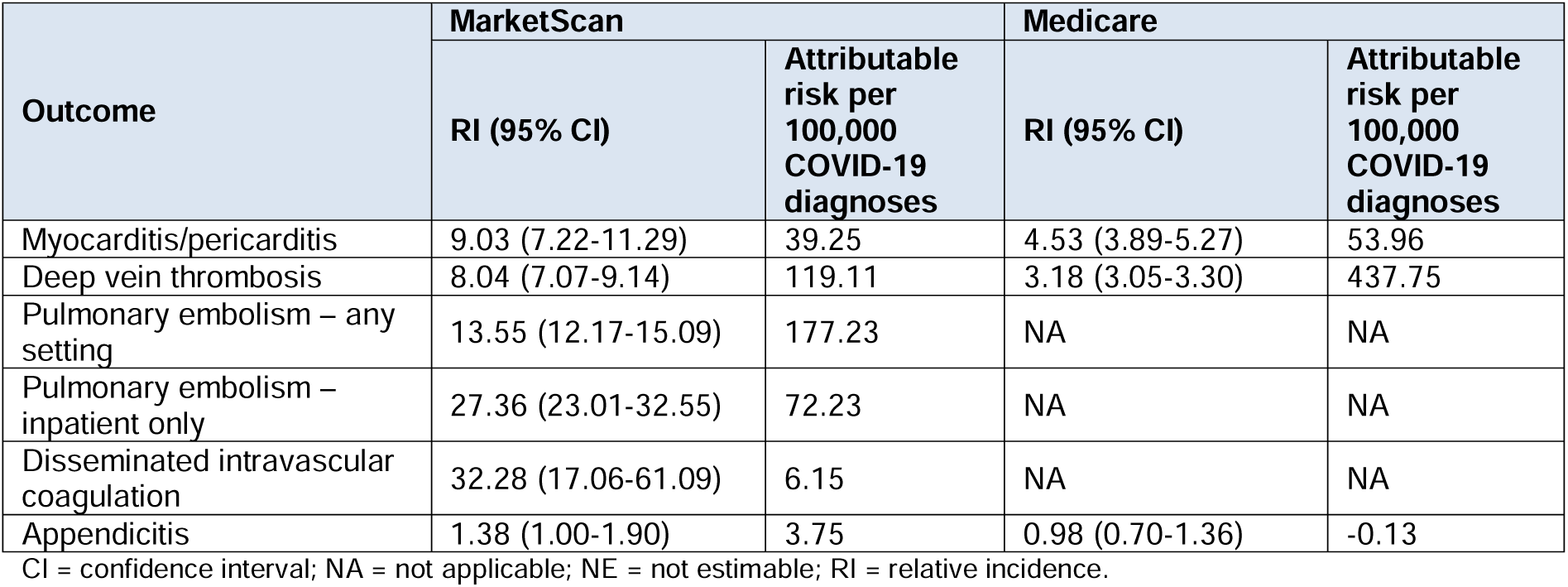
Estimated Association of a COVID-19 Diagnosis With Cardiovascular Adverse Events and Appendicitis, Self-Controlled Risk Interval Analysis, Beginning Follow-Up on Day 1.

| Outcome | MarketScan |  | Medicare |  |
| --- | --- | --- | --- | --- |
|  | RI (95% CI) | Attributable risk per 100,000 COVID-19 diagnoses | RI (95% CI) | Attributable risk per 100,000 COVID-19 diagnoses |
| Myocarditis/pericarditis | 9.03 (7.22-11.29) | 39.25 | 4.53 (3.89-5.27) | 53.96 |
| Deep vein thrombosis | 8.04 (7.07-9.14) | 119.11 | 3.18 (3.05-3.30) | 437.75 |
| Pulmonary embolism – any setting | 13.55 (12.17-15.09) | 177.23 | NA | NA |
| Pulmonary embolism – inpatient only | 27.36 (23.01-32.55) | 72.23 | NA | NA |
| Disseminated intravascular coagulation | 32.28 (17.06-61.09) | 6.15 | NA | NA |
| Appendicitis | 1.38 (1.00-1.90) | 3.75 | 0.98 (0.70-1.36) | -0.13 |

CI = confidence interval; NA = not applicable; NE = not estimable; RI = relative incidence.

In both data sources, a substantial number of AE cases were observed on Time 0 (the day of COVID-19 diagnosis) (sTable 3). RI estimates when Time 0 was included in the risk window were higher than those not including Time 0 in the risk window for all AEs in both data sources (sTable 3), and when the risk window consisted of only Time 0, RI estimates were extremely high (ie, RI > 40) for all outcomes.

### Cohort

In the final, matched cohorts in MarketScan, we identified 358,306 eligible individuals aged 18 through 65 years with a COVID-19 diagnosis. After matching, 319,300 (89.11%) eligible adults in the COVID-19-diagnosed group could be matched to an individual without a recorded COVID-19 diagnosis on or before the index date (sTable 4). In Medicare, we identified 1,085,418 eligible adults aged 65 years or older with new-onset COVID-19. Of these eligible adults, 1,017,410 (93.7%) could be matched to a comparator without a prior COVID-19 diagnosis (sTable 4). Among the 8,738 patients with a COVID-19 diagnosis in MarketScan who could not be matched, 22.4% were hospitalized at Time 0; whereas in Medicare, among the 68,008 patients with a COVID-19 diagnosis who could not be matched, 99.6% were hospitalized at Time 0 (sTable 5).

In the MarketScan cohort, the mean age of both the COVID-19-diagnosed group and comparator group was 41.8 years (SD 13.6), with 56% of the cohort being female. In Medicare, the mean age in both groups was 77.7 years (SD 8.6 years), and 58% were female. In both data sources, the characteristics of the overall COVID-19-diagnosed group and the comparator group were largely similar and well balanced (sTable 6). Within each outcome-specific analysis set and after sIPT weighting, all characteristics remained well balanced (sFigure 1).

The HRs for the analyses starting follow-up on Day 1 demonstrated generally elevated weighted HRs for all outcomes across both data sources (Table 4), with the largest HRs observed for inpatient pulmonary embolism (MarketScan HR = 8.65, 95% CI, 6.06-12.35; Medicare HR = 3.06, 95% CI, 2.88-3.26). The weakest associations were observed for appendicitis (MarketScan HR = 1.14, 95% CI, 0.86-1.50; Medicare HR = 1.22, 95% CI, 0.97-1.53).

**Table 4.**
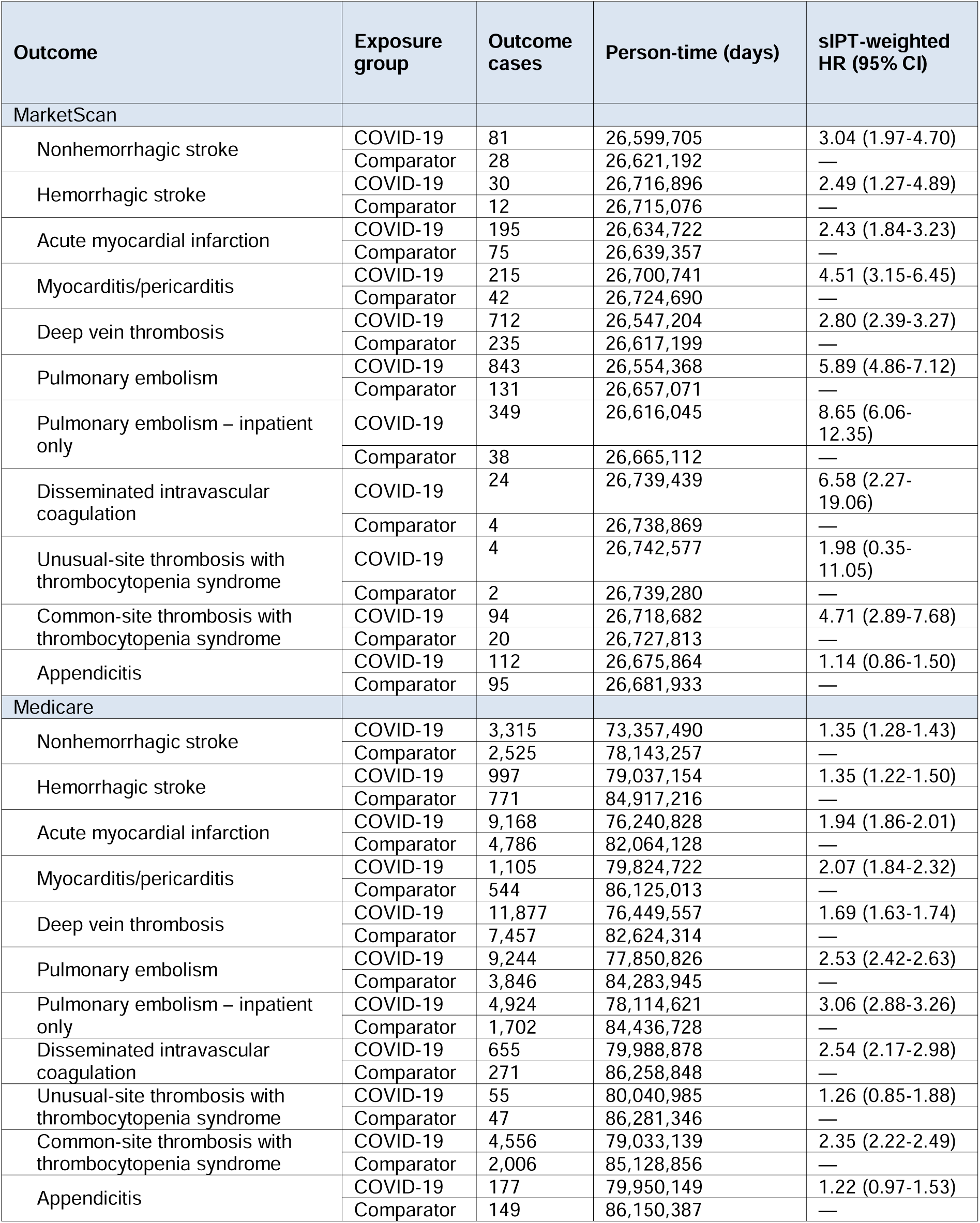
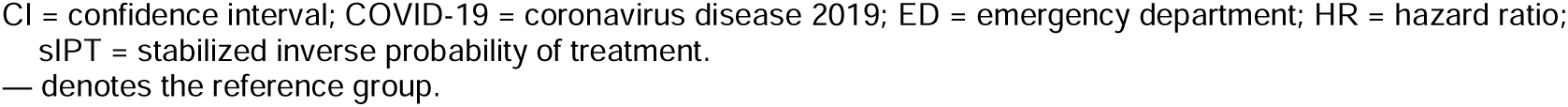
Estimated Association of a COVID-19 Diagnosis With Cardiovascular Adverse Events and Appendicitis, Cohort Analysis, Beginning Follow-Up on Day 1.

| Outcome | Exposure group | Outcome cases | Person-time (days) | sIPT-weighted HR (95% CI) |
| --- | --- | --- | --- | --- |
| **MarketScan** |  |  |  |  |
| Nonhemorrhagic stroke | COVID-19 | 81 | 26,599,705 | 3.04 (1.97-4.70) |
|  | Comparator | 28 | 26,621,192 | — |
| Hemorrhagic stroke | COVID-19 | 30 | 26,716,896 | 2.49 (1.27-4.89) |
|  | Comparator | 12 | 26,715,076 | — |
| Acute myocardial infarction | COVID-19 | 195 | 26,634,722 | 2.43 (1.84-3.23) |
|  | Comparator | 75 | 26,639,357 | — |
| Myocarditis/pericarditis | COVID-19 | 215 | 26,700,741 | 4.51 (3.15-6.45) |
|  | Comparator | 42 | 26,724,690 | — |
| Deep vein thrombosis | COVID-19 | 712 | 26,547,204 | 2.80 (2.39-3.27) |
|  | Comparator | 235 | 26,617,199 | — |
| Pulmonary embolism | COVID-19 | 843 | 26,554,368 | 5.89 (4.86-7.12) |
|  | Comparator | 131 | 26,657,071 | — |
| Pulmonary embolism – inpatient only | COVID-19 | 349 | 26,616,045 | 8.65 (6.06-12.35) |
|  | Comparator | 38 | 26,665,112 | — |
| Disseminated intravascular coagulation | COVID-19 | 24 | 26,739,439 | 6.58 (2.27-19.06) |
|  | Comparator | 4 | 26,738,869 | — |
| Unusual-site thrombosis with thrombocytopenia syndrome | COVID-19 | 4 | 26,742,577 | 1.98 (0.35-11.05) |
|  | Comparator | 2 | 26,739,280 | — |
| Common-site thrombosis with thrombocytopenia syndrome | COVID-19 | 94 | 26,718,682 | 4.71 (2.89-7.68) |
|  | Comparator | 20 | 26,727,813 | — |
| Appendicitis | COVID-19 | 112 | 26,675,864 | 1.14 (0.86-1.50) |
|  | Comparator | 95 | 26,681,933 | — |
| **Medicare** |  |  |  |  |
| Nonhemorrhagic stroke | COVID-19 | 3,315 | 73,357,490 | 1.35 (1.28-1.43) |
|  | Comparator | 2,525 | 78,143,257 | — |
| Hemorrhagic stroke | COVID-19 | 997 | 79,037,154 | 1.35 (1.22-1.50) |
|  | Comparator | 771 | 84,917,216 | — |
| Acute myocardial infarction | COVID-19 | 9,168 | 76,240,828 | 1.94 (1.86-2.01) |
|  | Comparator | 4,786 | 82,064,128 | — |
| Myocarditis/pericarditis | COVID-19 | 1,105 | 79,824,722 | 2.07 (1.84-2.32) |
|  | Comparator | 544 | 86,125,013 | — |
| Deep vein thrombosis | COVID-19 | 11,877 | 76,449,557 | 1.69 (1.63-1.74) |
|  | Comparator | 7,457 | 82,624,314 | — |
| Pulmonary embolism | COVID-19 | 9,244 | 77,850,826 | 2.53 (2.42-2.63) |
|  | Comparator | 3,846 | 84,283,945 | — |
| Pulmonary embolism – inpatient only | COVID-19 | 4,924 | 78,114,621 | 3.06 (2.88-3.26) |
|  | Comparator | 1,702 | 84,436,728 | — |
| Disseminated intravascular coagulation | COVID-19 | 655 | 79,988,878 | 2.54 (2.17-2.98) |
|  | Comparator | 271 | 86,258,848 | — |
| Unusual-site thrombosis with thrombocytopenia syndrome | COVID-19 | 55 | 80,040,985 | 1.26 (0.85-1.88) |
|  | Comparator | 47 | 86,281,346 | — |
| Common-site thrombosis with thrombocytopenia syndrome | COVID-19 | 4,556 | 79,033,139 | 2.35 (2.22-2.49) |
|  | Comparator | 2,006 | 85,128,856 | — |
| Appendicitis | COVID-19 | 177 | 79,950,149 | 1.22 (0.97-1.53) |
|  | Comparator | 149 | 86,150,387 | — |

CI = confidence interval; COVID-19 = coronavirus disease 2019; ED = emergency department; HR = hazard ratio; SIPT = stabilized inverse probability of treatment. — denotes the reference group.

Many AE cases were observed on Time 0, though the distribution of Time 0 cases across exposure groups differed by AE. For some severe AEs (NHS, HS, AMI, and unusual-site TTS in Medicare), AEs occurring on Time 0 were more common in the comparator group (Figure 3, sTable 7). For myocarditis/pericarditis, AEs on Time 0 were much more common in the COVID-19-diagnosed group (Figure 4, sTable 7). In the analyses including Time 0 in follow-up in both data sources, HR estimates for NHS, HS, and AMI were substantially lower than the analyses starting on Day 1; in Medicare, paradoxical HRs below 1 were observed for HS and NHS (sTable 7). However, for myocarditis/pericarditis, analyses including Time 0 in follow-up resulted in much higher HR estimates than those starting follow-up on Day 1, particularly in MarketScan (sTable 7). In both instances, plots of the cumulative incidence by exposure group starting on Time 0 suggested that the largest difference between exposure groups occurred on Time 0, though the direction of the difference differed by AE (Figure 3, Figure 4).

**Figure 3.**
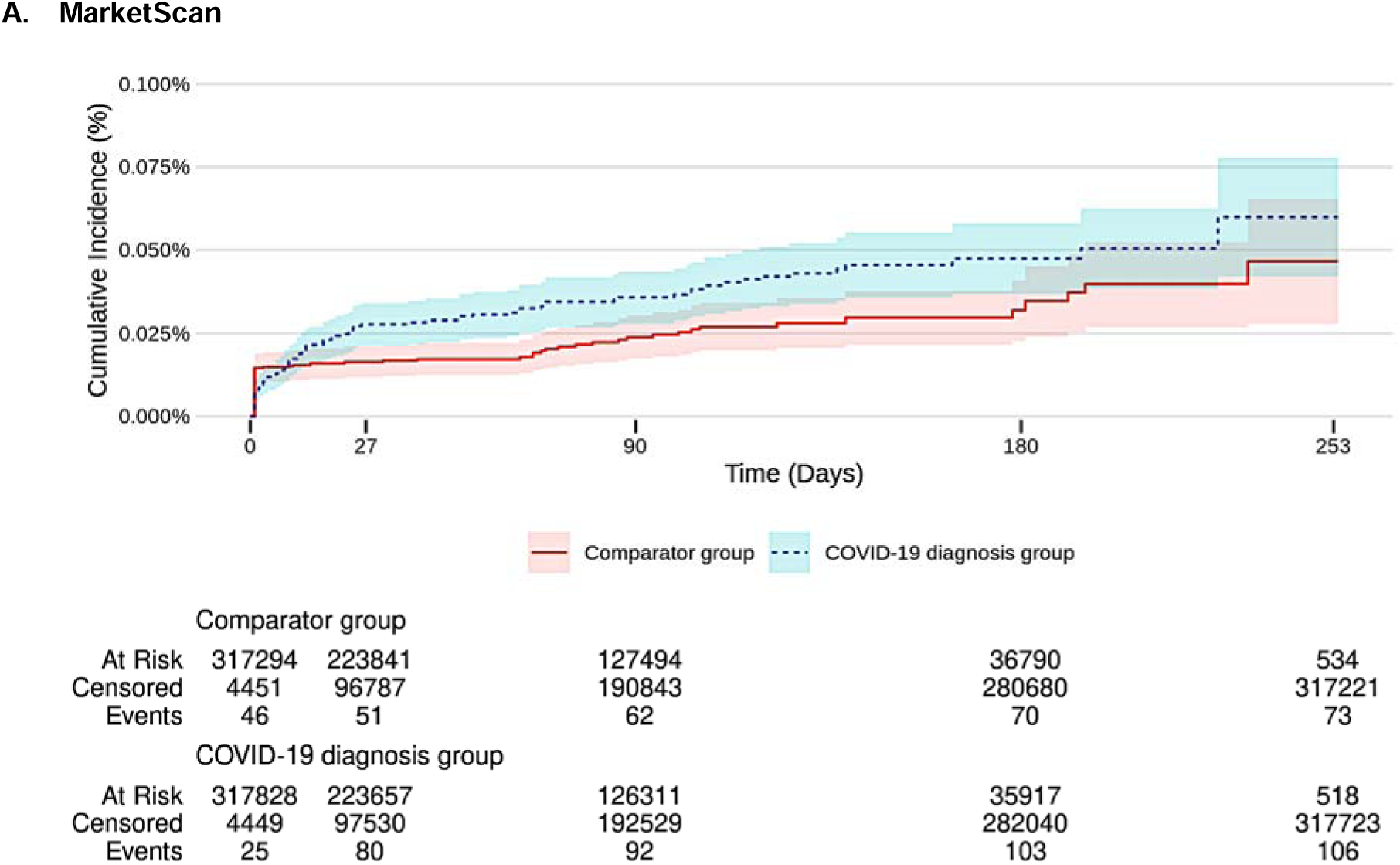
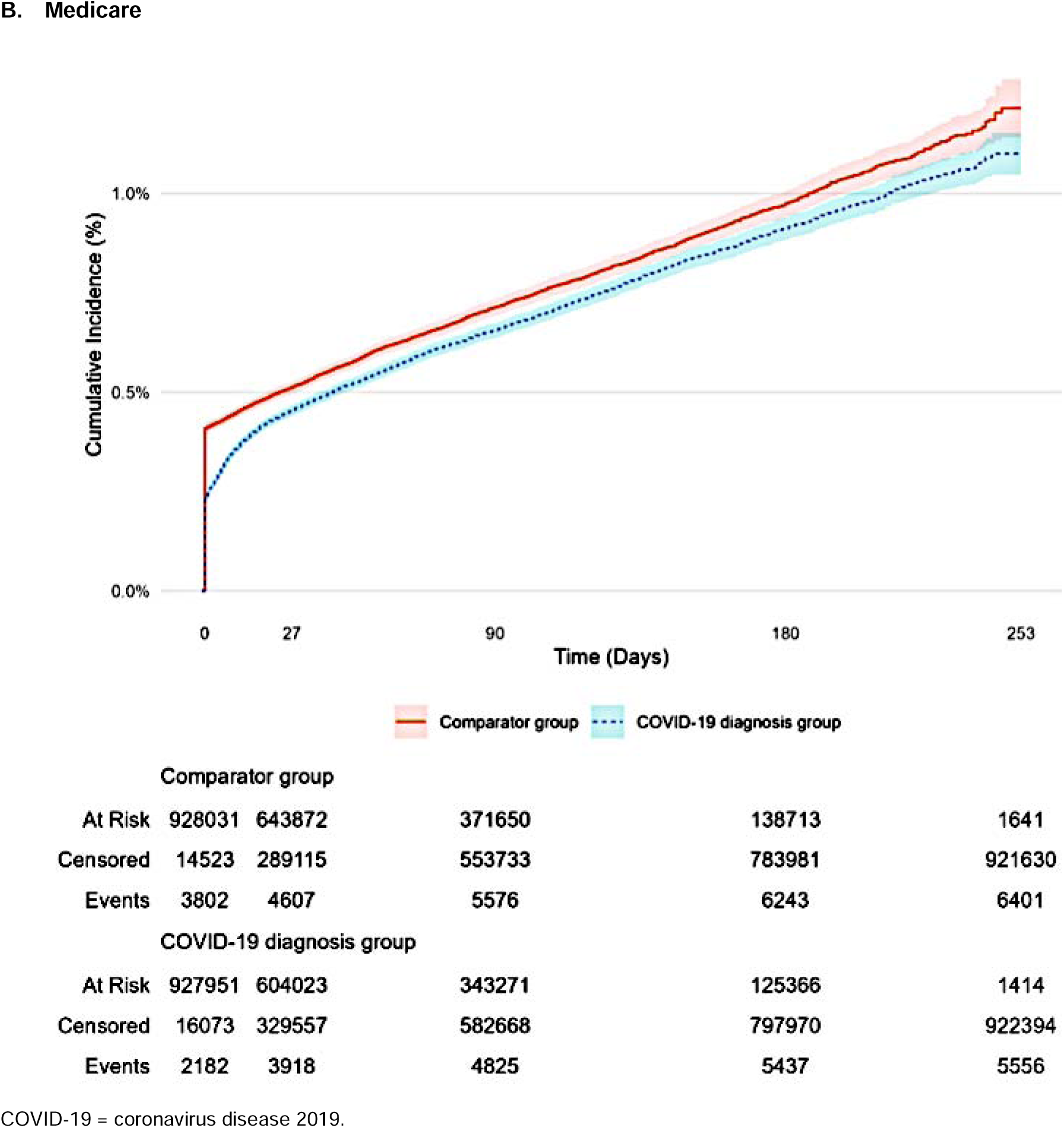
Weighted Cumulative Incidence of Nonhemorrhagic Stroke by Exposure Group in the Cohort Analysis, Starting Follow-Up on Time 0.

**Figure 4.**
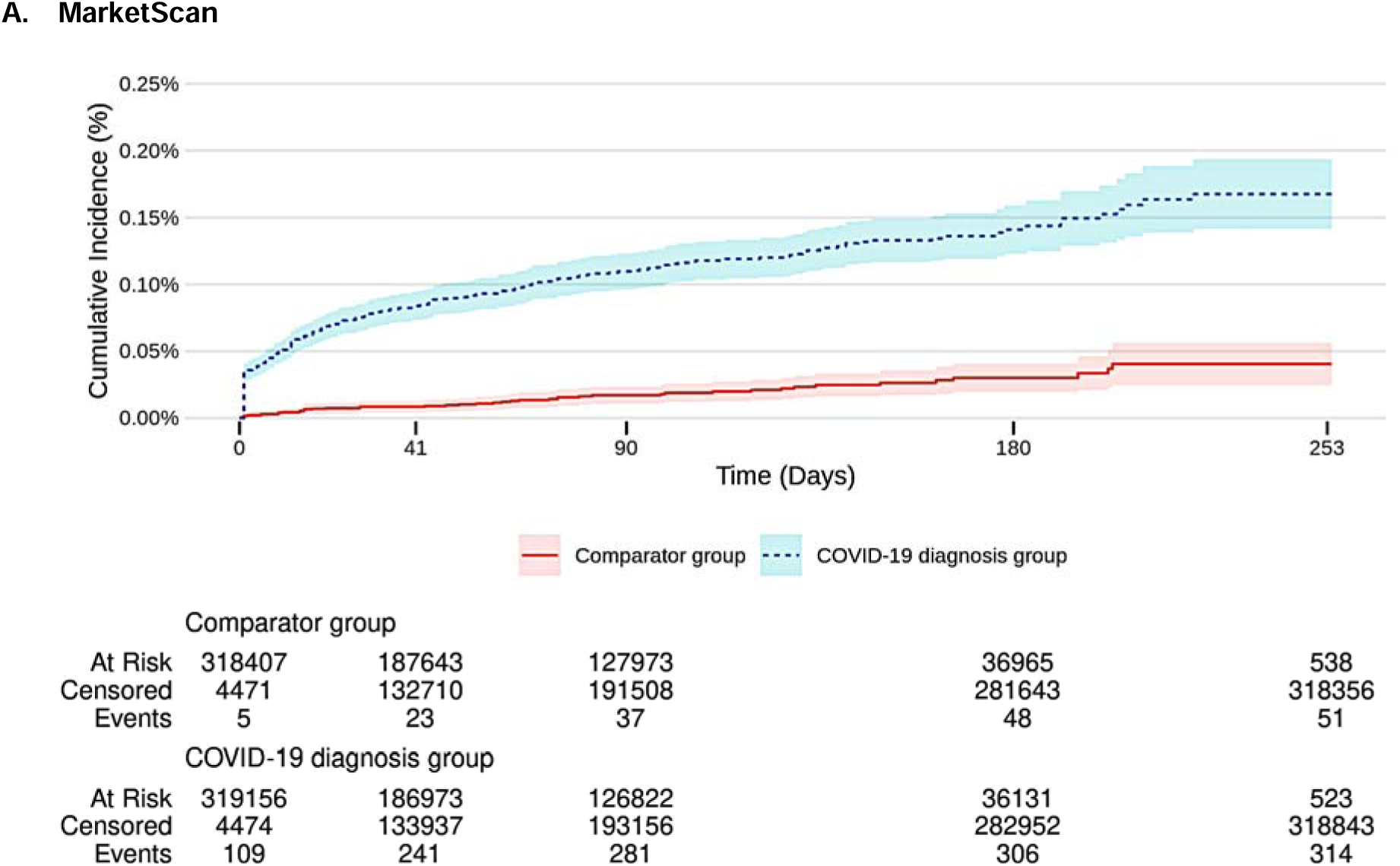
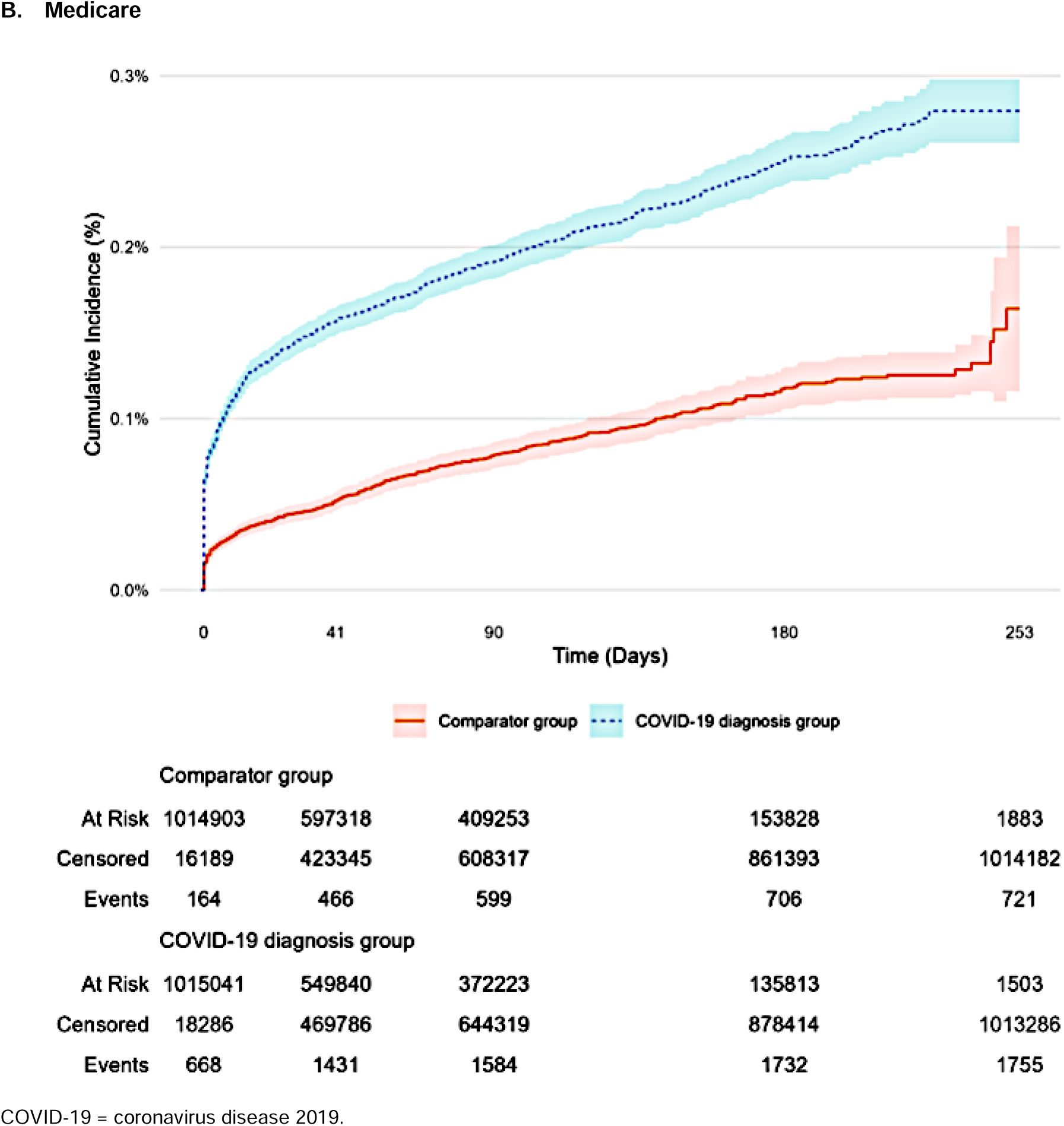
Weighted Cumulative Incidence of Myocarditis/Pericarditis by Exposure Group in the Cohort Analysis, Starting Follow-Up on Time 0.

In subgroup analyses by hospitalization status at Time 0, more differences in baseline characteristics were seen between exposure groups in the hospitalized subgroup, where some comorbidities and many measures of healthcare utilization were more common in the hospitalized comparator group than in the hospitalized COVID-19-diagnosed group (sTable 8). For all AEs, HRs were lower in the hospitalized subgroup than in the not-hospitalized subgroup (sTable 9).

## Discussion

Diagnosis of COVID-19 was associated with a higher risk of cardiovascular AEs in both the cohort analyses and SCRI analyses and in both MarketScan and Medicare. RI estimates ranged from 3.18 (95% CI, 3.05-3.30) for DVT in Medicare to 32.28 (95% CI, 17.06-61.09) for DIC in MarketScan; HR estimates ranged from 1.26 (95% CI, 0.85-1.88) for unusual-site TTS in Medicare to 8.65 (95% CI, 6.06-12.35) for inpatient pulmonary embolism in MarketScan. Estimated associations were weak and inconsistent for appendicitis.

The strength of these observed associations varied by AE as well as by methodological aspects, including when to start follow-up, in both study designs. For the SCRI analyses, increased risk was observed for all AEs (except for appendicitis) in patients with COVID-19 diagnosis, regardless of whether Time 0 was included in the risk window; however, estimates not including Time 0 in the risk window were generally attenuated. For the cohort analyses, the direction and strength of association varied substantially depending on whether Time 0 was included or excluded in the follow-up period. As described in the following sections, HR and RI estimates starting follow-up on Day 1 were used as the primary basis to draw conclusions about the association between COVID-19 diagnoses and AEs.

The association of COVID-19 with cardiovascular events has been suggested by multiple studies from around the world. Studies (including our study) and reviews consistently identified strong associations between COVID-19 and subsequent onset of myocarditis^30^ and pericarditis.^1^ Additionally, relationships with COVID-19 have been suggested for AMI,^1,3,6–8,10,31^ pulmonary embolism,^32–38^ DVT,^38,39^ stroke,^40,41^ and DIC.^42^ Little population-level research exists estimating the association between COVID-19 diagnosis and appendicitis, and we found little evidence to support an association. While the results of our study are generally consistent with many other studies suggesting increased risks of cardiovascular AEs associated with COVID-19, our study utilized 2 large data sources with national coverage, 2 study designs to address various potential biases, and methods to identify and describe potential effects of AE cases occurring on Time 0. Additionally, the estimated attributable risks provide information about the population-level impacts.

### Variation Across Methodological Approaches

In both the SCRI and cohort designs, large numbers of AE events occurred on Time 0, which may suggest the presence of various biases that may manifest in distinct ways (ie, reverse causation, selection bias). Some concurrent cases removed by excluding Time 0 may be true cases where true COVID-19 illness precedes the AE, and investigators may be confronted with difficult decisions about ways to handle Time 0 events: eg, to exclude Time 0 from follow-up to reduce bias, at the expense of precision, generalizability, and complete case ascertainment.

#### Reverse Causation

For some individuals, the occurrence of an AE requiring healthcare attention may result in the recognition and recording of a COVID-19 diagnosis that otherwise would have gone unrecorded. Thus, a form of reverse causation may be present, where clinical presentation of individuals who experience an AE (ie, the study outcome) prompts the recognition and recording of a diagnosis of COVID-19 (ie, the study exposure) thus overestimating the effect of the exposure on the outcome.^13^ As an example in our study, the analyses of myocarditis/pericarditis are consistent with this phenomenon, with large numbers of AEs on Time 0 in both the SCRI and cohort study; and in the cohort study, more AEs occurred on Time 0 in the COVID-19-diagnosed group than the comparator group relative to other nearby days of follow-up (Figure 4, sTable 3, sTable 7). This resulted in strongly elevated RI and HR estimates when Time 0 was included in the risk window/follow-up, and attenuated estimates when Time 0 was excluded. All RI estimates from the SCRI approach were attenuated when the start of follow-up was shifted from Time 0 to Day 1, which suggests (but does not confirm) the possibility of reverse causation and has been previously identified as a concern in SCRI analyses that start follow-up on Time 0.^13^

#### Conditioning on Hospitalization

Matching on hospitalization status at Time 0 was included in the cohort analysis to address disease severity and surveillance bias.^14^ However, conditioning on hospitalization status during periods of pandemic-related healthcare disruptions may have enriched the comparator group with individuals at greater risk of experiencing severe, emergent AEs. If individuals during the study period were less likely to present to clinicians or healthcare facilities for lower-severity events compared to the pre-pandemic period,^43^ the matched hospitalized comparator group may, on average, contain more individuals with high-severity events. Matching on hospitalization status may have inadvertently introduced selection bias by conditioning on a collider on Time 0,^44^ where hospitalization status on Time 0 results from both COVID-19 and occurrence of the AE. Subgroup analyses of patients who were hospitalized at Time 0 revealed that the comparator group in both data sources had a history of more inpatient admissions, ED visits, and risk factors for many of the AEs in this study.

While these factors are expected to affect analyses in both MarketScan and Medicare, this relationship was observed most strongly in Medicare, a population at higher baseline risk of most of the considered AE, as well as severe COVID-19. In Medicare, when follow-up started on Time 0, we observed 4 AEs with paradoxically protective HR estimates in Medicare, appendicitis, unusual-site TTS, HS, and NHS. For each of these AEs, AEs on Time 0 were more common in the comparator group than the COVID-19 group (sTable 7).

### Strengths and Limitations

#### Strengths

Our study has several strengths, including 2 large, diverse data sources analyzed with 2 distinct analytic approaches able to account for changes in incidence rates during the early pandemic period. In the matched cohort study, a high proportion of cases were matched to comparators, and advanced control of observed confounders was achieved using sIPT weights. The use of a younger, commercially insured adult population along with the older Medicare population addresses questions about susceptibility among different adult age groups.

#### Limitations

While 2 large data sources with national reach were included, limitations of the study include a potential lack of generalizability. The study population included individuals with commercial or Medicare fee-for-service insurance, who may not be representative of the general population. Additionally, in the cohort analysis, large numbers of hospitalized COVID-19 cases were excluded due to failing to match, and thus the exposure group may not be reflective of all diagnosed COVID-19 cases. The use of diagnosis codes to identify COVID-19 only captured individuals who received a medically attended diagnosis, which restricts the generalizability of these findings by excluding COVID-19 cases who did not present to healthcare settings. Thus, there were likely individuals in the comparator group with medically undiagnosed COVID-19. The temporal ordering of events is also a limitation common to studies of infectious disease wherein infection occurs before diagnosis and the period between infection and elevated risk of an AE is not always well understood. If diagnosis of COVID-19 is sufficiently delayed, COVID-19-mediated events that occurred prior to diagnosis would be excluded, as they occurred in the pre-diagnosis washout window. Information bias is also an expectation, especially in the Medicare data, as diagnoses from facilities are often dated using the date of admission instead of the date of diagnosis, resulting in more COVID-19 and AE diagnoses being recorded on Time 0.

## Conclusion

Estimates from the cohort and SCRI analyses suggest moderate to very strong associations between COVID-19 diagnosis and cardiovascular AEs. These results suggest that COVID-19 contributed to potentially large numbers of thrombotic cardiovascular AEs in the US during the COVID-19 pandemic. Consistent evidence of an association with appendicitis was not observed. While some variation was observed across study designs, conclusions from both the SCRI and cohort analyses were generally consistent with one another.

We based the overall conclusions for this study on the analyses starting follow-up on Day 1. The influence of AEs occurring on Time 0 was strong for both data sources, with estimates including or excluding Time 0 in follow-up occasionally being in opposite directions. Researchers should consider the risk of bias from sample selection that can be introduced by the study design (eg, matching on collider variables such as hospital admission) or in the analysis (eg, restricting analyses on disease severity). Also, trade-offs between the inclusion and exclusion of Time 0 need to be evaluated and considered.

These analyses add to mounting evidence of the importance of diagnosis and specialty care for CV events associated with COVID-19. Additionally, these findings further demonstrate the impact of COVID-19 on individuals and on public health and provide important context for COVID-19 vaccine safety surveillance and the benefit-risk balance of COVID-19 vaccines and therapeutics.

## Data Availability

The data that support the findings of this study are available from the respective data holders (IBM Consulting, Acumen, LLC), but restrictions apply to the availability of these data, which were used under license for the current study, and so are not publicly available.

## Acknowledgments

The authors acknowledge the support of the following: Sarah Harris, MA, of RTI International for project management and support; Melissa McPheeters, PhD, of RTI International for supervision; Deepa C Youssef, MBA, and Sofia Aschettino, BS, of IBM Consulting for project management and support; Pradeep Rajan, ScD, Marissa Meucci, PhD, and Molly Hensche, MS, of IBM Consulting for study design and protocol development; Veronica Hernandez-Medina, BS, Sania Abhari, MS, and Anna Go, BS, of IBM Consulting, for data acquisition and statistical analysis; Yu Sun, MS, of IBM Consulting, for data acquisition; Pablo Freyria Duenas, MA, and Yangping Chen, MPH, of Acumen LLC, for study design and implementation of statistical analysis.

