## supplemental information for "Evaluating the Risk of Cardiovascular Adverse Events and Appendicitis After COVID-19 Diagnosis in Adults in the United States: Implications of the Start of Follow-Up"

ORIGINAL RESEARCH

J. Bradley Layton et al

| J. Bradley Layton^1^  Arnstein Lindaas^2^  Stella G Muthuri^3^*  Patricia C. Lloyd^4^  Morgan M. Richey^1^*  Joann F. Gruber^4^  Hai Lyu^2^  Mollie M. McKillop^3^*  Lisa S. Kowarski^3^*  Christine Bui^1^  Shelby S. Fisher^5^  Tainya C. Clarke^4^ | Angela S. Cheng^2^*  Zhiruo Wan^2^  Pablo Freyria Duenas^2^  Yangping Chen^2^  Timothy Burrell^3^  Minya Sheng^3^*  Azadeh Shoaibi^4^*  Yoganand Chillarige^2^  Jeffrey Beers^3^  Mary S. Anthony^1^  Richard A. Forshee^4^  Steven A. Anderson^4^ |
| --- | --- |

* Affiliation at time of study

^1^RTI Health Solutions, Research Triangle Park, NC, USA; ^2^Acumen LLC, USA; ^3^IBM Consulting, Bethesda, MD, USA; ^4^US Food and Drug Administration, Center for Biologics Evaluation and Research, Silver Spring, MD, USA; ^5^RTI International, Research Triangle Park, NC, USA.

**Correspondence**: J. Bradley Layton

RTI Health Solutions

3040 East Cornwallis Rd, PO Box 12194

Research Triangle Park, North Carolina, USA 27709

**Source(s) of Support**: US Food and Drug Administration

### Supplemental Table/Figure Legend

sTable 1.	Covariates for AE-Specific Propensity Score Models

sTable 2.	Attrition of Individuals With a COVID-19 Diagnosis for the Self-Controlled Risk Interval Study

A.	MarketScan

B.	Medicare

sTable 3.	Association of a COVID-19 Diagnosis With Adverse Events by Definition of Risk Windows, SCRI Design

sTable 4.	Attrition of Individuals With a COVID-19 Diagnosis for the Cohort Study

A.	MarketScan

B.	Medicare

sTable 5.	Characteristics of Individuals With a COVID-19 Diagnosis Included in the Cohort Study and Those Excluded for Failing to Match

sTable 6.	Characteristics of Individuals With a COVID-19 Diagnosis and Comparator Individuals Without a COVID-19 Diagnosis Included in the Cohort Study

sTable 7.	Association of a COVID-19 Diagnosis With Adverse Events, by Follow-up Period, Cohort Design

sTable 8.	Selected Characteristics of Patients Hospitalized and Not Hospitalized at Time 0 by Exposure Group

A.	MarketScan, Nonhemorrhagic Stroke Analysis Set

B.	Medicare, Overall Cohort

sTable 9.	Association of a COVID-19 Diagnosis With Adverse Events, by Hospitalization Status, Cohort Design

sFigure 1.	Balance of Covariate Distributions Among Individuals With a COVID-19 Diagnosis and Comparator Individuals Without a COVID-19 Diagnosis, Cohort Design, Before and After Stabilized Inverse Probability of Treatment Weighting

A.	Appendicitis, MarketScan

B.	Nonhemorrhagic Stroke, MarketScan

C.	Hemorrhagic Stroke, MarketScan

D.	Acute Myocardial Infarction, MarketScan

E.	Myocarditis/Pericarditis, MarketScan

F.	Deep Vein Thrombosis, MarketScan

G.	Pulmonary Embolism, MarketScan

H.	Disseminated Intravascular Coagulation, MarketScan

I.	Unusual-Site Thrombosis With Thrombocytopenia Syndrome, MarketScan

J.	Common-Site Thrombosis With Thrombocytopenia Syndrome, MarketScan

K.	Appendicitis, Medicare

L.	Nonhemorrhagic Stroke, Medicare

M.	Hemorrhagic Stroke, Medicare

N.	Acute Myocardial Infarction, Medicare

O.	Myocarditis/Pericarditis, Medicare

P.	Deep Vein Thrombosis, Medicare

Q.	Pulmonary Embolism, Medicare

R.	Disseminated Intravascular Coagulation, Medicare

S.	Unusual-Site Thrombosis With Thrombocytopenia Syndrome, Medicare

T.	Common-Site Thrombosis With Thrombocytopenia Syndrome, Medicare

### Supplemental Information

sTable 1. Covariates for AE-Specific Propensity Score Models

| Covariate | Model form | Nonhemorrhagic stroke | Hemorrhagic stroke | Acute myocardial infarction | Myocarditis/pericarditis | Deep vein thrombosis | Pulmonary embolism ^a^ | Disseminated intravascular coagulation | Unusual-site TTS | Common-site TTS | Appendicitis |
| --- | --- | --- | --- | --- | --- | --- | --- | --- | --- | --- | --- |
| Age ^b^ | Age, age^2^, age^3^ | × | × | × | × | × | × | × | × | × | × |
| Sex ^b^ | Binary | × | × | × | × | × | × | × | × | × | × |
| County/MSA and state of residence ^b^ | Categorical, indicator for each level | × | × | × | × | × | × | × | × | × | × |
| Race/ethnicity ^c^ | Categorical, indicator for each level | × | × | × | × | × | × | × | × | × | × |
| Dual Medicare/Medicaid eligibility ^c^ | Binary | × | × | × | × | × | × | × | × | × | × |
| Reason for entering Medicare ^c^ | Categorical, indicator for each level | × | × | × | × | × | × | × | × | × | × |
| Hospitalization status on Time 0 ^b^ | Binary | × | × | × | × | × | × | × | × | × | × |
| SNF/LTC residence on Time 0 ^b, c^ | Binary | × | × | × | × | × | × | × | × | × | × |
| Animal exposure/bites or rabies | Binary |  |  |  | × |  |  |  |  |  |  |
| Antiphospholipid syndrome | Binary |  |  |  |  |  |  |  | × | × |  |
| Autoimmune disorders | Binary | × | × | × | × | × | × | × | × | × |  |
| Brain lesions related to secondary narcolepsy | Binary |  |  |  |  |  |  |  |  |  |  |
| Cancer | Binary | × | × | × | × | × | × | × | × | × |  |
| Chronic kidney disease/renal disease | Binary | × | × | × | × | × | × | × | × | × |  |
| End-stage renal disease ^c^ | Binary | × | × | × | × | × | × | × | × | × | × |
| Chronic liver disease | Binary | × | × | × | × | × | × | × |  |  |  |
| Chronic lung disease | Binary | × | × | × |  |  |  |  | × | × |  |
| Dementia/neurologic conditions | Binary | × | × |  |  | × | × | × | × | × |  |
| Diabetes mellitus, type 1 or 2 | Binary | × | × | × | × | × | × |  | × | × |  |
| Disseminated intravascular coagulation | Binary |  |  |  |  |  |  |  | × | × |  |
| Heart conditions | Binary | × | × | × | × | × | × |  | × | × |  |
| Hemiplegia/paraplegia | Binary | × | × |  |  | × | × |  | × | × |  |
| Herpes simplex virus infection | Binary |  |  |  | × |  |  |  |  |  |  |
| Hypertension | Binary | × | × | × | × |  |  |  |  |  |  |
| Immunocompromised state ^b^ | Binary | × | × | × | × | × | × | × | × | × | × |
| Infection associated with myocarditis/pericarditis | Binary |  |  |  | × |  |  |  |  |  |  |
| Inpatient surgery | Binary |  |  |  |  | × | × | × | × | × |  |
| Lipid abnormality | Binary | × |  | × |  | × | × |  | × | × |  |
| Mental health conditions | Binary |  | × | × |  |  |  |  |  |  |  |
| Nutritional deficiencies | Binary |  |  |  | × |  |  |  |  |  |  |
| Obesity | Binary | × |  | × | × | × | × |  | × | × |  |
| Peripheral vascular disease | Binary | × | × | × | × | × | × |  | × | × |  |
| Pneumonia or lower respiratory tract infection | Binary | × | × | × | × | × | × | × | × | × | × |
| Pregnancy ^d^ | Binary | × | × | × |  | × | × | × | × | × |  |
| Sepsis | Binary |  |  |  | × |  |  | × |  |  |  |
| Sickle cell disease/thalassemia | Binary | × | × | × |  | × | × |  |  | × |  |
| Smoking/nicotine dependency | Binary | × | × | × |  | × | × | × | × | × |  |
| Stroke/cerebrovascular disease | Binary | × | × | × |  | × | × |  |  | × |  |
| Thromboembolism | Binary |  |  |  |  | × | × | × | × | × |  |
| Thrombophilia | Binary | × |  | × |  | × | × | × | × | × |  |
| Trauma | Binary | × | × |  | × | × | × | × |  | × |  |
| Tuberculosis | Binary |  |  |  | × | × |  |  | × | × |  |
| Inpatient hospital stays before Time 0 | Categorical, indicator for each level | × | × | × | × | × | × | × | × | × | × |
| ED visits | Categorical, indicator for each level | × | × | × | × | × | × | × | × | × | × |
| Outpatient visits | Categorical, indicator for each level | × | × | × | × | × | × | × | × | × | × |
| SNF/LTC stay ^e^ | Binary | × | × | × |  | × | × | × | × | × |  |
| Influenza vaccination in previous year | Binary | × | × | × | × | × | × | × | × | × | × |

AE = adverse event; ED = emergency department; ENC = encephalitis/encephalomyelitis; GBS = Guillain-Barré syndrome; HS = hemorrhagic stroke; ITP = immune thrombocytopenia; LTC = long-term care; MSA = metropolitan statistical area; SNF = skilled nursing facility; TM = transverse myelitis; TTS = thrombosis with thrombocytopenia syndrome.

^a^ Evaluated both as inpatient only, and overall. Both outcomes used the same propensity score model in the same analysis set.

^b^ Matching variables were included in all propensity score models.

^c^ Only included in Medicare.

^d^ Only included in MarketScan.

^e^ SNF and LTC both included in Medicare; only SNF included in MarketScan.

sTable 2. Attrition of Individuals With a COVID-19 Diagnosis for the Self-Controlled Risk Interval Study
A. MarketScan

| Characteristic | Value |
| --- | --- |
| Overall SCRI population |  |
| Individuals with COVID-19 diagnosis during the study period | 509,875 |
| Excluded for being aged outside age range, N (%) | 51,848 (10.2) |
| Excluded for lacking 365 days of continuous database enrollment before Time 0, n (%) | 128,310 (25.2) |
| Excluded for having a previous COVID-19 diagnosis | 2,135 (0.4) |
| Excluded for having a previous select respiratory infection | 119 (0.0) |
| Excluded for lacking ≥ 1 day in the pre-exposure reference window | 25,306 (5.0) |
| Total SCRI population | 330,799 |
| Outcome-specific analysis sets with 42-day risk windows ^a^ |  |
| Excluded for not having appendicitis in risk or reference window | 330,453 (99.9) |
| Excluded for lacking 365 days of continuous database enrollment before outcome date | 0 (0.0) |
| Excluded for having appendicitis in outcome washout window | 24 (0.0) |
| Appendicitis analysis set | 296 |
| Excluded for not having myocarditis/pericarditis in risk or reference window | 330,392 (99.9) |
| Excluded for lacking 365 days of continuous database enrollment before outcome date | 0 (0.0) |
| Excluded for having myocarditis/pericarditis in outcome washout window | 27 (0.0) |
| Myocarditis/pericarditis analysis set | 339 |
| Outcome-specific analysis sets with 28-day risk windows ^a^ |  |
| Excluded for not having deep vein thrombosis in risk or reference window | 329,228 (99.5) |
| Excluded for lacking 365 days of continuous database enrollment before outcome date | 0 (0.0) |
| Excluded for having deep vein thrombosis in outcome washout window | 374 (0.1) |
| Deep vein thrombosis analysis set | 1,013 |
| Excluded for not having pulmonary embolism, inpatient, in risk or reference window | 330,062 (99.8) |
| Excluded for lacking 365 days of continuous database enrollment before outcome date | 0 (0.0) |
| Excluded for having pulmonary embolism in outcome washout window | 118 (0.0) |
| Pulmonary embolism, inpatient, analysis set | 560 |
| Excluded for not having pulmonary embolism, overall, in risk or reference window | 328,773 (99.4) |
| Excluded for lacking 365 days of continuous database enrollment before outcome date | 0 (0.0) |
| Excluded for having pulmonary embolism in outcome washout window | 382 (0.1) |
| Pulmonary embolism, overall, analysis set | 1,444 |
| Excluded for not having disseminated intravascular coagulation in risk or reference window | 330,748 (100.0) |
| Excluded for lacking 365 days of continuous database enrollment before outcome date | 0 (0.0) |
| Excluded for having disseminated intravascular coagulation in outcome washout window | 0 (0.0) |
| Disseminated intravascular coagulation analysis set | 42 |

B. Medicare

| Characteristic | Value |
| --- | --- |
| Overall SCRI population |  |
| Individuals with COVID-19 diagnosis during the study period | 1,571,901 |
| Excluded for being aged outside age range, n (%) | 219,427 (14.0) |
| Excluded for lacking 365 days of continuous database enrollment before Time 0, n (%) | 366,363 (23.3) |
| Excluded for having a previous COVID-19 diagnosis | 85,728 (5.5) |
| Excluded for having a previous select respiratory infection | 407 (0.0) |
| Excluded for lacking ≥ 1 day in the pre-exposure reference window | 44,911 (2.9) |
| Total SCRI population | 855,065 (54.4) |
| Outcome-specific analysis sets with 42-day risk windows ^a^ |  |
| Excluded for not having appendicitis in risk or reference window | 854,633 (99.9) |
| Excluded for lacking 365 days of continuous database enrollment before outcome date | 0 (0.0) |
| Excluded for having appendicitis in outcome washout window | 36 (0.0) |
| Appendicitis analysis set | 396 (0.0) |
| Excluded for not having myocarditis/pericarditis in risk or reference window | 853,313 (99.8) |
| Excluded for lacking 365 days of continuous database enrollment before outcome date | < 11 |
| Excluded for having myocarditis/pericarditis in outcome washout window | > 11 |
| Myocarditis/pericarditis analysis set | 1,605 (0.2) |
| Outcome-specific analysis sets with 28-day risk windows ^a^ |  |
| Excluded for not having deep vein thrombosis in risk or reference window | 828,927 (96.9) |
| Excluded for lacking 365 days of continuous database enrollment before outcome date | 13 (0.0) |
| Excluded due to having deep vein thrombosis in outcome washout window | 9,079 (1.1) |
| Deep vein thrombosis analysis set | 17,046 (2.0) |

COVID-19 = coronavirus disease 2019; SCRI = self-controlled risk interval.

^a^ Denominators for all outcome-specific exclusions are the total SCRI after excluding those with ≥ 1 day in risk and reference windows; exclusions are noncumulative across analysis sets.

Note: Privacy rules require masking cell sizes containing fewer than 11 individuals.

sTable 3. Association of a COVID-19 Diagnosis With Adverse Events by Definition of Risk Windows, SCRI Design

| Outcomes | Risk window | Cases in risk window | Cases in reference window | RI (95% CI) |
| --- | --- | --- | --- | --- |
| MarketScan | | | | |
| Myocarditis/pericarditis | Time 0 – end | 274 | 65 | 24.64 (17.27-35.17) |
|  | Time 0 | 128 | 65 | 260.47 (208.27-325.76) |
|  | Day 1 – end | 146 | 65 | 9.03 (7.22-11.29) |
| Deep vein thrombosis | Time 0 – end | 627 | 386 | 11.80 (10.11-13.77) |
|  | Time 0 | 177 | 386 | 64.62 (54.77-76.25) |
|  | Day 1 – end | 450 | 386 | 8.04 (7.07-9.14) |
| Pulmonary embolism – any setting | Time 0 – end | 1,136 | 308 | 30.49 (25.95-35.83) |
|  | Time 0 | 503 | 308 | 227.88 (204.11-254.42) |
|  | Day 1 – end | 633 | 308 | 13.55 (12.17-15.09) |
| Pulmonary embolism – inpatient only | Time 0 – end | 493 | 67 | 100.95 (67.05-151.97) |
|  | Time 0 | 245 | 67 | 556.35 (469.18-659.72) |
|  | Day 1 – end | 248 | 67 | 27.36 (23.01-32.55) |
| Disseminated intravascular coagulation | Time 0 – end | 33 | 9 | 83.52 (18.72-372.74) |
|  | Time 0 | 12 | 9 | 361.01 (181.34-718.70) |
|  | Day 1 – end | 21 | 9 | 32.28 (17.06-61.09) |
| Appendicitis | Time 0 – end | 140 | 156 | 5.00 (3.75-6.67) |
|  | Time 0 | 95 | 156 | 91.27 (71.38-116.70) |
|  | Day 1 – end | 45 | 156 | 1.38 (1.00-1.90) |
| Medicare | | | | |
| Myocarditis/pericarditis | Time 0 – end | 1,176 | 429 | 11.73 (10.05-13.69) |
|  | Time 0 | 584 | 429 | 118.00 (102.10-136.39) |
|  | Day 1 – end | 592 | 429 | 4.53 (3.89-5.27) |
| Deep vein thrombosis | Time 0 – end | 9,068 | 7,978 | 5.58 (5.38-5.79) |
|  | Time 0 | 3,608 | 7,978 | 43.61 (41.78-45.51) |
|  | Day 1 – end | 5,460 | 7,978 | 3.18 (3.05-3.30) |
| Appendicitis | Time 0 – end | 163 | 233 | 3.53 (2.73-4.56) |
|  | Time 0 | 111 | 233 | 59.69 (46.24-77.04) |
|  | Day 1 – end | 52 | 233 | 0.98 (0.70-1.36) |

CI = confidence interval; RI = relative incidence.

Note: Models were adjusted for calendar month to account for potential seasonality and time trends

sTable 4. Attrition of Individuals With a COVID-19 Diagnosis for the Cohort Study
A. MarketScan

| Characteristic | Value |
| --- | --- |
| Overall study cohort |  |
| Individuals with COVID-19 diagnosis during the study period | 569,754 |
| Excluded for being aged outside age range, n (%) | 53,723 (9.43) |
| Excluded for lacking 365 days of continuous data base enrollment before Time 0, n (%) | 142,532 (25.02) |
| Excluded for lacking ≥ 1 healthcare claim during 365 days before Time 0, n (%) | 51,066 (8.96) |
| Excluded for having a COVID-19 diagnosis before Time 0, n (%) | 7,002 (1.23) |
| Excluded for having a select respiratory infection before Time 0, n (%) | 382 (0.07) |
| Eligible adults with new-onset COVID-19 | 358,306 |
| Excluded for failing to match, n (%) | 39,006 (10.89) |
| Matched individuals with COVID-19 | 319,300 |
| Matched comparator observations ^a^ (unique individuals) | 319,300 (312,282) |
| Total study cohort | 638,600 |
| Outcome-specific analysis sets ^b^ |  |
| Excluded for having previous appendicitis | 1,295 (0.20) |
| Appendicitis analysis set | 637,305 |
| Excluded for having previous nonhemorrhagic stroke | 2,697 (0.42) |
| Nonhemorrhagic stroke analysis set | 635,903 |
| Excluded for having previous hemorrhagic stroke | 539 (0.08) |
| Hemorrhagic stroke analysis set | 638,061 |
| Excluded for having previous acute myocardial infarction | 2,010 (0.31) |
| Acute myocardial infarction analysis set | 636,590 |
| Excluded for having previous myocarditis pericarditis | 341 (0.05) |
| Myocarditis/pericarditis analysis set | 638,259 |
| Excluded for having previous deep vein thrombosis | 2,558 (0.40) |
| Deep vein thrombosis analysis set | 636,042 |
| Excluded for having previous pulmonary embolism | 1,869 (0.29) |
| Pulmonary embolism analysis set ^c^ | 636,731 |
| Excluded for having previous disseminated intravascular coagulation | 57 (0.01) |
| Disseminated intravascular coagulation analysis set | 638,543 |
| Excluded for having previous unusual-site thrombosis with thrombocytopenia | 47 (0.01) |
| Unusual-site thrombosis with thrombocytopenia analysis set | 638,553 |
| Excluded for having previous common-site thrombosis with thrombocytopenia | 281 (0.04) |
| Common-site thrombosis with thrombocytopenia analysis set | 638,319 |

B. Medicare

| Characteristic | Value |
| --- | --- |
| Overall study cohort |  |
| Individuals with COVID-19 diagnosis during the study period | 1,807,670 |
| Excluded for being aged outside age range, n (%) | 253,377 (14.0%) |
| Excluded for lacking 365 days of continuous data base enrollment before Time 0, n (%) | 436,595 (24.2%) |
| Excluded for lacking ≥ 1 healthcare claim during 365 days before Time 0, n (%) | 9,722 (0.5%) |
| Excluded for having a COVID-19 diagnosis before Time 0, n (%) | 10,510 (0.6%) |
| Excluded for having a select respiratory infection before Time 0, n (%) | 12,048 (0.7%) |
| Eligible adults with new-onset COVID-19 | 1,085,418 |
| Excluded for failing to match, n (%) | 68,008 (6.3%) |
| Matched individuals with COVID-19 | 1,017,410 |
| Matched comparator observations ^a^ (unique individuals) | 1,017,410 (930,513) |
| Total study cohort | 2,034,820 |
| Outcome-specific analysis sets ^b^ |  |
| Excluded for having previous appendicitis | 2,771 (0.1%) |
| Appendicitis analysis set | 2,032,049 (99.9%) |
| Excluded for having previous nonhemorrhagic stroke | 177,011 (8.7%) |
| Nonhemorrhagic stroke analysis set | 1,857,809 (91.3%) |
| Excluded for having previous hemorrhagic stroke | 28,459 (1.4%) |
| Hemorrhagic stroke analysis set | 2,006,361 (98.6%) |
| Excluded for having previous acute myocardial infarction | 83,837 (4.1%) |
| Acute myocardial infarction analysis set | 1,950,983 (95.9%) |
| Excluded for having previous myocarditis pericarditis | 3,297 (0.2%) |
| Myocarditis/pericarditis analysis set | 2,031,523 (99.8%) |
| Excluded for having previous deep vein thrombosis | 71,089 (3.5%) |
| Deep vein thrombosis analysis set | 1,963,731 (96.5%) |
| Excluded for having previous pulmonary embolism | 40,225 (2.0%) |
| Pulmonary embolism analysis set ^c^ | 1,994,595 (98.0%) |
| Excluded for having previous disseminated intravascular coagulation | 1,204 (0.1%) |
| Disseminated intravascular coagulation analysis set | 2,033,616 (99.9%) |
| Excluded for having previous unusual-site thrombosis with thrombocytopenia | 783 (0.0%) |
| Unusual-site thrombosis with thrombocytopenia analysis set | 2,034,037 (100.0%) |
| Excluded for having previous common-site thrombosis with thrombocytopenia | 22,400 (1.1%) |
| Common-site thrombosis with thrombocytopenia analysis set | 2,012,420 (98.9%) |

COVID-19 = coronavirus disease 2019.

^a^ A unique individual may be included multiple times because of matching with replacement, and an individual may be included in both the exposure and comparator groups. This table counts each instance of an individual’s entry separately as distinct observations; the number of unique individuals is reported separately.

^b^ Denominators for all outcome-specific exclusions are the total study cohort; exclusions are noncumulative across analysis sets.

^c^ Both pulmonary embolism outcomes (ie, inpatient-only, and in any setting) were evaluated in the same analysis set.

sTable 5. Characteristics of Individuals With a COVID-19 Diagnosis Included in the Cohort Study and Those Excluded for Failing to Match

| Characteristic | MarketScan | | Medicare | |
| --- | --- | --- | --- | --- |
|  | COVID-19 diagnosis (exposure) group retained by matching | Individuals with a COVID-19 diagnosis who failed to match | COVID-19 diagnosis (exposure) group retained by matching | Individuals with a COVID-19 diagnosis who failed to match |
| Total unique individuals | 319,300 | 39,006 | 1,017,410 | 68,008 |
| Demographic characteristics |  |  |  |  |
| Age, years |  |  |  |  |
| Mean (SD) | 41.8 (13.6) | 44.1 (13.6) | 77.7 (8.6) | 80.8 (9.0)) |
| Median (Q1, Q3) | 43 (30, 53) | 47 (33, 56) | 76.0 (71.0, 84.0) | 81.0 (73.0, 88.0) |
| Age groups, n (%) |  |  |  |  |
| 18-34 years | 105,842 (33.15) | 10,485 (26.88) | NA | NA |
| 35-49 years | 101,071 (31.65) | 12,060 (30.92) | NA | NA |
| 50-64 years | 112,387 (35.20) | 16,461 (42.20) | NA | NA |
| 65-79 years | NA | NA | 629,709 (61.9) | 31,064 (45.7) |
| ≥ 80 years | NA | NA | 387,701 (38.1) | 36,944 (54.3) |
| Sex, n (%) |  |  |  |  |
| Female | 178,975 (56.05) | 22,793 (58.43) | 590,584 (58.0) | 34,257 (50.4) |
| Male | 140,325 (43.95) | 16,213 (41.57) | 426,826 (42.0) | 33,751 (49.6) |
| US geographic region, n (%) |  |  |  |  |
| Northeast | 62,215 (19.48) | 949 (2.43) | 218,277 (21.5) | 9,103 (13.4) |
| North central | 71,170 (22.29) | 1,788 (4.58) | 250,139 (24.6) | 19,879 (29.2) |
| South | 149,868 (46.94) | 26,883 (68.92) | 385,012 (37.8) | 31,590 (46.5) |
| West | 36,037 (11.29) | 9,057 (23.22) | 161,002 (15.8) | 7,093 (10.4) |
| Unknown | 10 (0.00) | 329 (0.84) | 2,980 (0.3) | 343 (0.5) |
| Race/ethnicity, n (%) | NA | NA |  |  |
| American Indian/Alaska Native |  |  | 7,510 (0.7) | 928 (1.4) |
| Asian |  |  | 16,819 (1.7) | 904 (1.3) |
| Black or African American |  |  | 90,997 (8.9) | 9,789 (14.4) |
| Hispanic/Latin American/Latinx |  |  | 26,460 (2.6) | 1,640 (2.4) |
| White |  |  | 845,042 (83.1) | 53,588 (78.8) |
| A race/ethnicity not listed |  |  | 13,898 (1.4) | 730 (1.1) |
| Not provided |  |  | 16,684 (1.6) | 429 (0.6) |
| Dual Medicare/Medicaid eligibility, n (%) | NA | NA | 297,499 (29.2) | 30,409 (44.7) |
| Original reason for Medicare eligibility, n (%) | NA | NA |  |  |
| Age |  |  | 860,430 (84.6) | 53,381 (78.5) |
| Disability |  |  | 150,916 (14.8) | 14,052 (20.7) |
| End-stage renal disease |  |  | 6,064 (0.6) | 575 (0.8) |
| Hospitalized on Time 0, n (%) | 4,046 (1.27) | 8,738 (22.40) | 166,154 (16.3) | 67,764 (99.6) |
| SNF/LTC ^a^ residence on Time 0, n (%) | NA | NA | 250,193 (24.6) | 33,311 (49.0) |
| Comorbidities, n (%) |  |  |  |  |
| Animal exposure/bites or rabies | 976 (0.31) | 124 (0.32) | 3,105 (0.3) | 130 (0.2) |
| Antiphospholipid syndrome | 160 (0.05) | 28 (0.07) | 829 (0.1) | 40 (0.1) |
| Autoimmune disorders | 12,677 (3.97) | 1,759 (4.51) | 93,071 (9.1) | 5,722 (8.4) |
| Brain lesions related to secondary narcolepsy | 30 (0.01) | 2 (0.01) | 519 (0.1) | 54 (0.1) |
| Cancer | 13,421 (4.20) | 1,987 (5.09) | 272,867 (26.8) | 16,744 (24.6) |
| Chronic lymphocytic leukemia | 213 (0.07) | 41 (0.11) | 6,995 (0.7) | 576 (0.8) |
| Chronic kidney disease or renal disease (other than end-stage renal disease) | 6,052 (1.90) | 1,495 (3.83) | 276,654 (27.2) | 29,364 (43.2) |
| End-stage renal disease | NA | NA | 20,229 (2.0) | 2,489 (3.7) |
| Chronic liver disease | 10,763 (3.37) | 1,566 (4.01) | 80,781 (7.9) | 5,759 (8.5) |
| Chronic lung diseases | 32,616 (10.21) | 4,285 (10.99) | 293,426 (28.8) | 25,836 (38.0) |
| Dementia or other neurologic condition | 19,722 (6.18) | 2,551 (6.54) | 350,351 (34.4) | 34,022 (50.0) |
| Diabetes, Type 1 or 2 | 33,117 (10.37) | 6,374 (16.34) | 406,448 (39.9) | 33,886 (49.8) |
| Disseminated intravascular coagulation ^a^ | 24 (0.01) | 9 (0.02) | 324 (0.0) | 28 (0.0) |
| Heart conditions | 37,470 (11.74) | 5,494 (14.09) | 614,855 (60.4) | 49,731 (73.1) |
| Hemiplegia or paraplegia | 857 (0.27) | 202 (0.52) | 61,853 (6.1) | 7,582 (11.1) |
| Herpes simplex infection | 6,082 (1.90) | 709 (1.82) | 16,959 (1.7) | 948 (1.4) |
| Hypertension | 78,717 (24.65) | 13,371 (34.28) | 823,414 (80.9) | 59,938 (88.1) |
| Immunocompromised state | 37,672 (11.80) | 5,422 (13.90) | 438,954 (43.1) | 27,919 (41.1) |
| Infection associated with GBS | 17,961 (5.63) | 2,091 (5.36) | 51,279 (5.0) | 4,484 (6.6) |
| Viral infection associated with ITP | 1,290 (0.40) | 209 (0.54) | 3,702 (0.4) | 237 (0.3) |
| Infection associated with myocarditis/pericarditis | 16,753 (5.25) | 2,322 (5.95) | 111,923 (11.0) | 11,207 (16.5) |
| Inpatient surgery | 1,985 (0.62) | 379 (0.97) | 43,070 (4.2) | 5,421 (8.0) |
| Lipid abnormality | 73,480 (23.01) | 10,908 (27.96) | 733,659 (72.1) | 48,762 (71.7) |
| Mental health conditions | 73,913 (23.15) | 10,422 (26.72) | 441,954 (43.4) | 35,803 (52.6) |
| Nutritional deficiency | 2,088 (0.65) | 374 (0.96) | 80,039 (7.9) | 9,196 (13.5) |
| Obese or severely obese | 63,585 (19.91) | 9,775 (25.06) | 233,833 (23.0) | 16,401 (24.1) |
| Peripheral vascular disease | 4,393 (1.38) | 733 (1.88) | 318,384 (31.3) | 28,432 (41.8) |
| Pneumonia or lower respiratory tract infection | 41,355 (12.95) | 5,598 (14.35) | 237,118 (23.3) | 22,636 (33.3) |
| Pregnancy | 2,616 (0.82) | 266 (0.68) | NA | NA |
| Sepsis | 1,277 (0.40) | 340 (0.87) | 59,822 (5.9) | 8,147 (12.0) |
| Sickle cell disease or thalassemia | 750 (0.23) | 112 (0.29) | 2,611 (0.3) | 177 (0.3) |
| Smoking/nicotine dependency | 11,536 (3.61) | 1,391 (3.57) | 64,705 (6.4) | 5,848 (8.6) |
| Stroke or cerebrovascular disease | 4,730 (1.48) | 715 (1.83) | 228,802 (22.5) | 21,214 (31.2) |
| Thromboembolism | 3,531 (1.11) | 642 (1.65) | 77,562 (7.6) | 7,678 (11.3) |
| Thrombophilia | 1,096 (0.34) | 137 (0.35) | 11,107 (1.1) | 763 (1.1) |
| Trauma | 204 (0.06) | 38 (0.10) | 24,546 (2.4) | 3,371 (5.0) |
| Tuberculosis | 138 (0.04) | 21 (0.05) | 2,585 (0.3) | 218 (0.3) |
| Healthcare utilization, n (%) |  |  |  |  |
| Inpatient stays in previous 365 days |  |  |  |  |
| 0 | 305,018 (95.53) | 36,528 (93.65) | 729,890 (71.7) | 37,200 (54.7) |
| 1-2 | 13,162 (4.12) | 2,138 (5.48) | 204,101 (20.1) | 19,704 (29.0) |
| ≥ 3 | 1,120 (0.35) | 340 (0.87) | 83,419 (8.2) | 11,104 (16.3) |
| ED visits in previous 365 days |  |  |  |  |
| 0 | 255,758 (80.10) | 30,760 (78.86) | 572,454 (56.3) | 26,433 (38.9) |
| 1-2 | 54,798 (17.16) | 7,022 (18.00) | 282,442 (27.8) | 22,149 (32.6) |
| ≥ 3 | 8,744 (2.74) | 1,224 (3.14) | 162,514 (16.0) | 19,426 (28.6) |
| Outpatient provider visits ^b^ in previous 365 days |  |  |  |  |
| 0 | 2,076 (0.65) | 248 (0.64) | 93 (0.0) | 13 (0.0) |
| 1-2 | 56,880 (17.81) | 5,749 (14.74) | 21,024 (2.1) | 1,189 (1.7) |
| 3-5 | 76,244 (23.88) | 7,326 (18.78) | 51,668 (5.1) | 2,598 (3.8) |
| 6-10 | 76,772 (24.04) | 8,223 (21.08) | 118,531 (11.7) | 5,352 (7.9) |
| ≥ 11 | 107,328 (33.61) | 17,460 (44.76) | 826,094 (81.2) | 58,856 (86.5) |
| SNF/LTC stay ^c^ | 341 (0.11) | 104 (0.27) | 295,527 (29.0) | 36,920 (54.3) |
| Influenza vaccination in previous year | 90,018 (28.19) | 12,166 (31.19) | 570,066 (56.0) | 33,767 (49.7) |

COVID‑19 = coronavirus disease 2019; ED = emergency department; GBS = Guillain-Barré syndrome; ITP = immune thrombocytopenia; LTC = long-term care; Q1 = first quartile, Q3 = third quartile; SD = standard deviation; SNF = skilled nursing facility; US = United States.

^a^ This serves as an outcome-specific exclusion criterion when the condition is evaluated as an outcome, but history of the condition serves as a covariate for analyses of other outcomes.

^b^ Unique days on which an outpatient visit occurred.

^c^ SNF and LTC both identified in Medicare; SNF identified in MarketScan only.

sTable 6. Characteristics of Individuals With a COVID-19 Diagnosis and Comparator Individuals Without a COVID-19 Diagnosis Included in the Cohort Study

| Characteristic | MarketScan | | | Medicare | | |
| --- | --- | --- | --- | --- | --- | --- |
|  | COVID-19 diagnosis (exposure) group | Comparator group | ASD | COVID-19 diagnosis (exposure) group | Comparator group | ASD |
| Total selected observations ^a^ (unique individuals) | 319,300 (319,300) | 319,300 (312,282) |  | 1,017,410 (1,017,410) | 1,017,410 (930,513) |  |
| Demographic characteristics |  |  |  |  |  |  |
| Age, years |  |  |  |  |  |  |
| Mean (SD) | 41.8 (13.6) | 41.8 (13.6) | 0.00 | 77.7 (8.6) | 77.7 (8.6) | 0.00 |
| Median (Q1, Q3) | 43 (30, 53) | 43 (30, 53) |  | 76 (71, 84) | 76 (71, 84) |  |
| Age groups, n (%) |  |  |  |  |  |  |
| 18-34 years | 105,842 (33.15) | 105,842 (33.15) |  |  |  |  |
| 35-49 years | 101,071 (31.65) | 101,071 (31.65) |  |  |  |  |
| 50-64 years | 112,387 (35.20) | 112,387 (35.20) |  |  |  |  |
| 65-79 years |  |  |  | 629,709 (61.9) | 629,709 (61.9) |  |
| ≥ 80 years |  |  |  | 387,701 (38.1) | 387,701 (38.1) |  |
| Sex, n (%) |  |  | 0.00 |  |  | 0.00 |
| Female | 178,975 (56.05) | 178,975 (56.05) |  | 590,584 (58.0) | 590,584 (58.0) |  |
| Male | 140,325 (43.95) | 140,325 (43.95) |  | 426,826 (42.0) | 426,826 (42.0) |  |
| US geographic region, n (%) |  |  | 0.00 |  |  | 0.00 |
| Northeast | 62,215 (19.48) | 62,224 (19.49) |  | 218,277 (21.5) | 218,107 (21.4) |  |
| North Central | 71,170 (22.29) | 71,149 (22.28) |  | 250,139 (24.6) | 250,130 (24.6) |  |
| South | 149,868 (46.94) | 149,880 (46.94) |  | 385,012 (37.8) | 385,108 (37.9) |  |
| West | 36,037 (11.29) | 36,037 (11.29) |  | 161,002 (15.8) | 161,085 (15.8) |  |
| Unknown | 10 (0.00) | 10 (0.00) |  | 2,980 (0.3) | 2,980 (0.3) |  |
| Race/ethnicity, n (%) ^b^ |  |  |  |  |  | 0.08 |
| American Indian/ Alaska Native | NA | NA |  | 7,510 (0.7) | 5,240 (0.5) |  |
| Asian | NA | NA |  | 16,819 (1.7) | 19,101 (1.9) |  |
| Black or African American | NA | NA |  | 90,997 (8.9) | 87,757 (8.6) |  |
| Hispanic/Latin American/Latinx | NA | NA |  | 26,460 (2.6) | 16,390 (1.6) |  |
| White | NA | NA |  | 845,042 (83.1) | 857,204 (84.3) |  |
| A race/ethnicity not listed | NA | NA |  | 13,898 (1.4) | 14,649 (1.4) |  |
| Not provided | NA | NA |  | 16,684 (1.6) | 17,069 (1.7) |  |
| Dual Medicare/Medicaid eligibility, n (%) ^b^ | NA | NA |  | 297,499 (29.2) | 267,943 (26.3) | 0.06 |
| Original reason for Medicare eligibility, n (%) ^b^ |  |  |  |  |  | 0.04 |
| Age | NA | NA |  | 860,430 (84.6) | 871,968 (85.7) |  |
| Disability | NA | NA |  | 150,916 (14.8) | 141,239 (13.9) |  |
| End-stage renal disease | NA | NA |  | 6,064 (0.6) | 4,203 (0.4) |  |
| Hospitalized on Time 0, n (%) | 4,046 (1.27) | 4,046 (1.27) | 0.00 | 166,154 (16.3) | 166,154 (16.3) | 0.00 |
| SNF/LTC ^b^ residence on Time 0, n (%) | NA | NA |  | 250,193 (24.6) | 250,193 (24.6) | 0.00 |
| Comorbidities, n (%) |  |  |  |  |  |  |
| Animal exposure/bites or rabies | 976 (0.31) | 1,017 (0.32) | 0.00 | 3,105 (0.3) | 3,204 (0.3) | 0.00 |
| Antiphospholipid syndrome | 160 (0.05) | 142 (0.04) | 0.00 | 829 (0.1) | 855 (0.1) | 0.00 |
| Autoimmune disorders | 12,677 (3.97) | 12,973 (4.06) | 0.00 | 93,071 (9.1) | 89,588 (8.8) | 0.01 |
| Brain lesions related to secondary narcolepsy | 30 (0.01) | 22 (0.01) | 0.00 | 519 (0.1) | 517 (0.1) | 0.00 |
| Cancer | 13,421 (4.20) | 14,271 (4.47) | 0.01 | 272,867 (26.8) | 283,933 (27.9) | 0.02 |
| Chronic lymphocytic leukemia | 213 (0.07) | 175 (0.05) | 0.00 | 6,995 (0.7) | 6,673 (0.7) | 0.00 |
| Chronic kidney disease or renal disease (other than end-stage renal disease) | 6,052 (1.90) | 5,101 (1.60) | 0.02 | 276,654 (27.2) | 262,576 (25.8) | 0.03 |
| End-stage renal disease ^b^ | NA | NA |  | 20,229 (2.0) | 14,209 (1.4) | 0.05 |
| Chronic liver disease | 10,763 (3.37) | 8,968 (2.81) | 0.03 | 80,781 (7.9) | 78,853 (7.8) | 0.01 |
| Chronic lung diseases | 32,616 (10.21) | 25,419 (7.96) | 0.08 | 293,426 (28.8) | 265,622 (26.1) | 0.06 |
| Dementia or other neurologic condition | 19,722 (6.18) | 16,421 (5.14) | 0.04 | 350,351 (34.4) | 308,057 (30.3) | 0.09 |
| Diabetes, Type 1 or 2 | 33,117 (10.37) | 25,491 (7.98) | 0.08 | 406,448 (39.9) | 364,235 (35.8) | 0.09 |
| Disseminated intravascular coagulation ^c^ | 24 (0.01) | 15 (0.00) | 0.00 | 324 (0.0) | 331 (0.0) | 0.00 |
| Heart conditions | 37,470 (11.74) | 31,315 (9.81) | 0.06 | 614,855 (60.4) | 586,102 (57.6) | 0.06 |
| Hemiplegia or paraplegia | 857 (0.27) | 815 (0.26) | 0.00 | 61,853 (6.1) | 61,794 (6.1) | 0.00 |
| Herpes simplex infection | 6,082 (1.90) | 4,738 (1.48) | 0.03 | 16,959 (1.7) | 15,509 (1.5) | 0.01 |
| Hypertension | 78,717 (24.65) | 69,065 (21.63) | 0.07 | 823,414 (80.9) | 800,051 (78.6) | 0.06 |
| Immunocompromised state | 37,672 (11.80) | 37,672 (11.80) | 0.00 | 438,954 (43.1) | 438,954 (43.1) | 0.00 |
| Infection associated with GBS | 17,961 (5.63) | 13,575 (4.25) | 0.06 | 51,279 (5.0) | 43,254 (4.3) | 0.04 |
| Viral infection associated with ITP | 1,290 (0.40) | 1,115 (0.35) | 0.01 | 3,702 (0.4) | 3,358 (0.3) | 0.01 |
| Infection associated with myocarditis/pericarditis | 16,753 (5.25) | 13,462 (4.22) | 0.05 | 111,923 (11.0) | 104,135 (10.2) | 0.02 |
| Inpatient surgery | 1,985 (0.62) | 2,925 (0.92) | 0.03 | 43,070 (4.2) | 53,725 (5.3) | 0.05 |
| Lipid abnormality | 73,480 (23.01) | 66,630 (20.87) | 0.05 | 733,659 (72.1) | 714,996 (70.3) | 0.04 |
| Mental health conditions | 73,913 (23.15) | 69,877 (21.88) | 0.03 | 441,954 (43.4) | 404,912 (39.8) | 0.07 |
| Nutritional deficiency | 2,088 (0.65) | 2,095 (0.66) | 0.00 | 80,039 (7.9) | 79,309 (7.8) | 0.00 |
| Obese or severely obese | 63,585 (19.91) | 49,693 (15.56) | 0.11 | 233,833 (23.0) | 209,074 (20.5) | 0.06 |
| Peripheral vascular disease | 4,393 (1.38) | 3,712 (1.16) | 0.02 | 318,384 (31.3) | 292,986 (28.8) | 0.05 |
| Pneumonia or lower respiratory tract infection | 41,355 (12.95) | 28,356 (8.88) | 0.13 | 237,118 (23.3) | 188,874 (18.6) | 0.12 |
| Pregnancy | 2,616 (0.82) | 2,288 (0.72) | 0.01 | NA | NA |  |
| Sepsis | 1,277 (0.40) | 942 (0.30) | 0.02 | 59,822 (5.9) | 58,347 (5.7) | 0.01 |
| Sickle cell disease or thalassemia | 750 (0.23) | 637 (0.20) | 0.01 | 2,611 (0.3) | 2,472 (0.2) | 0.00 |
| Smoking/nicotine dependency | 11,536 (3.61) | 14,735 (4.61) | 0.05 | 64,705 (6.4) | 80,909 (8.0) | 0.06 |
| Stroke or cerebrovascular disease | 4,730 (1.48) | 4,021 (1.26) | 0.02 | 228,802 (22.5) | 221,952 (21.8) | 0.02 |
| Thromboembolism | 3,531 (1.11) | 2,885 (0.90) | 0.02 | 77,562 (7.6) | 75,096 (7.4) | 0.01 |
| Thrombophilia | 1,096 (0.34) | 906 (0.28) | 0.01 | 11,107 (1.1) | 10,876 (1.1) | 0.00 |
| Trauma | 204 (0.06) | 259 (0.08) | 0.01 | 24,546 (2.4) | 29,888 (2.9) | 0.03 |
| Tuberculosis | 138 (0.04) | 115 (0.04) | 0.00 | 2,585 (0.3) | 2,299 (0.2) | 0.01 |
| Healthcare utilization, n (%) |  |  |  |  |  |  |
| Inpatient stays in previous 365 days |  |  | 0.05 |  |  | 0.02 |
| 0 | 305,018 (95.53) | 303,556 (95.07) |  | 729,890 (71.7) | 721,877 (71.0) |  |
| 1-2 | 13,162 (4.12) | 14,411 (4.51) |  | 204,101 (20.1) | 207,086 (20.4) |  |
| ≥ 3 | 1,120 (0.35) | 1,333 (0.42) |  | 83,419 (8.2) | 88,447 (8.7) |  |
| ED visits in previous 365 days |  |  | 0.13 |  |  | 0.06 |
| 0 | 255,758 (80.10) | 271,663 (85.08) |  | 572,454 (56.3) | 602,727 (59.2) |  |
| 1-2 | 54,798 (17.16) | 42,181 (13.21) |  | 282,442 (27.8) | 259,078 (25.5) |  |
| ≥ 3 | 8,744 (2.74) | 5,456 (1.71) |  | 162,514 (16.0) | 155,605 (15.3) |  |
| Outpatient provider visits ^d^ in previous 365 days |  |  | 0.19 |  |  | 0.17 |
| 0 | 2,076 (0.65) | 2,815 (0.88) |  | 93 (0.0) | 172 (0.0) |  |
| 1-2 | 56,880 (17.81) | 78,501 (24.59) |  | 21,024 (2.1) | 39,570 (3.9) |  |
| 3-5 | 76,244 (23.88) | 80,629 (25.25) |  | 51,668 (5.1) | 75,724 (7.4) |  |
| 6-10 | 76,772 (24.04) | 68,388 (21.42) |  | 118,531 (11.7) | 139,787 (13.7) |  |
| ≥ 11 | 107,328 (33.61) | 88,967 (27.86) |  | 826,094 (81.2) | 762,157 (74.9) |  |
| SNF/LTC stay ^e^ | 341 (0.11) | 158 (0.05) | 0.02 | 295,527 (29.0) | 286,390 (28.1) | 0.02 |
| Influenza vaccination in previous year | 90,018 (28.19) | 95,167 (29.80) | 0.04 | 570,066 (56.0) | 572,746 (56.3) | 0.01 |

ASD = absolute standardized difference; COVID‑19 = coronavirus disease 2019; ED = emergency department; GBS = Guillain-Barré syndrome; ITP = immune thrombocytopenia; LTC = long-term care; Q1 = first quartile, Q3 = third quartile; SD = standard deviation; SNF = skilled nursing facility; US = United States.

^a^ A unique individual may be included multiple times because of matching with replacement, and an individual may be included in both the exposure and comparator groups. This table counts each instance of an individual’s entry separately as distinct observations; the number of unique individuals is reported separately.

^b^ Only available in Medicare.

^c^ This serves as an outcome-specific exclusion criterion when the condition is evaluated as an outcome, but history of the condition serves as a covariate for analyses of other outcomes.

^d^ Unique days on which an outpatient visit occurred.

^e^ SNF and LTC both identified in Medicare; SNF identified in MarketScan.

sTable 7. Association of a COVID-19 Diagnosis With Adverse Events, by Follow-up Period, Cohort Design

| Outcome | Follow-up period | Exposure group | Outcome cases | Person-time (days) | sIPT-weighted HR (95% CI) |
| --- | --- | --- | --- | --- | --- |
| MarketScan | | | | | |
| Nonhemorrhagic stroke | Time 0 – end | COVID-19 | 106 | 26,599,730 | 1.45 (1.07-1.97) |
|  |  | Comparator | 75 | 26,621,239 | — |
|  | Time 0 | COVID-19 | 25 | 317,884 | 0.55 (0.33-0.90) |
|  |  | Comparator | 47 | 318,019 | — |
|  | Day 1 – end | COVID-19 | 81 | 26,599,705 | 3.04 (1.97-4.70) |
|  |  | Comparator | 28 | 26,621,192 | — |
| Hemorrhagic stroke | Time 0 – end | COVID-19 | 34 | 26,716,900 | 1.21 (0.67-2.20) |
|  |  | Comparator | 28 | 26,715,092 | — |
|  | Time 0 | COVID-19 | 4 | 319,035 | 0.23 (0.07-0.80) |
|  |  | Comparator | 16 | 319,026 | — |
|  | Day 1 – end | COVID-19 | 30 | 26,716,896 | 2.49 (1.27-4.89) |
|  |  | Comparator | 12 | 26,715,076 | — |
| Acute myocardial infarction | Time 0 – end | COVID-19 | 252 | 26,634,779 | 1.37 (1.12-1.68) |
|  |  | Comparator | 176 | 26,639,458 | — |
|  | Time 0 | COVID-19 | 57 | 318,271 | 0.57 (0.40-0.80) |
|  |  | Comparator | 101 | 318,319 | — |
|  | Day 1 – end | COVID-19 | 195 | 26,634,722 | 2.43 (1.84-3.23) |
|  |  | Comparator | 75 | 26,639,357 | — |
| Myocarditis/pericarditis | Time 0 – end | COVID-19 | 326 | 26,700,852 | 6.20 (4.46-8.62) |
|  |  | Comparator | 47 | 26,724,695 | — |
|  | Time 0 | COVID-19 | 111 | 319,096 | 21.04 (8.52-51.96) |
|  |  | Comparator | 5 | 319,163 | — |
|  | Day 1 – end | COVID-19 | 215 | 26,700,741 | 4.51 (3.15-6.45) |
|  |  | Comparator | 42 | 26,724,690 | — |
| Deep vein thrombosis | Time 0 – end | COVID-19 | 835 | 26,547,327 | 2.83 (2.45-3.28) |
|  |  | Comparator | 273 | 26,617,237 | — |
|  | Time 0 | COVID-19 | 123 | 317,909 | 3.09 (2.13-4.46) |
|  |  | Comparator | 38 | 318,133 | — |
|  | Day 1 – end | COVID-19 | 712 | 26,547,204 | 2.80 (2.39-3.27) |
|  |  | Comparator | 235 | 26,617,199 | — |
| Pulmonary embolism | Time 0 – end | COVID-19 | 1167 | 26,554,692 | 5.79 (4.93-6.80) |
|  |  | Comparator | 187 | 26,657,127 | — |
|  | Time 0 | COVID-19 | 324 | 318,207 | 5.58 (4.14-7.52) |
|  |  | Comparator | 56 | 318,524 | — |
|  | Day 1 – end | COVID-19 | 843 | 26,554,368 | 5.89 (4.86-7.12) |
|  |  | Comparator | 131 | 26,657,071 | — |
| Pulmonary embolism - inpatient only | Time 0 – end | COVID-19 | 442 | 26,616,138 | 4.95 (3.87-6.33) |
|  |  | Comparator | 86 | 26,665,160 | — |
|  | Time 0 | COVID-19 | 93 | 318,207 | 1.95 (1.36-2.81) |
|  |  | Comparator | 48 | 318,524 | — |
|  | Day 1 – end | COVID-19 | 349 | 26,616,045 | 8.65 (6.06-12.35) |
|  |  | Comparator | 38 | 26,665,112 | — |
| Disseminated intravascular coagulation | Time 0 – end | COVID-19 | 35 | 26,739,450 | 3.41 (1.67-6.93) |
|  |  | Comparator | 10 | 26,738,875 | — |
|  | Time 0 | COVID-19 | 11 | 319,264 | 1.64 (0.60-4.48) |
|  |  | Comparator | 6 | 319,279 | — |
|  | Day 1 – end | COVID-19 | 24 | 26,739,439 | 6.58 (2.27-19.06) |
|  |  | Comparator | 4 | 26,738,869 | — |
| Unusual-site thrombosis with thrombocytopenia syndrome | Time 0 – end | COVID-19 | 4 | 26,742,577 | 1.98 (0.35-11.05) |
|  |  | Comparator | 2 | 26,739,280 | — |
|  | Time 0 | COVID-19 | 0 | 319,276 | NE |
|  |  | Comparator | 0 | 319,277 | — |
|  | Day 1 – end | COVID-19 | 4 | 26,742,577 | 1.98 (0.35-11.05) |
|  |  | Comparator | 2 | 26,739,280 | — |
| Common-site thrombosis with thrombocytopenia syndrome | Time 0 – end | COVID-19 | 126 | 26,718,714 | 3.96 (2.67-5.88) |
|  |  | Comparator | 32 | 26,727,825 | — |
|  | Time 0 | COVID-19 | 32 | 319,147 | 2.76 (1.40-5.41) |
|  |  | Comparator | 12 | 319,172 | — |
|  | Day 1 – end | COVID-19 | 94 | 26,718,682 | 4.71 (2.89-7.68) |
|  |  | Comparator | 20 | 26,727,813 | — |
| Appendicitis | Time 0 – end | COVID-19 | 182 | 26,675,934 | 1.57 (1.23-1.99) |
|  |  | Comparator | 114 | 26,681,952 | — |
|  | Time 0 | COVID-19 | 70 | 318,640 | 3.70 (2.16-6.34) |
|  |  | Comparator | 19 | 318,665 | — |
|  | Day 1 – end | COVID-19 | 112 | 26,675,864 | 1.14 (0.86-1.50) |
|  |  | Comparator | 95 | 26,681,933 | — |
| Medicare | | | | | |
| Nonhemorrhagic stroke | Time 0 – end | COVID-19 | 5,569 | 74,436,513 | 0.89 (0.86-0.93) |
|  |  | Comparator | 6,362 | 79,256,964 | — |
|  | Time 0 | COVID-19 | 2,156 | 928,618 | 0.57 (0.54-0.61) |
|  |  | Comparator | 3,798 | 929,191 | — |
|  | Day 1 – end | COVID-19 | 3,315 | 73,357,490 | 1.35 (1.28-1.43) |
|  |  | Comparator | 2,525 | 78,143,257 | — |
| Hemorrhagic stroke | Time 0 – end | COVID-19 | 1,580 | 80,060,770 | 0.86 (0.8-0.92) |
|  |  | Comparator | 1,910 | 85,983,250 | — |
|  | Time 0 | COVID-19 | 561 | 1,003,495 | 0.52 (0.46-0.58) |
|  |  | Comparator | 1,119 | 1,002,866 | — |
|  | Day 1 – end | COVID-19 | 997 | 79,037,154 | 1.35 (1.22-1.50) |
|  |  | Comparator | 771 | 84,917,216 | — |
| Acute myocardial infarction | Time 0 – end | COVID-19 | 18,774 | 77,328,845 | 1.65 (1.61-1.70) |
|  |  | Comparator | 11,233 | 83,134,571 | — |
|  | Time 0 | COVID-19 | 9,219 | 975,854 | 1.41 (1.36-1.46) |
|  |  | Comparator | 6,396 | 975,129 | — |
|  | Day 1 – end | COVID-19 | 9,168 | 76,240,828 | 1.94 (1.86-2.01) |
|  |  | Comparator | 4,786 | 82,064,128 | — |
| Myocarditis/pericarditis | Time 0 – end | COVID-19 | 1,774 | 80,839,930 | 2.53 (2.29-2.79) |
|  |  | Comparator | 703 | 87,140,811 | — |
|  | Time 0 | COVID-19 | 669 | 1,015,725 | 4.08 (3.35-4.96) |
|  |  | Comparator | 159 | 1,015,798 | — |
|  | Day 1 – end | COVID-19 | 1,105 | 79,824,722 | 2.07 (1.84-2.32) |
|  |  | Comparator | 544 | 86,125,013 | — |
| Deep vein thrombosis | Time 0 – end | COVID-19 | 15,548 | 77,420,414 | 1.67 (1.63-1.72) |
|  |  | Comparator | 9,735 | 83,606,920 | — |
|  | Time 0 | COVID-19 | 3,676 | 981,125 | 1.64 (1.55-1.73) |
|  |  | Comparator | 2,278 | 982,606 | — |
|  | Day 1 – end | COVID-19 | 11,877 | 76,449,557 | 1.69 (1.63-1.74) |
|  |  | Comparator | 7,457 | 82,624,314 | — |
| Pulmonary embolism | Time 0 – end | COVID-19 | 14,274 | 78,835,990 | 2.47 (2.39-2.56) |
|  |  | Comparator | 6,021 | 85,281,541 | — |
|  | Time 0 | COVID-19 | 5,030 | 996,999 | 2.40 (2.27-2.53) |
|  |  | Comparator | 2,175 | 997,596 | — |
|  | Day 1 – end | COVID-19 | 9,244 | 77,850,826 | 2.53 (2.42-2.63) |
|  |  | Comparator | 3,846 | 84,283,945 | — |
| Pulmonary embolism - inpatient only | Time 0 – end | COVID-19 | 8,776 | 79,173,482 | 2.60 (2.49-2.72) |
|  |  | Comparator | 3,539 | 85,457,650 | — |
|  | Time 0 | COVID-19 | 3,650 | 996,999 | 2.08 (1.95-2.21) |
|  |  | Comparator | 1,830 | 997,596 | — |
|  | Day 1 – end | COVID-19 | 4,924 | 78,114,621 | 3.06 (2.88-3.26) |
|  |  | Comparator | 1,702 | 84,436,728 | — |
| Disseminated intravascular coagulation | Time 0 – end | COVID-19 | 1,269 | 81,006,596 | 2.73 (2.43-3.07) |
|  |  | Comparator | 480 | 87,276,904 | — |
|  | Time 0 | COVID-19 | 609 | 1,016,803 | 2.95 (2.49-3.49) |
|  |  | Comparator | 209 | 1,016,813 | — |
|  | Day 1 – end | COVID-19 | 655 | 79,988,878 | 2.54 (2.17-2.98) |
|  |  | Comparator | 271 | 86,258,848 | — |
| Unusual-site thrombosis with thrombocytopenia syndrome | Time 0 – end | COVID-19 | 83 | 81,057,517 | 0.82 (0.61-1.1) |
|  |  | Comparator | 109 | 87,298,860 | — |
|  | Time 0 | COVID-19 | 29 | 1,017,065 | 0.50 (0.32-0.79) |
|  |  | Comparator | 63 | 1,016,972 | — |
|  | Day 1 – end | COVID-19 | 55 | 80,040,985 | 1.26 (0.85-1.88) |
|  |  | Comparator | 47 | 86,281,346 | — |
| Common-site thrombosis with thrombocytopenia syndrome | Time 0 – end | COVID-19 | 7,393 | 80,060,181 | 2.19 (2.09-2.29) |
|  |  | Comparator | 3,453 | 86,171,433 | — |
|  | Time 0 | COVID-19 | 3,036 | 1,006,735 | 2.09 (1.95-2.23) |
|  |  | Comparator | 1,464 | 1,005,685 | — |
|  | Day 1 – end | COVID-19 | 4,556 | 79,033,139 | 2.35 (2.22-2.49) |
|  |  | Comparator | 2,006 | 85,128,856 | — |
| Appendicitis | Time 0 – end | COVID-19 | 310 | 80,967,062 | 0.73 (0.62-0.85) |
|  |  | Comparator | 442 | 87,170,269 | — |
|  | Time 0 | COVID-19 | 125 | 1,016,139 | 0.45 (0.36-0.56) |
|  |  | Comparator | 293 | 1,015,910 | — |
|  | Day 1 – end | COVID-19 | 177 | 79,950,149 | 1.22 (0.97-1.53) |
|  |  | Comparator | 149 | 86,150,387 | — |

CI = confidence interval; COVID‑19 = coronavirus disease 2019; ED = emergency department; HR = hazard ratio; sIPT = stabilized inverse probability of treatment.

— denotes the reference group.

Note: Privacy rules for Medicare require masking cell sizes of fewer than 11 individuals.

sTable 8. Selected Characteristics of Patients Hospitalized and Not Hospitalized at Time 0 by Exposure Group
A. MarketScan, Nonhemorrhagic Stroke Analysis Set

| Characteristic | Hospitalized at Time 0 | | | Not hospitalized at Time 0 | | |
| --- | --- | --- | --- | --- | --- | --- |
|  | COVID-19 diagnosis (exposure) group | Comparator group | ASD | COVID-19 diagnosis (exposure) group | Comparator group | ASD |
| Total selected individuals ^a^ (unique individuals) | 3,975 (3,975) | 3,876 (3,539) |  | 313,909 (313,909) | 314,143 (307,614) |  |
| Demographic characteristics |  |  |  |  |  |  |
| Age, years |  |  |  |  |  |  |
| Mean (SD) | 48.7 (12.5) | 48.5 (12.6) | 0.02 | 41.6 (13.6) | 41.7 (13.6) | 0.00 |
| Median (Q1, Q3) | 53 (40, 59) | 52 (39, 59) |  | 43 (30, 53) | 43 (30, 53) |  |
| Age groups, n (%) |  |  |  |  |  |  |
| 18-34 years | 711 (17.89) | 711 (18.34) |  | 105,033 (33.46) | 105,061 (33.44) |  |
| 35-49 years | 945 (23.77) | 921 (23.76) |  | 99,793 (31.79) | 99,862 (31.79) |  |
| 50-64 years | 2,319 (58.34) | 2,244 (57.89) |  | 109,083 (34.75) | 109,220 (34.77) |  |
| Sex, n (%) |  |  | 0.00 |  |  | 0.00 |
| Female | 2,007 (50.49) | 1,964 (50.67) |  | 176,208 (56.13) | 176,395 (56.15) |  |
| Male | 1,968 (49.51) | 1,912 (49.33) |  | 137,701 (43.87) | 137,748 (43.85) |  |
| US geographic region, n (%) |  |  | 0.03 |  |  | 0.14 |
| Northeast | 676 (17.01) | 657 (16.95) |  | 61,175 (19.49) | 61,291 (19.51) |  |
| North central | 759 (19.09) | 747 (19.27) |  | 70,155 (22.35) | 70,166 (22.34) |  |
| South | 2,058 (51.77) | 1,994 (51.44) |  | 147,124 (46.87) | 147,222 (46.86) |  |
| West | 482 (12.13) | 478 (12.33) |  | 35,445 (11.29) | 35,454 (11.29) |  |
| Unknown |  |  |  | 10 (0.00) | 10 (0.00) |  |
| Hospitalized on Time 0, n (%) | 3,975 (100.0) | 3,876 (100.0) | 0.00 | 0 (0.00) | 0 (0.00) | 0.00 |
| Comorbidities, n (%) |  |  |  |  |  |  |
| Autoimmune disorders | 163 (4.10) | 127 (3.28) | 0.04 | 12,349 (3.93) | 12,717 (4.05) | 0.01 |
| Cancer | 233 (5.86) | 381 (9.83) | 0.15 | 13,023 (4.15) | 13,745 (4.38) | 0.01 |
| Chronic kidney disease or renal disease (other than end-stage renal disease) | 323 (8.13) | 404 (10.42) | 0.08 | 5,470 (1.74) | 4,481 (1.43) | 0.03 |
| Chronic liver disease | 245 (6.16) | 422 (10.89) | 0.17 | 10,366 (3.30) | 8,406 (2.68) | 0.04 |
| Chronic lung diseases | 647 (16.28) | 625 (16.12) | 0.00 | 31,648 (10.08) | 24,508 (7.80) | 0.08 |
| Dementia or other neurologic condition | 266 (6.69) | 403 (10.40) | 0.13 | 18,936 (6.03) | 15,638 (4.98) | 0.05 |
| Diabetes, Type 1 or 2 | 1,127 (28.35) | 777 (20.05) | 0.19 | 31,537 (10.05) | 24,295 (7.73) | 0.08 |
| Heart conditions | 840 (21.13) | 1,214 (31.32) | 0.23 | 35,795 (11.40) | 29,346 (9.34) | 0.07 |
| Hemiplegia or paraplegia | 27 (0.68) | 43 (1.11) | 0.05 | 497 (0.16) | 482 (0.15) | 0.00 |
| Hypertension | 1,844 (46.39) | 1,803 (46.52) | 0.00 | 75,851 (24.16) | 66,327 (21.11) | 0.07 |
| Immunocompromised state | 582 (14.64) | 566 (14.60) | 0.00 | 36,614 (11.66) | 36,738 (11.69) | 0.00 |
| Lipid abnormality | 1,457 (36.65) | 1,360 (35.09) | 0.03 | 71,126 (22.66) | 64,437 (20.51) | 0.05 |
| Obese or severely obese | 1,332 (33.51) | 1,070 (27.61) | 0.13 | 61,693 (19.65) | 48,190 (15.34) | 0.11 |
| Peripheral vascular disease | 161 (4.05) | 225 (5.80) | 0.08 | 4,042 (1.29) | 3,317 (1.06) | 0.02 |
| Pneumonia or lower respiratory tract infection | 751 (18.89) | 629 (16.23) | 0.07 | 40,257 (12.82) | 27,500 (8.75) | 0.13 |
| Pregnancy | 41 (1.03) | 16 (0.41) | 0.07 | 2,571 (0.82) | 2,270 (0.72) | 0.01 |
| Sickle cell disease or thalassemia | 15 (0.38) | 17 (0.44) | 0.01 | 723 (0.23) | 613 (0.20) | 0.01 |
| Smoking/nicotine dependency | 192 (4.83) | 644 (16.62) | 0.39 | 11,172 (3.56) | 13,891 (4.42) | 0.04 |
| Stroke or cerebrovascular disease | 96 (2.42) | 165 (4.26) | 0.10 | 3,218 (1.03) | 2,575 (0.82) | 0.02 |
| Thrombophilia | 16 (0.40) | 38 (0.98) | 0.07 | 999 (0.32) | 801 (0.25) | 0.01 |
| Trauma | 13 (0.33) | 75 (1.93) | 0.15 | 177 (0.06) | 176 (0.06) | 0.00 |
| Healthcare utilization, n (%) |  |  |  |  |  |  |
| Inpatient stays in previous 365 days |  |  | 0.79 |  |  | 0.00 |
| 0 | 3,505 (88.18) | 2,117 (54.62) |  | 300,691 (95.79) | 300,702 (95.72) |  |
| 1-2 | 384 (9.66) | 1,287 (33.20) |  | 12,329 (3.93) | 12,719 (4.05) |  |
| ≥ 3 | 86 (2.16) | 472 (12.18) |  | 889 (0.28) | 722 (0.23) |  |
| ED visits in previous 365 days |  |  | 0.21 |  |  | 0.19 |
| 0 | 2,778 (69.89) | 2,366 (61.04) |  | 252,358 (80.39) | 268,624 (85.51) |  |
| 1-2 | 973 (24.48) | 1,107 (28.56) |  | 53,308 (16.98) | 40,630 (12.93) |  |
| ≥ 3 | 224 (5.64) | 403 (10.40) |  | 8,243 (2.63) | 4,889 (1.56) |  |
| Outpatient provider visits ^b^ in previous 365 days |  |  | 0.37 |  |  | 0.20 |
| 0 | 32 (0.81) | 56 (1.44) |  | 2,038 (0.65) | 2,756 (0.88) |  |
| 1-2 | 493 (12.40) | 252 (6.50) |  | 56,362 (17.95) | 78,216 (24.90) |  |
| 3-5 | 771 (19.40) | 409 (10.55) |  | 75,393 (24.02) | 80,112 (25.50) |  |
| 6-10 | 912 (22.94) | 729 (18.81) |  | 75,691 (24.11) | 67,467 (21.48) |  |
| ≥ 11 | 1,767 (44.45) | 2,430 (62.69) |  | 104,425 (33.27) | 85,592 (27.25) |  |
| SNF/LTC stay ^c^ | 25 (0.63) | 33 (0.85) | 0.03 | 229 (0.07) | 96 (0.03) | 0.02 |
| Influenza vaccination in previous year | 1,166 (29.33) | 1,258 (32.46) | 0.07 | 88,367 (28.15) | 93,442 (29.75) | 0.04 |

ASD = absolute standardized difference; COVID‑19 = coronavirus disease 2019; ED = emergency department; LTC = long-term care; Q1 = first quartile, Q3 = third quartile; SD = standard deviation; SNF = skilled nursing facility; US = United States.

^a^ A unique individual may be included multiple times because of matching with replacement, and an individual may be included in both the exposure and comparator groups. This table counts each instance of an individual’s entry separately as distinct observations; the number of unique individuals is reported separately.

^b^ Unique days on which an outpatient visit occurred.

^c^ SNF and LTC both identified in Medicare; SNF identified in MarketScan.

B. Medicare, Overall Cohort

| Characteristic | Hospitalized at Time 0 | | | Not hospitalized at Time 0 | | |
| --- | --- | --- | --- | --- | --- | --- |
|  | COVID-19 diagnosis (exposure) group | Comparator group | ASD | COVID-19 diagnosis (exposure) group | Comparator group | ASD |
| Total selected observations ^a^ (unique individuals) | 166,154 (166,154) | 166,154 (146,571) |  | 851,256 (851,256) | 851,256 (791,689) |  |
| Demographic characteristics |  |  |  |  |  |  |
| Age, years |  |  |  |  |  |  |
| Mean (SD) | 78.1(8.0) | 78.1(8.0) | 0.00 | 77.6 (8.7) | 77.6 (8.7) | 0.00 |
| Median (Q1, Q3) | 77 (72, 84) | 77 (72, 84) |  | 76 (70, 84) | 76 (70, 84) |  |
| Age groups, n (%) |  |  |  |  |  |  |
| 65-79 years | 97,335 (58.6) | 97,335 (58.6) |  | 532,374 (62.5) | 532,374 (62.5) |  |
| ≥ 80 years | 68,819 (41.4) | 68,819 (41.4) |  | 318,882 (37.5) | 318,882 (37.5) |  |
| Sex, n (%) |  |  | 0.00 |  |  | 0.00 |
| Female | 85,276 (51.3) | 85,276 (51.3) |  | 505,308 (59.4) | 505,308 (59.4) |  |
| Male | 80,878 (48.7) | 80,878 (48.7) |  | 345,948 (40.6) | 345,948 (40.6) |  |
| US geographic region, n (%) |  |  | 0.00 |  |  | 0.00 |
| Northeast | 36,702 (22.1) | 36,540 (22.0) |  | 181,575 (21.3) | 181,567 (21.3) |  |
| North central | 40,001 (24.1) | 39,958 (24.0) |  | 210,138 (24.7) | 210,172 (24.7) |  |
| South | 62,861 (37.8) | 63,003 (37.9) |  | 322,151 (37.8) | 322,105 (37.8) |  |
| West | 26,550 (16.0) | 26,613 (16.0) |  | 134,452 (15.8) | 134,472 (15.8) |  |
| Unknown | 40 (0.0) | 40 (0.0) |  | 2,940 (0.3) | 2,940 (0.3) |  |
| Race/ethnicity, n (%) ^b^ |  |  | 0.20 |  |  | 0.07 |
| American Indian/Alaska Native | 1,440 (0.9) | 783 (0.5) |  | 6,070 (0.7) | 4,457 (0.5) |  |
| Asian | 4,082 (2.5) | 3,033 (1.8) |  | 12,737 (1.5) | 16,068 (1.9) |  |
| Black or African American | 22,505 (13.5) | 17,331 (10.4) |  | 68,492 (8.0) | 70,426 (8.3) |  |
| Hispanic/Latin American/Latinx | 6,891 (4.1) | 2,847 (1.7) |  | 19,569 (2.3) | 13,543 (1.6) |  |
| White | 126,070 (75.9) | 137,362 (82.7) |  | 718,972 (84.5) | 719,842 (84.6) |  |
| A race/ethnicity not listed | 2,861 (1.7) | 2,437 (1.5) |  | 11,037 (1.3) | 12,212 (1.4) |  |
| Not provided | 2,305 (1.4) | 2,361 (1.4) |  | 14,379 (1.7) | 14,708 (1.7) |  |
| Dual Medicare/Medicaid eligibility, n (%) ^b^ | 44,237 (26.6) | 32,416 (19.5) | 0.17 | 253,262 (29.8) | 235,527 (27.7) | 0.05 |
| Original reason for Medicare eligibility, n (%) ^b^ |  |  | 0.05 |  |  | 0.03 |
| Age | 135,171 (81.4) | 138,126 (83.1) |  | 725,259 (85.2) | 733,842 (86.2) |  |
| Disability | 28,674 (17.3) | 26,398 (15.9) |  | 122,242 (14.4) | 114,841 (13.5) |  |
| End-stage renal disease | 2,309 (1.4) | 1,630 (1.0) |  | 3,755 (0.4) | 2,573 (0.3) |  |
| Hospitalized on Time 0, n (%) | 166,154 (100.0) | 166,154 (100.0) | 0.00 | 0 (0.0) | 0 (0.0) | 0.00 |
| SNF/LTC ^b^ residence on Time 0, n (%) | 10,656 (6.4) | 10,656 (6.4) | 0.00 | 239,537 (28.1) | 239,537 (28.1) | 0.00 |
| Comorbidities, n (%) |  |  |  |  |  |  |
| Animal exposure/bites or rabies | 495 (0.3) | 698 (0.4) | 0.02 | 2,610 (0.3) | 2,506 (0.3) | 0.00 |
| Antiphospholipid syndrome | 171 (0.1) | 244 (0.1) | 0.01 | 658 (0.1) | 611 (0.1) | 0.00 |
| Autoimmune disorders | 16,815 (10.1) | 17,383 (10.5) | 0.01 | 76,256 (9.0) | 72,205 (8.5) | 0.02 |
| Brain lesions related to secondary narcolepsy | 103 (0.1) | 138 (0.1) | 0.01 | 416 (0.0) | 379 (0.0) | 0.00 |
| Cancer | 49,656 (29.9) | 58,375 (35.1) | 0.11 | 223,211 (26.2) | 225,558 (26.5) | 0.01 |
| Chronic lymphocytic leukemia | 1,809 (1.1) | 1,745 (1.1) | 0.00 | 5,186 (0.6) | 4,928 (0.6) | 0.00 |
| Chronic kidney disease or renal disease (other than end-stage renal disease) | 62,434 (37.6) | 71,844 (43.2) | 0.12 | 214,220 (25.2) | 190,732 (22.4) | 0.06 |
| End-stage renal disease ^b^ | 7,103 (4.3) | 5,715 (3.4) | 0.04 | 13,126 (1.5) | 8,494 (1.0) | 0.05 |
| Chronic liver disease | 17,134 (10.3) | 23,454 (14.1) | 0.12 | 63,647 (7.5) | 55,399 (6.5) | 0.04 |
| Chronic lung diseases | 59,580 (35.9) | 62,585 (37.7) | 0.04 | 233,846 (27.5) | 203,037 (23.9) | 0.08 |
| Dementia or other neurologic condition | 53,409 (32.1) | 52,600 (31.7) | 0.01 | 296,942 (34.9) | 255,457 (30.0) | 0.10 |
| Diabetes, Type 1 or 2 | 81,608 (49.1) | 74,701 (45.0) | 0.08 | 324,840 (38.2) | 289,534 (34.0) | 0.09 |
| Disseminated intravascular coagulation ^c^ | 69 (0.0) | 170 (0.1) | 0.02 | 255 (0.0) | 161 (0.0) | 0.01 |
| Heart conditions | 112,906 (68.0) | 128,981 (77.6) | 0.22 | 501,949 (59.0) | 457,121 (53.7) | 0.11 |
| Hemiplegia or paraplegia | 10,793 (6.5) | 13,834 (8.3) | 0.07 | 51,060 (6.0) | 47,960 (5.6) | 0.02 |
| Herpes simplex infection | 2,684 (1.6) | 3,031 (1.8) | 0.02 | 14,275 (1.7) | 12,478 (1.5) | 0.02 |
| Hypertension | 142,424 (85.7) | 145,476 (87.6) | 0.05 | 680,990 (80.0) | 654,575 (76.9) | 0.08 |
| Immunocompromised state | 82,503 (49.7) | 82,503 (49.7) | 0.00 | 356,451 (41.9) | 356,451 (41.9) | 0.00 |
| Infection associated with GBS | 9,714 (5.8) | 10,224 (6.2) | 0.01 | 41,565 (4.9) | 33,030 (3.9) | 0.05 |
| Viral infection associated with ITP | 914 (0.6) | 834 (0.5) | 0.01 | 2,788 (0.3) | 2,524 (0.3) | 0.01 |
| Infection associated with myocarditis/pericarditis | 22,001 (13.2) | 27,322 (16.4) | 0.09 | 89,922 (10.6) | 76,813 (9.0) | 0.05 |
| Inpatient surgery | 10,819 (6.5) | 25,685 (15.5) | 0.29 | 32,251 (3.8) | 28,040 (3.3) | 0.03 |
| Lipid abnormality | 126,585 (76.2) | 131,236 (79.0) | 0.07 | 607,074 (71.3) | 583,760 (68.6) | 0.06 |
| Mental health conditions | 67,856 (40.8) | 73,387 (44.2) | 0.07 | 374,098 (43.9) | 331,525 (38.9) | 0.10 |
| Nutritional deficiency | 15,775 (9.5) | 22,827 (13.7) | 0.13 | 64,264 (7.5) | 56,482 (6.6) | 0.04 |
| Obese or severely obese | 48,362 (29.1) | 47,187 (28.4) | 0.02 | 185,471 (21.8) | 161,887 (19.0) | 0.07 |
| Peripheral vascular disease | 56,842 (34.2) | 63,496 (38.2) | 0.08 | 261,542 (30.7) | 229,490 (27.0) | 0.08 |
| Pneumonia or lower respiratory tract infection | 46,921 (28.2) | 47,039 (28.3) | 0.00 | 190,197 (22.3) | 141,835 (16.7) | 0.14 |
| Sepsis | 13,956 (8.4) | 20,391 (12.3) | 0.13 | 45,866 (5.4) | 37,956 (4.5) | 0.04 |
| Sickle cell disease or thalassemia | 552 (0.3) | 628 (0.4) | 0.01 | 2,059 (0.2) | 1,844 (0.2) | 0.01 |
| Smoking/nicotine dependency | 13,359 (8.0) | 21,867 (13.2) | 0.17 | 51,346 (6.0) | 59,042 (6.9) | 0.04 |
| Stroke or cerebrovascular disease | 42,407 (25.5) | 53,240 (32.0) | 0.14 | 186,395 (21.9) | 168,712 (19.8) | 0.05 |
| Thromboembolism | 17,129 (10.3) | 22,397 (13.5) | 0.10 | 60,433 (7.1) | 52,699 (6.2) | 0.04 |
| Thrombophilia | 2,388 (1.4) | 3,344 (2.0) | 0.04 | 8,719 (1.0) | 7,532 (0.9) | 0.01 |
| Trauma | 4,956 (3.0) | 11,989 (7.2) | 0.19 | 19,590 (2.3) | 17,899 (2.1) | 0.01 |
| Tuberculosis | 609 (0.4) | 671 (0.4) | 0.01 | 1,976 (0.2) | 1,628 (0.2) | 0.01 |
| Healthcare utilization, n (%) |  |  |  |  |  |  |
| Inpatient stays in previous 365 days |  |  | 0.46 |  |  | 0.08 |
| 0 | 102,446 (61.7) | 65,135 (39.2) |  | 627,444 (73.7) | 656,742 (77.1) |  |
| 1-2 | 41,039 (24.7) | 63,162 (38.0) |  | 163,062 (19.2) | 143,924 (16.9) |  |
| ≥ 3 | 22,669 (13.6) | 37,857 (22.8) |  | 60,750 (7.1) | 50,590 (5.9) |  |
| ED visits in previous 365 days |  |  | 0.32 |  |  | 0.13 |
| 0 | 73,774 (44.4) | 50,116 (30.2) |  | 498,680 (58.6) | 552,611 (64.9) |  |
| 1-2 | 52,748 (31.7) | 57,407 (34.6) |  | 229,694 (27.0) | 201,671 (23.7) |  |
| ≥ 3 | 39,632 (23.9) | 58,631 (35.3) |  | 122,882 (14.4) | 96,974 (11.4) |  |
| Outpatient provider visits ^d^ in previous 365 days |  |  | 0.13 |  |  | 0.22 |
| 0 | 38 (0.0) | 53 (0.0) |  | 55 (0.0) | 119 (0.0) |  |
| 1-2 | 4,185 (2.5) | 3,284 (2.0) |  | 16,839 (2.0) | 36,286 (4.3) |  |
| 3-5 | 8,644 (5.2) | 6,020 (3.6) |  | 43,024 (5.1) | 69,704 (8.2) |  |
| 6-10 | 18,130 (10.9) | 13,560 (8.2) |  | 100,401 (11.8) | 126,227 (14.8) |  |
| ≥ 11 | 135,157 (81.3) | 143,237 (86.2) |  | 690,937 (81.2) | 618,920 (72.7) |  |
| SNF/LTC stay ^e^ | 28,873 (17.4) | 30,503 (18.4) | 0.03 | 266,654 (31.3) | 255,887 (30.1) | 0.03 |
| Influenza vaccination in previous year | 93,215 (56.1) | 97,178 (58.5) | 0.05 | 476,851 (56.0) | 475,568 (55.9) | 0.00 |

ASD = absolute standardized difference; COVID‑19 = coronavirus disease 2019; ED = emergency department; GBS = Guillain-Barré syndrome; ITP = immune thrombocytopenia; LTC = long-term care; Q1 = first quartile, Q3 = third quartile; SD = standard deviation; SNF = skilled nursing facility; US = United States.

^a^ A unique individual may be included multiple times because of matching with replacement, and an individual may be included in both the exposure and comparator groups. This table counts each instance of an individual’s entry separately as distinct observations; the number of unique individuals is reported separately.

^b^ Only available in Medicare.

^c^ This serves as an outcome-specific exclusion criterion when the condition is evaluated as an outcome, but history of the condition serves as a covariate for analyses of other outcomes.

^d^ Unique days on which an outpatient visit occurred.

^e^ SNF and LTC both identified in Medicare; only SNF identified in MarketScan.

Note: Results presented in the overall, descriptive cohort before subdividing into adverse event-specific analysis sets.

sTable 9. Association of a COVID-19 Diagnosis With Adverse Events, by Hospitalization Status, Cohort Design

| Outcome | Analysis | sIPT-weighted HR (95% CI) | |
| --- | --- | --- | --- |
|  |  | MarketScan | Medicare |
| Nonhemorrhagic stroke | Overall (Time 0 – end) | 1.45 (1.07-1.97) | 0.89 (0.86-0.93) |
|  | Hospitalized at Time 0 | 0.50 (0.30-0.84) | 0.54 (0.51-0.57) |
|  | Not hospitalized at Time 0 | 2.84 (1.83-4.40) | 1.67 (1.56-1.78) |
| Hemorrhagic stroke | Overall (Time 0 – end) | 1.21 (0.67-2.20) | 0.86 (0.80-0.92) |
|  | Hospitalized at Time 0 | 0.42 (0.15-1.14) | 0.54 (0.49-0.59) |
|  | Not hospitalized at Time 0 | 2.55 (1.18-5.50) | 1.79 (1.58-2.04) |
| Acute Myocardial Infarction | Overall (Time 0 – end) | 1.37 (1.12-1.68) | 1.65 (1.61-1.70) |
|  | Hospitalized at Time 0 | 0.51 (0.35-0.73) | 1.22 (1.18-1.26) |
|  | Not hospitalized at Time 0 | 2.63 (1.97-3.52) | 2.46 (2.35-2.58) |
| Myocarditis/Pericarditis | Overall (Time 0 – end) | 6.20 (4.46-8.62) | 2.53 (2.29-2.79) |
|  | Hospitalized at Time 0 | 1.40 (0.42-4.70) | 2.03 (1.77-2.33) |
|  | Not hospitalized at Time 0 | 7.37 (5.25-10.35) | 3.47 (2.95-4.07) |
| Deep Vein Thrombosis | Overall (Time 0 – end) | 2.83 (2.45-3.28) | 1.67 (1.63-1.72) |
|  | Hospitalized at Time 0 | 1.23 (0.89-1.71) | 1.28 (1.23-1.33) |
|  | Not hospitalized at Time 0 | 3.60 (3.04-4.26) | 2.17 (2.09-2.26) |
| Pulmonary Embolism – any setting | Overall (Time 0 – end) | 5.79 (4.93-6.80) | 2.47 (2.39-2.56) |
|  | Hospitalized at Time 0 | 1.87 (1.38-2.54) | 1.75 (1.68-1.83) |
|  | Not hospitalized at Time 0 | 8.32 (6.75-10.25) | 3.70 (3.51-3.91) |
| Pulmonary Embolism - inpatient only | Overall (Time 0 – end) | 4.95 (3.87-6.33) | 2.60 (2.49-2.72) |
|  | Hospitalized at Time 0 | 1.64 (1.13-2.38) | 1.74 (1.65-1.84) |
|  | Not hospitalized at Time 0 | 10.53 (7.00-15.84) | 4.67 (4.32-5.06) |
| Disseminated Intravascular Coagulation | Overall (Time 0 – end) | 3.41 (1.67-6.93) | 2.73 (2.43-3.07) |
|  | Hospitalized at Time 0 | 0.86 (0.28-2.68) | 2.25 (1.96-2.58) |
|  | Not hospitalized at Time 0 | 12.80 (3.00-54.51) | 4.44 (3.55-5.55) |
| Unusual-site thrombosis with thrombocytopenia syndrome | Overall (Time 0 – end) | 1.98 (0.35-11.05) | 0.82 (0.61-1.10) |
|  | Hospitalized at Time 0 | NE | 0.58 (0.40-0.86) |
|  | Not hospitalized at Time 0 | 3.27 (0.36-29.42) | 1.83 (1.05-3.19) |
| Common-site thrombosis with thrombocytopenia syndrome | Overall (Time 0 – end) | 3.96 (2.67-5.88) | 2.19 (2.09-2.29) |
|  | Hospitalized at Time 0 | 2.01 (1.06-3.82) | 1.81 (1.71-1.91) |
|  | Not hospitalized at Time 0 | 9.02 (4.65-17.47) | 3.27 (3.01-3.54) |
| Appendicitis | Overall (Time 0 – end) | 1.57 (1.23-1.99) | 0.73 (0.62-0.85) |
|  | Hospitalized at Time 0 | 0.64 (0.30-1.35) | 0.36 (0.29-0.44) |
|  | Not hospitalized at Time 0 | 1.76 (1.36-2.28) | 1.55 (1.21-1.99) |

CI = confidence interval; COVID‑19 = coronavirus disease 2019; ED = emergency department; HR = hazard ratio; sIPT = stabilized inverse probability of treatment.

— denotes the reference group.

sFigure 1. Balance of Covariate Distributions Among Individuals With a COVID-19 Diagnosis and Comparator Individuals Without a COVID-19 Diagnosis, Cohort Design, Before and After Stabilized Inverse Probability of Treatment Weighting
A. Appendicitis, MarketScan

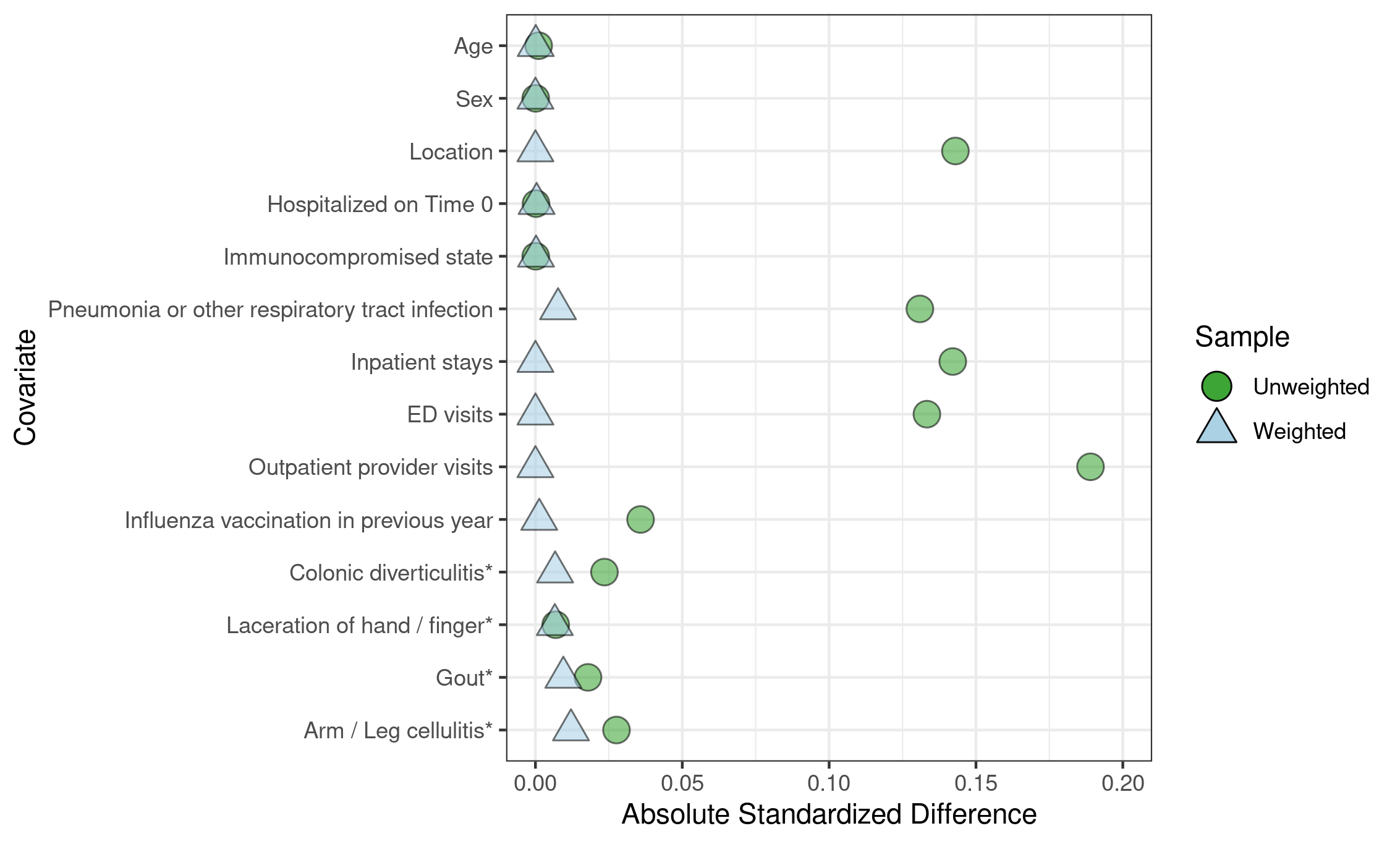

B. Nonhemorrhagic Stroke, MarketScan

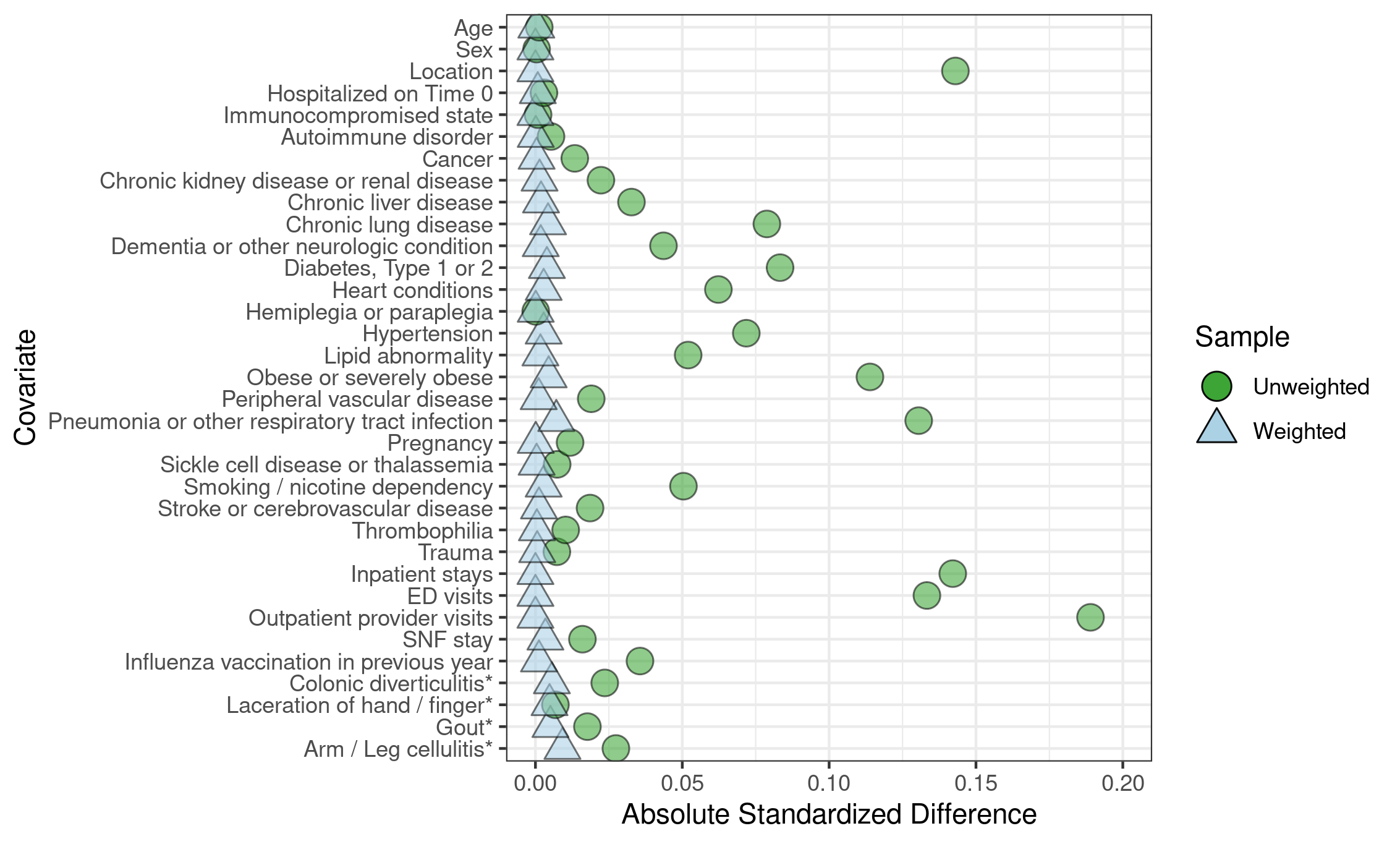

C. Hemorrhagic Stroke, MarketScan

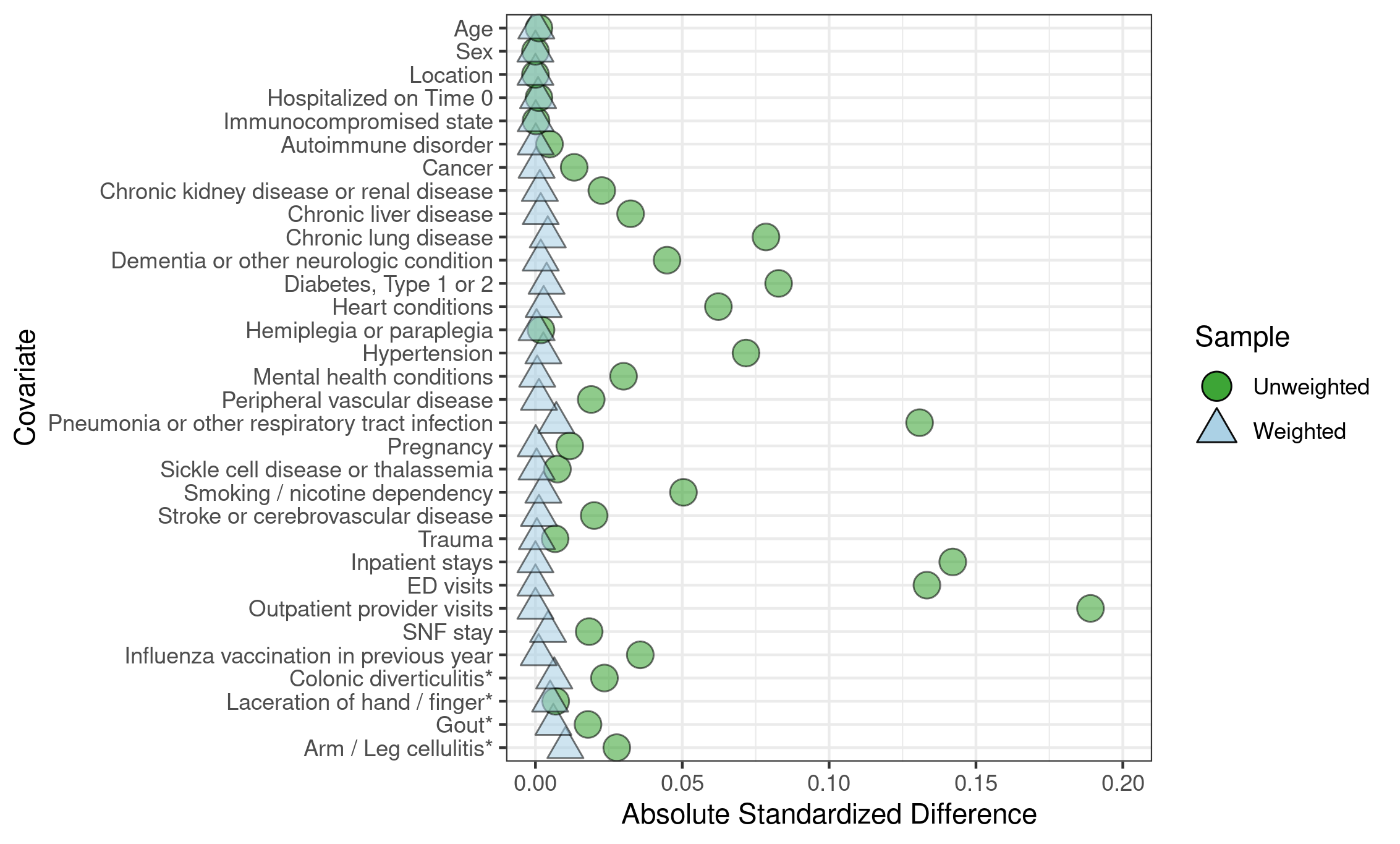

D. Acute Myocardial Infarction, MarketScan

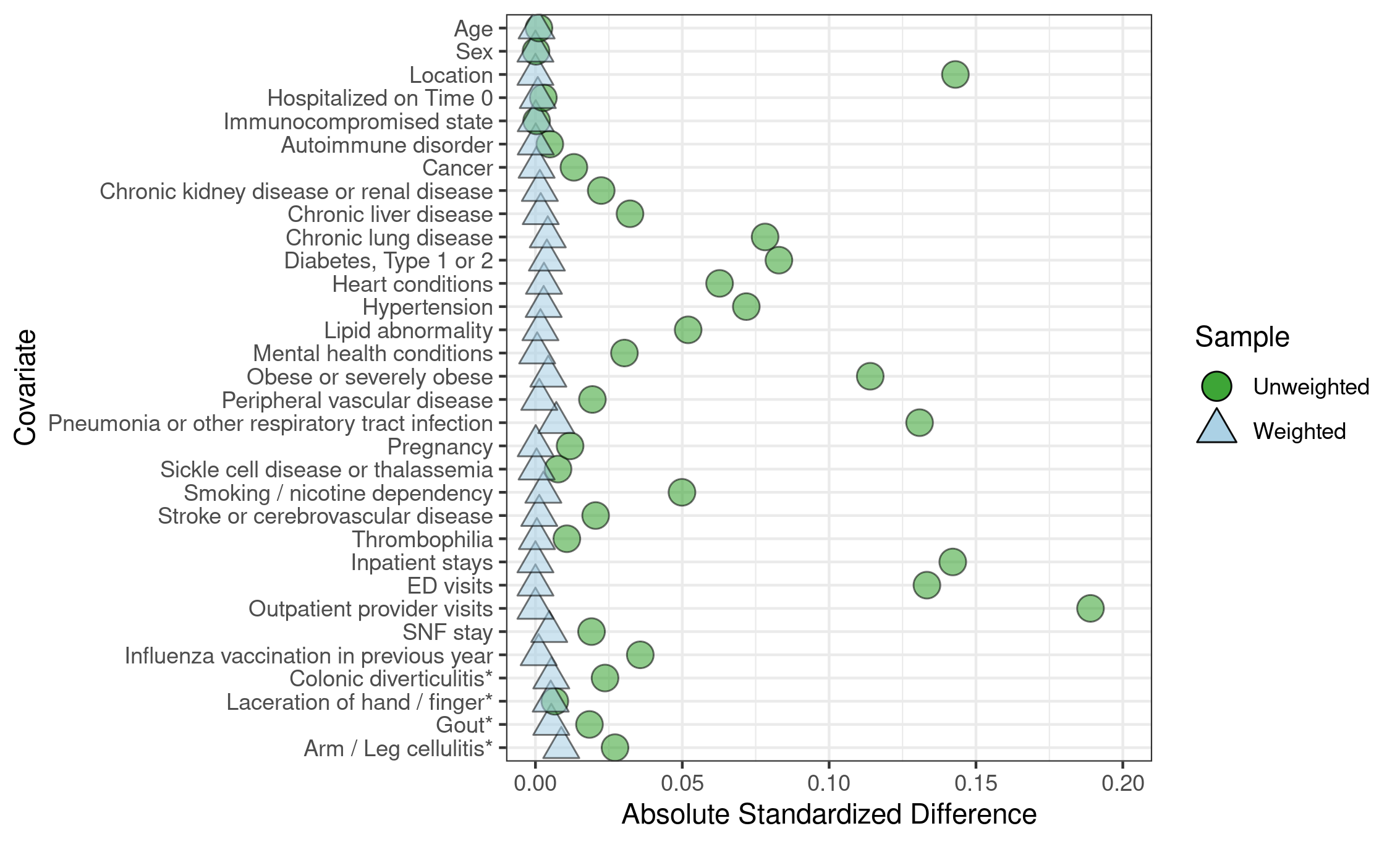

E. Myocarditis/Pericarditis, MarketScan

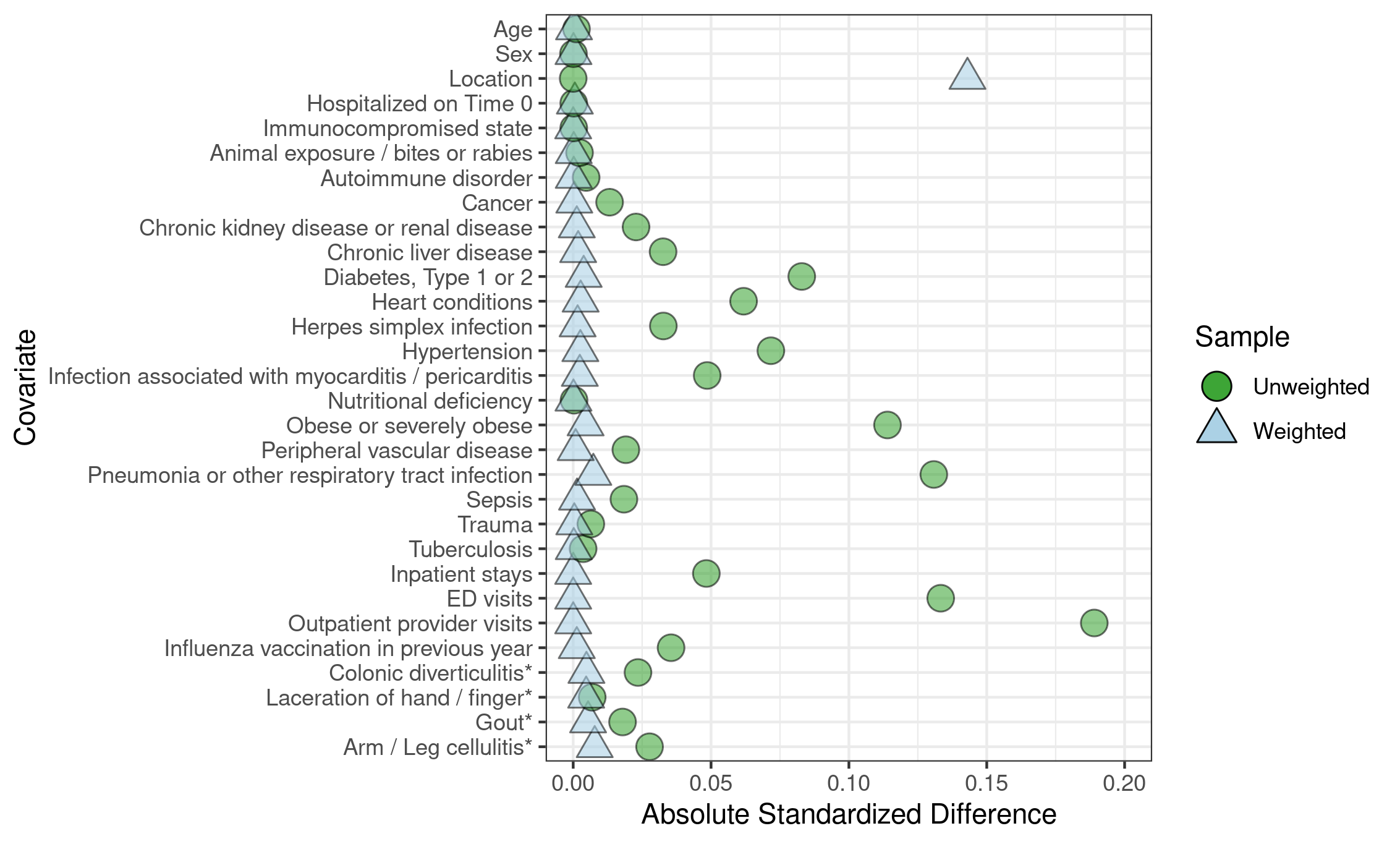

F. Deep Vein Thrombosis, MarketScan

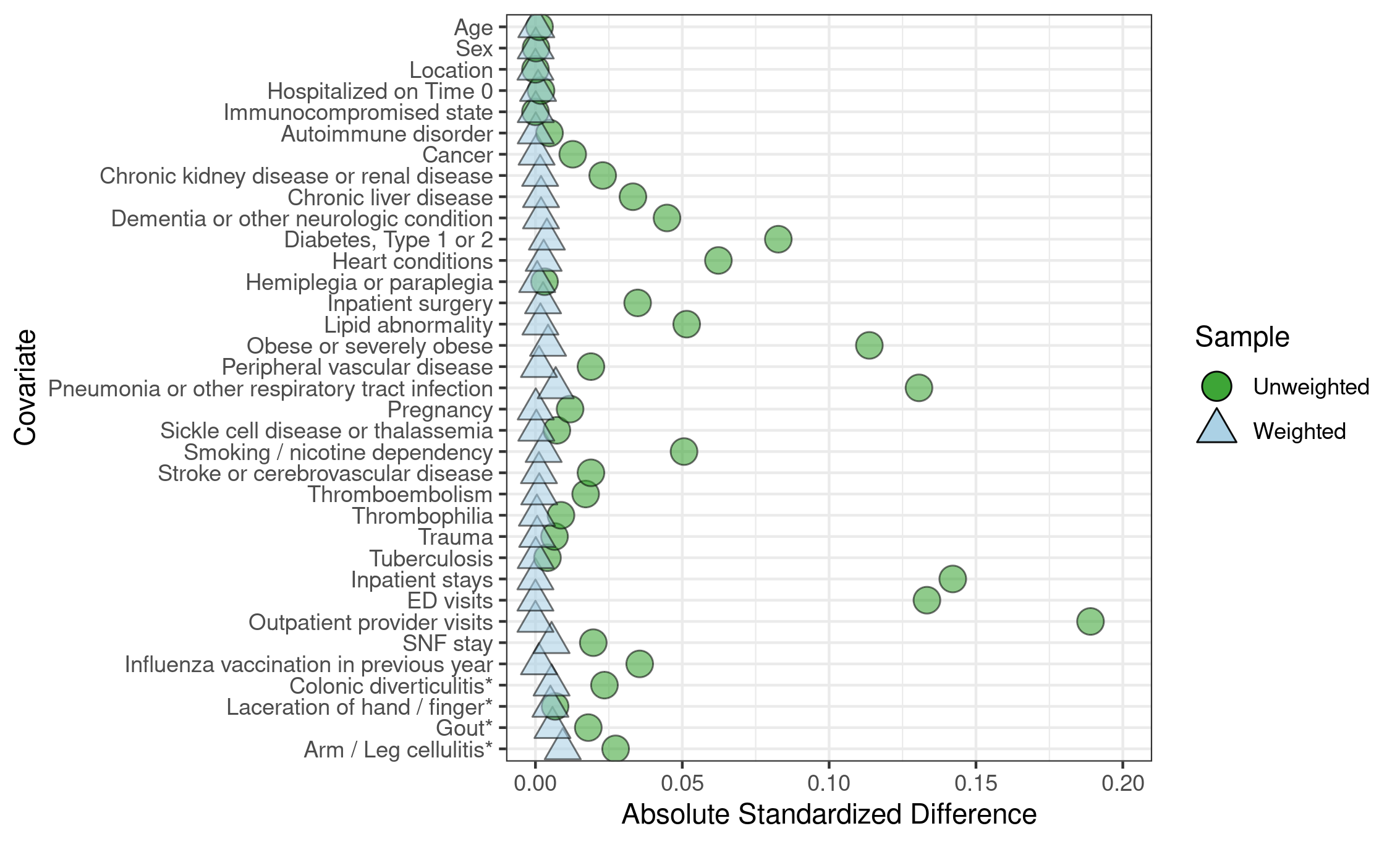

G. Pulmonary Embolism, MarketScan

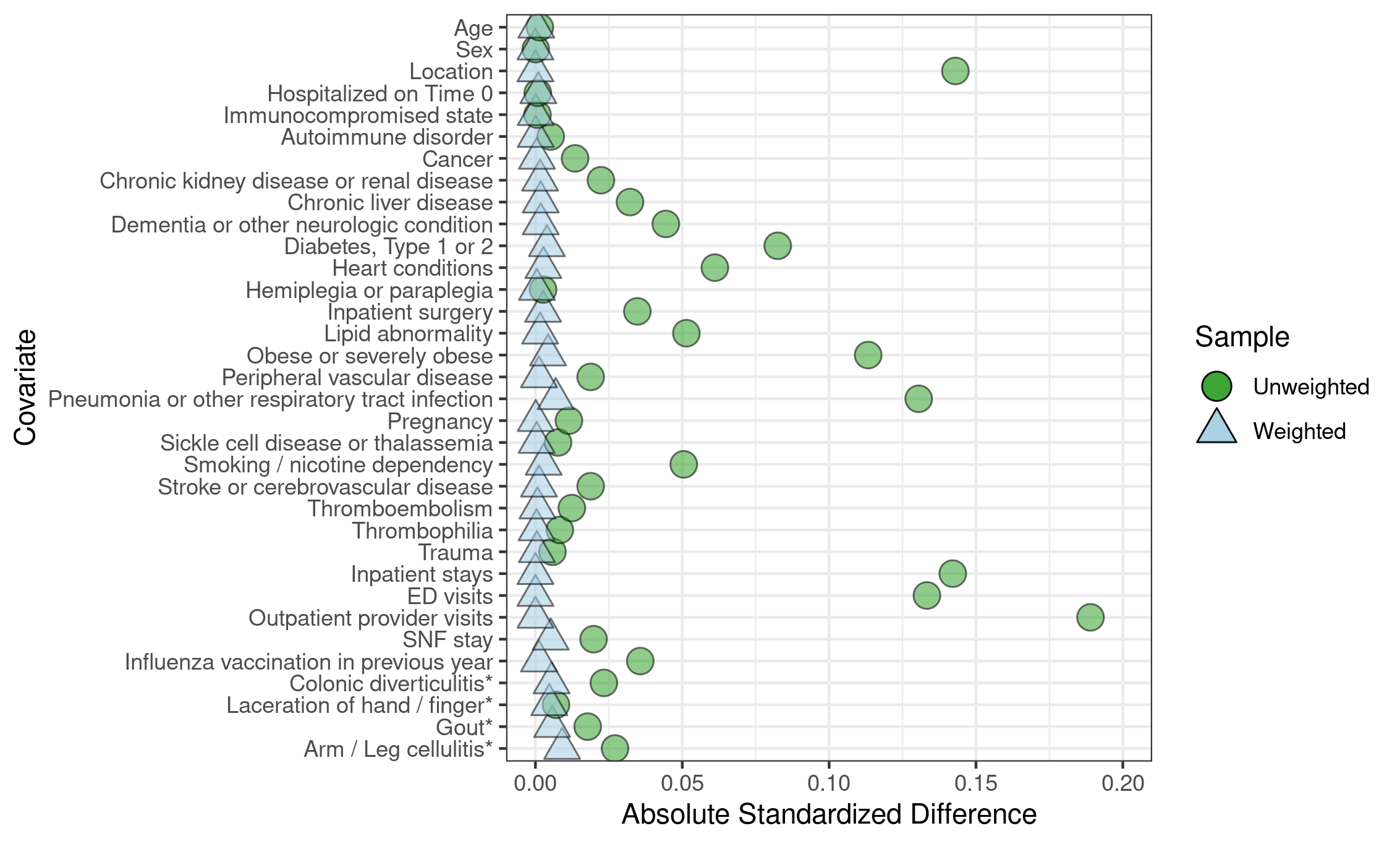

H. Disseminated Intravascular Coagulation, MarketScan

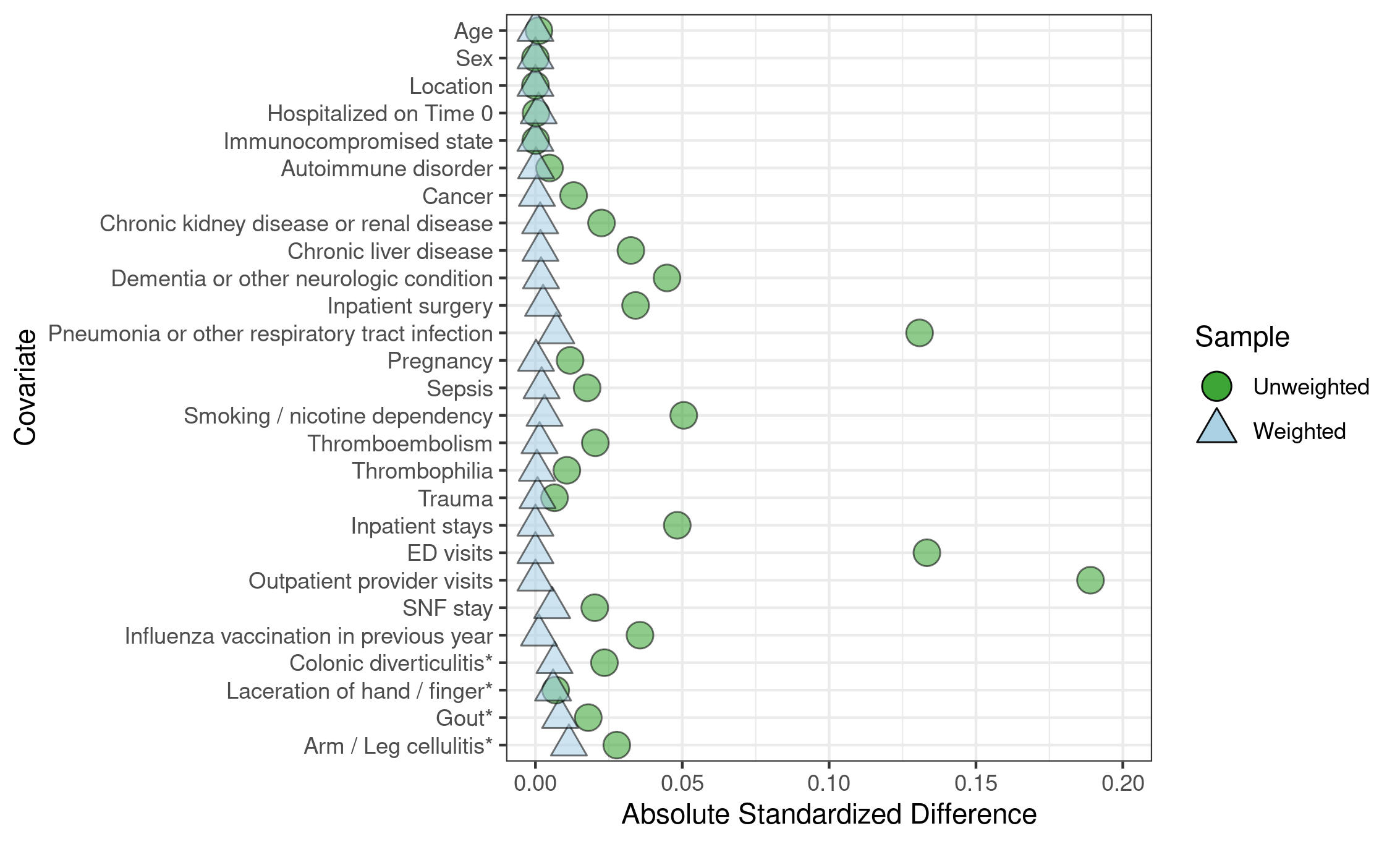

I. Unusual-Site Thrombosis With Thrombocytopenia Syndrome, MarketScan

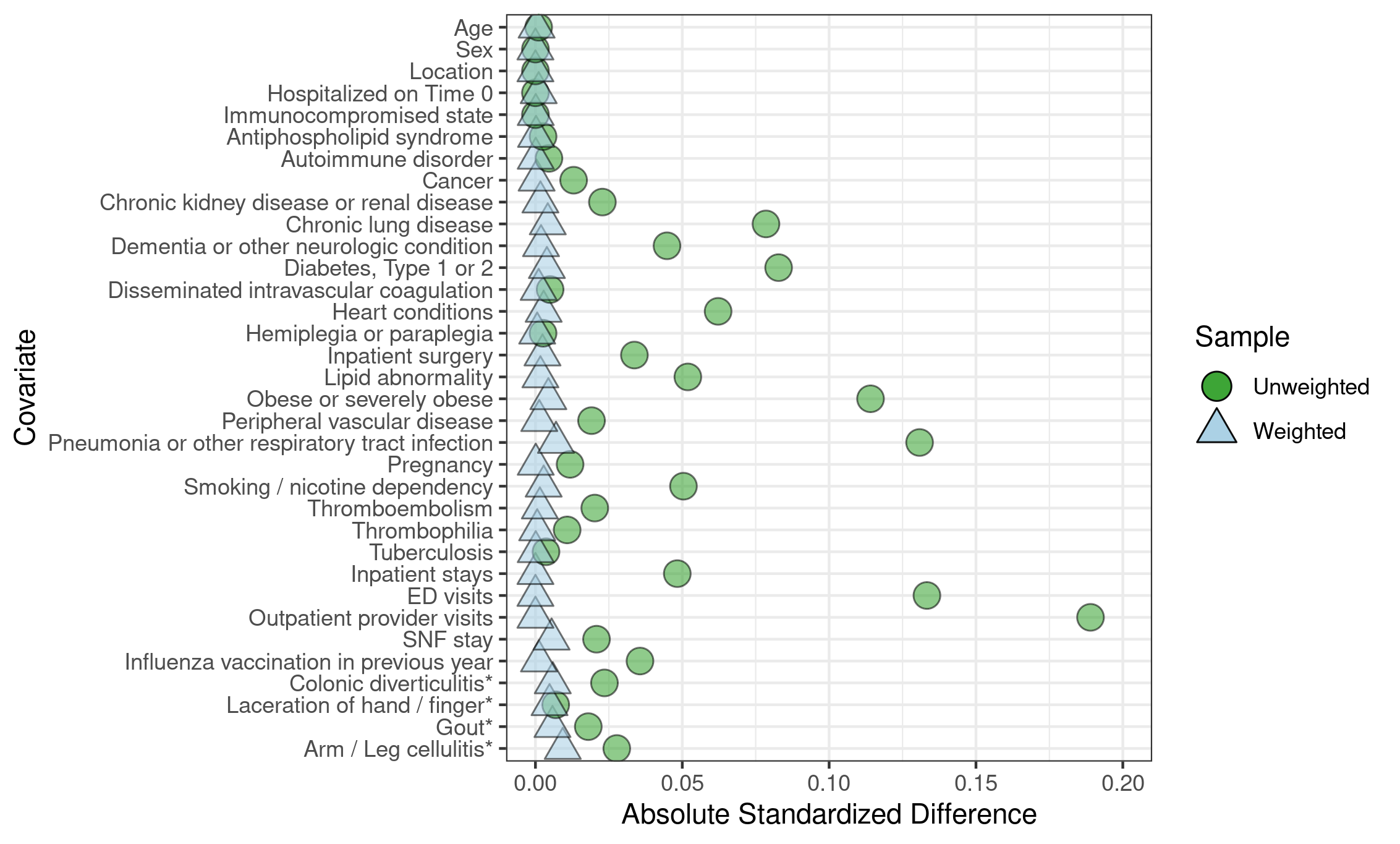

J. Common-Site Thrombosis With Thrombocytopenia Syndrome, MarketScan

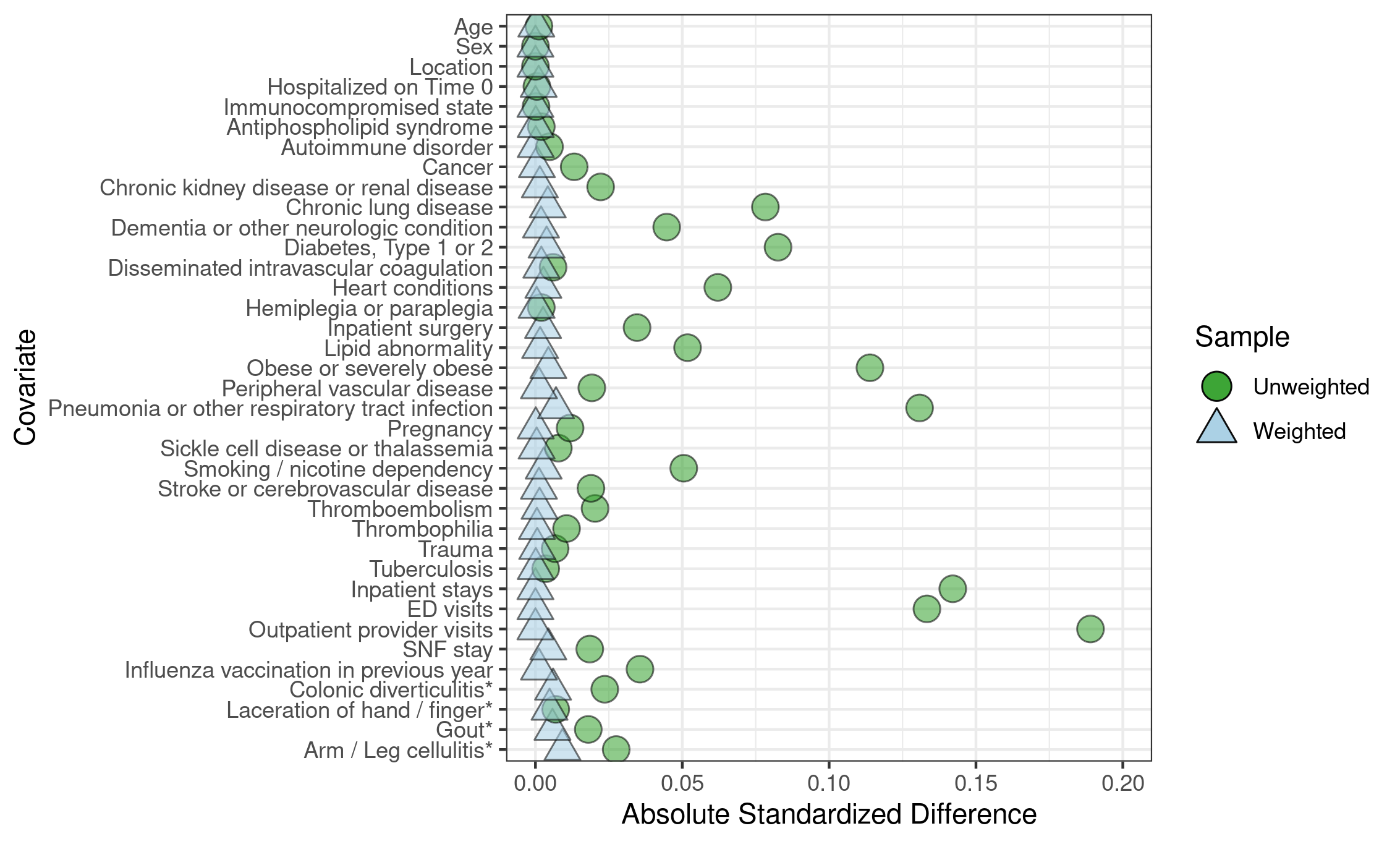

K. Appendicitis, Medicare

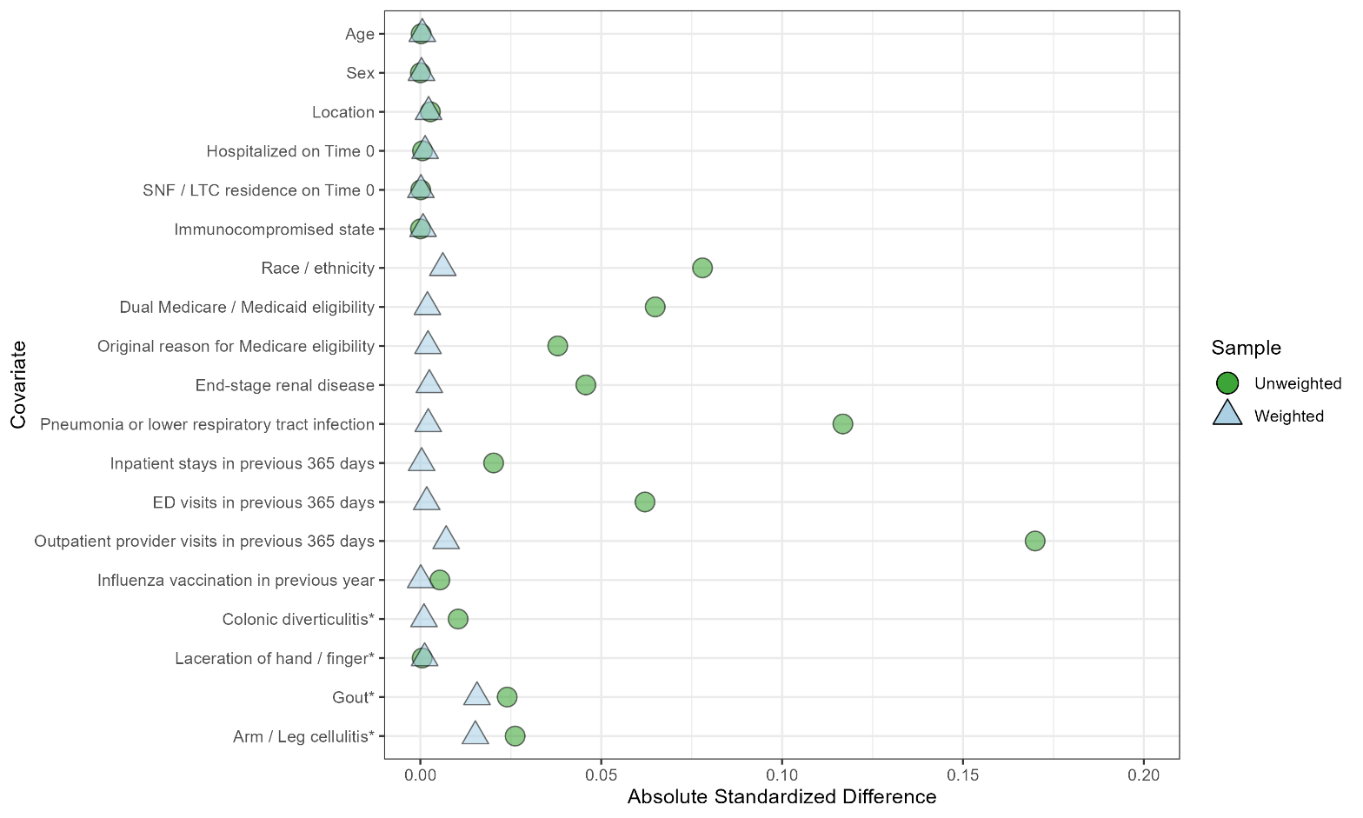

L. Nonhemorrhagic Stroke, Medicare

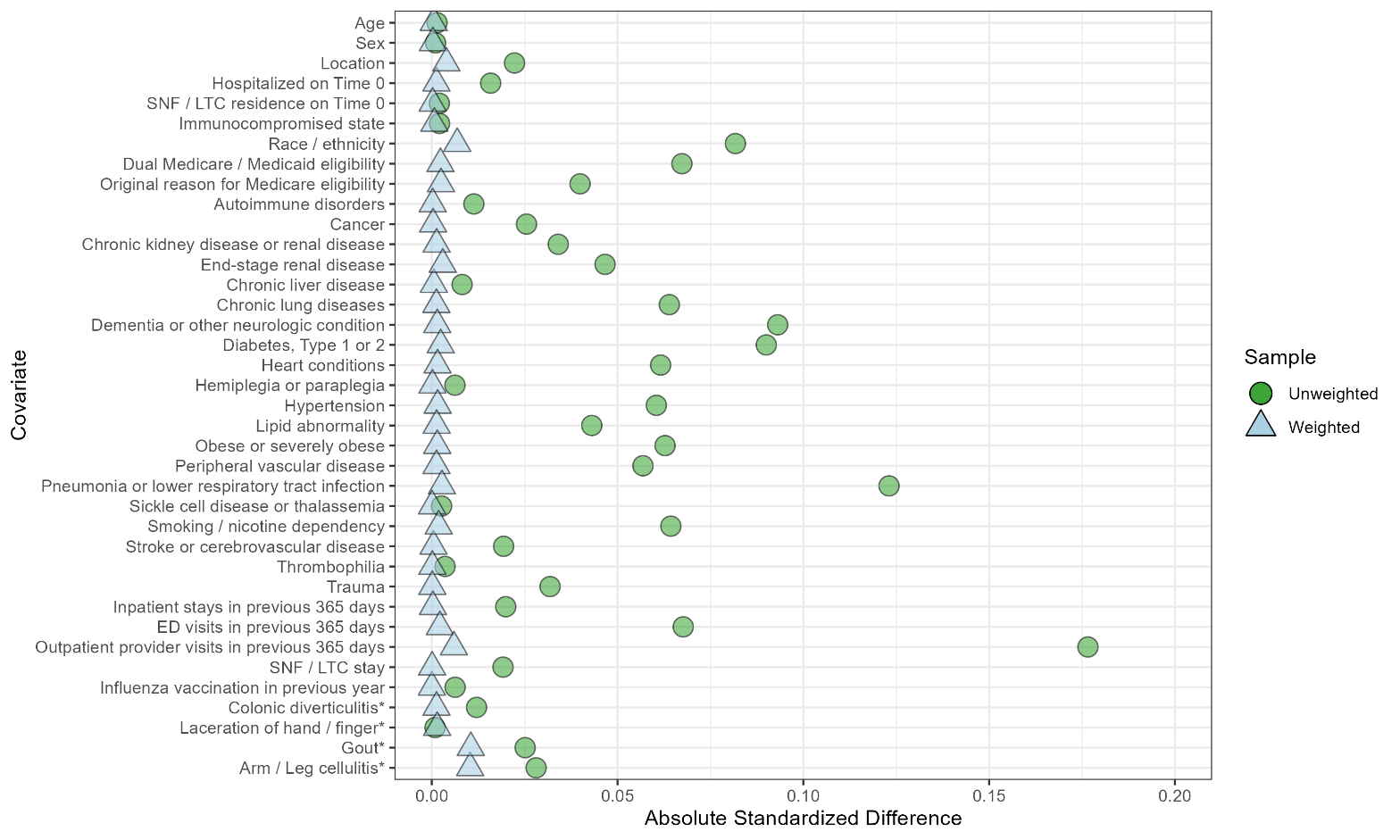

M. Hemorrhagic Stroke, Medicare

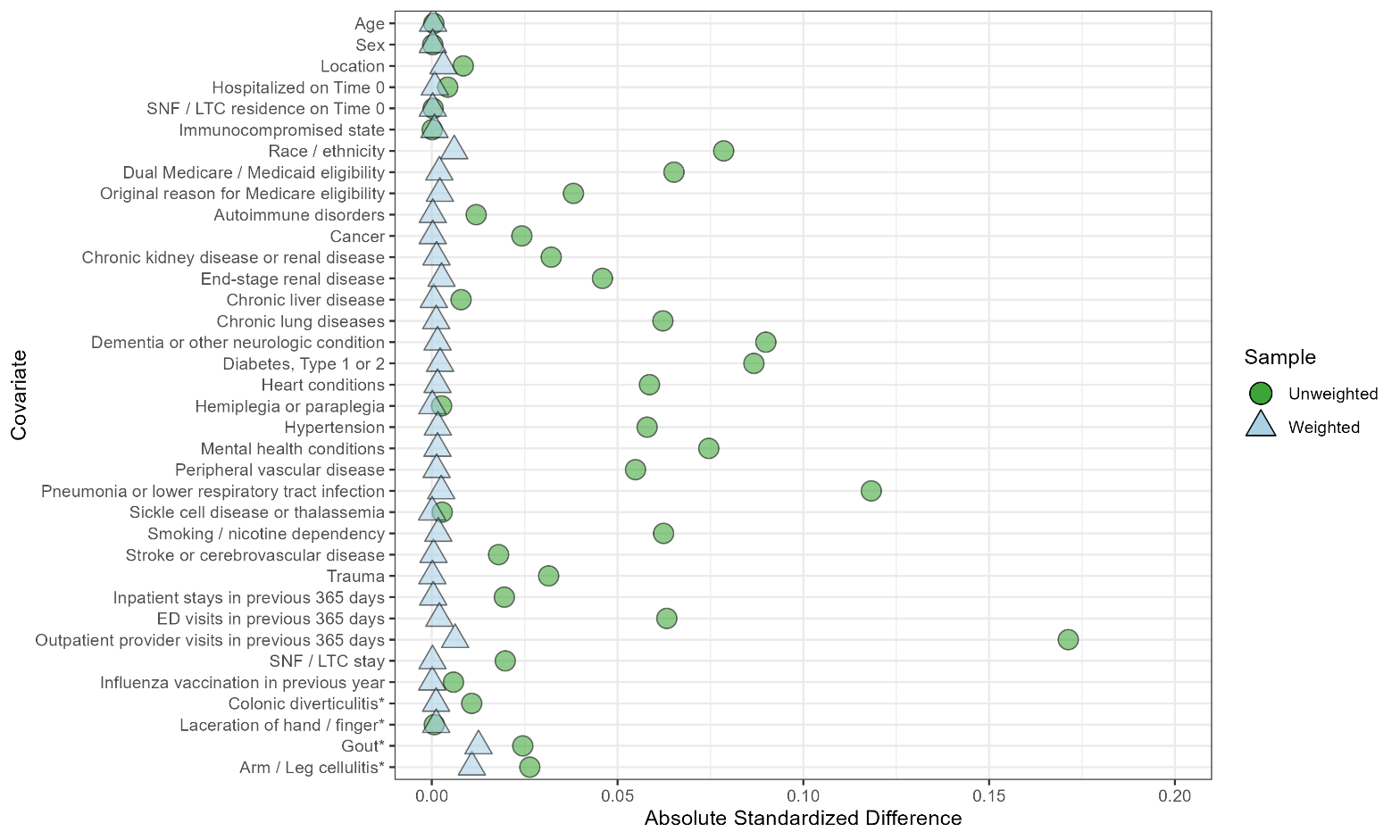

N. Acute Myocardial Infarction, Medicare

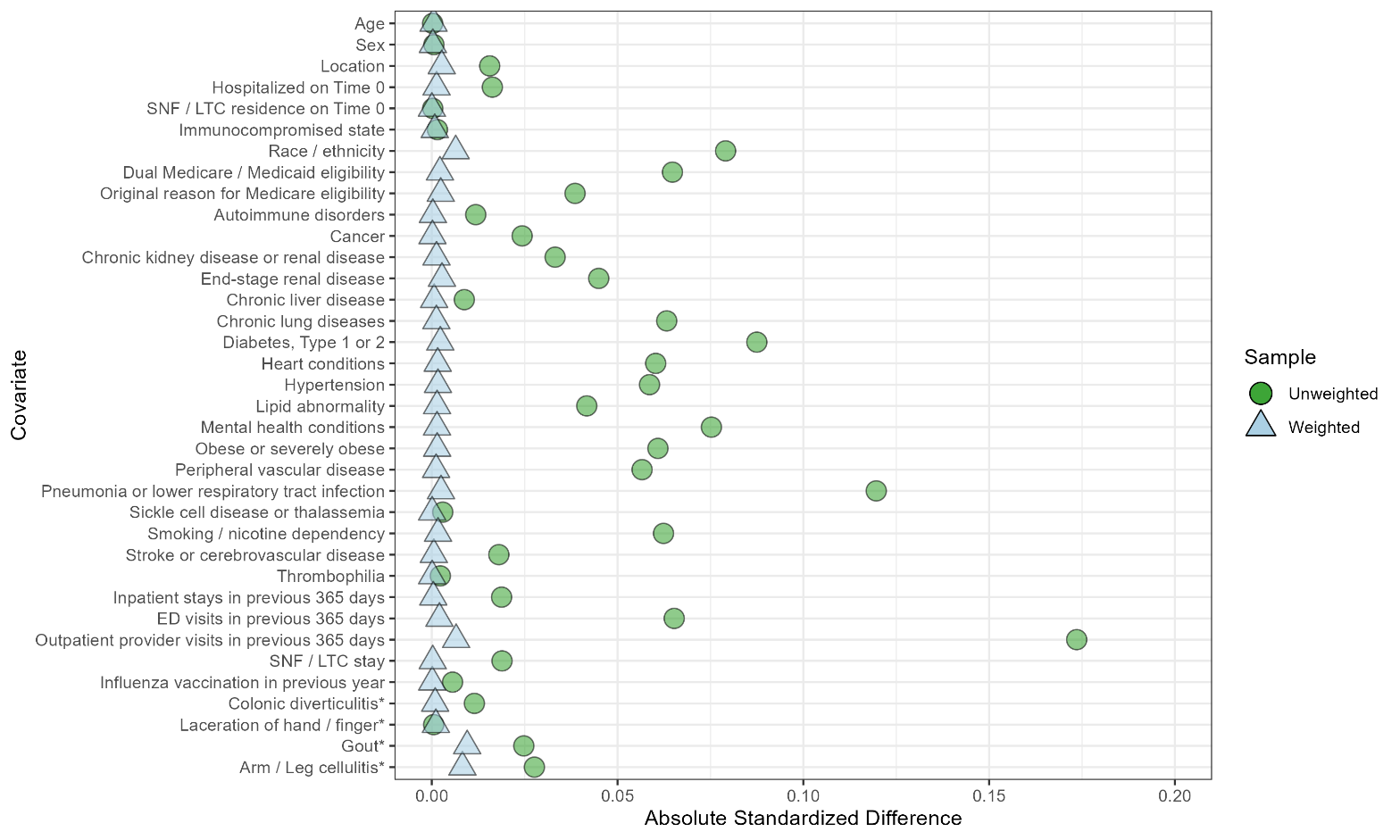

O. Myocarditis/Pericarditis, Medicare

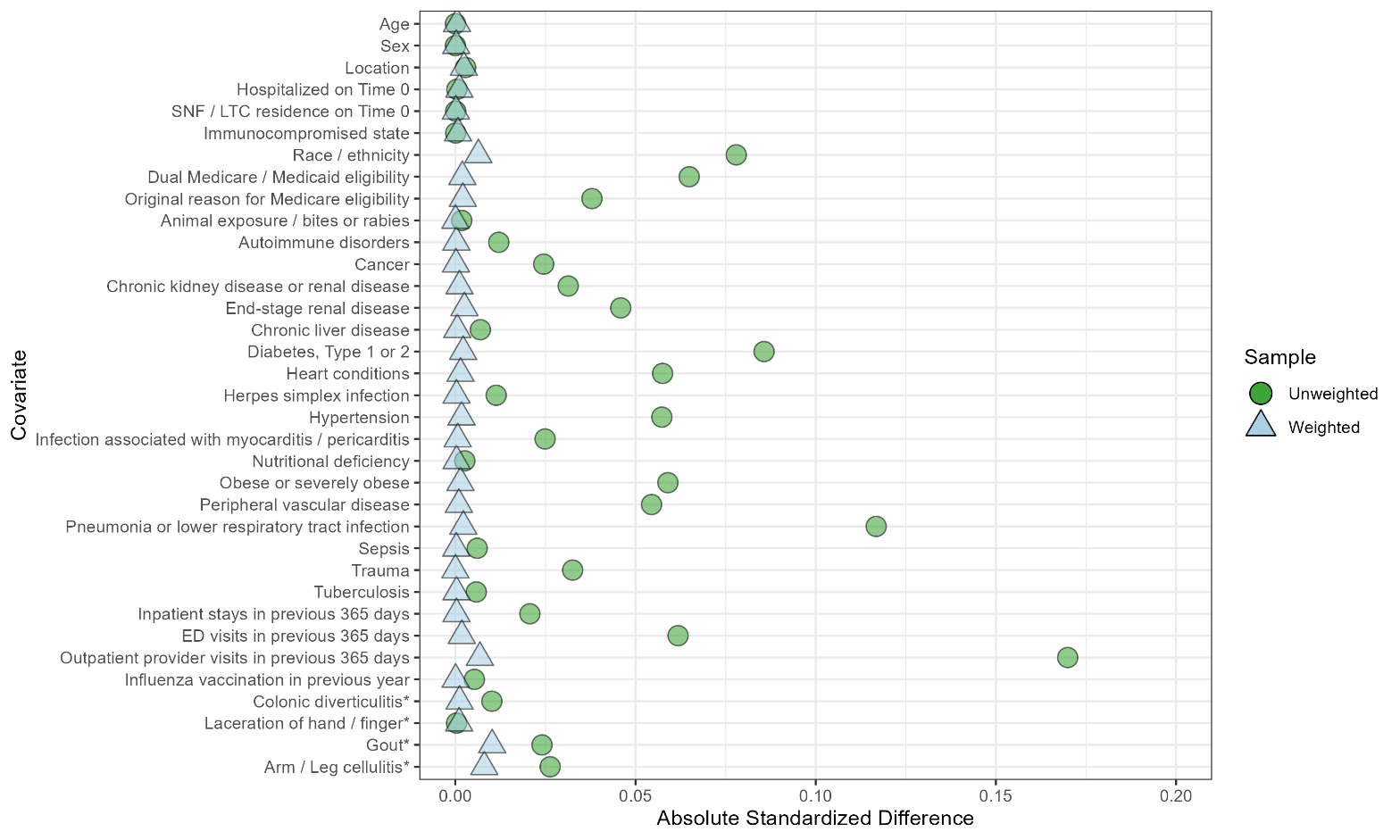

P. Deep Vein Thrombosis, Medicare

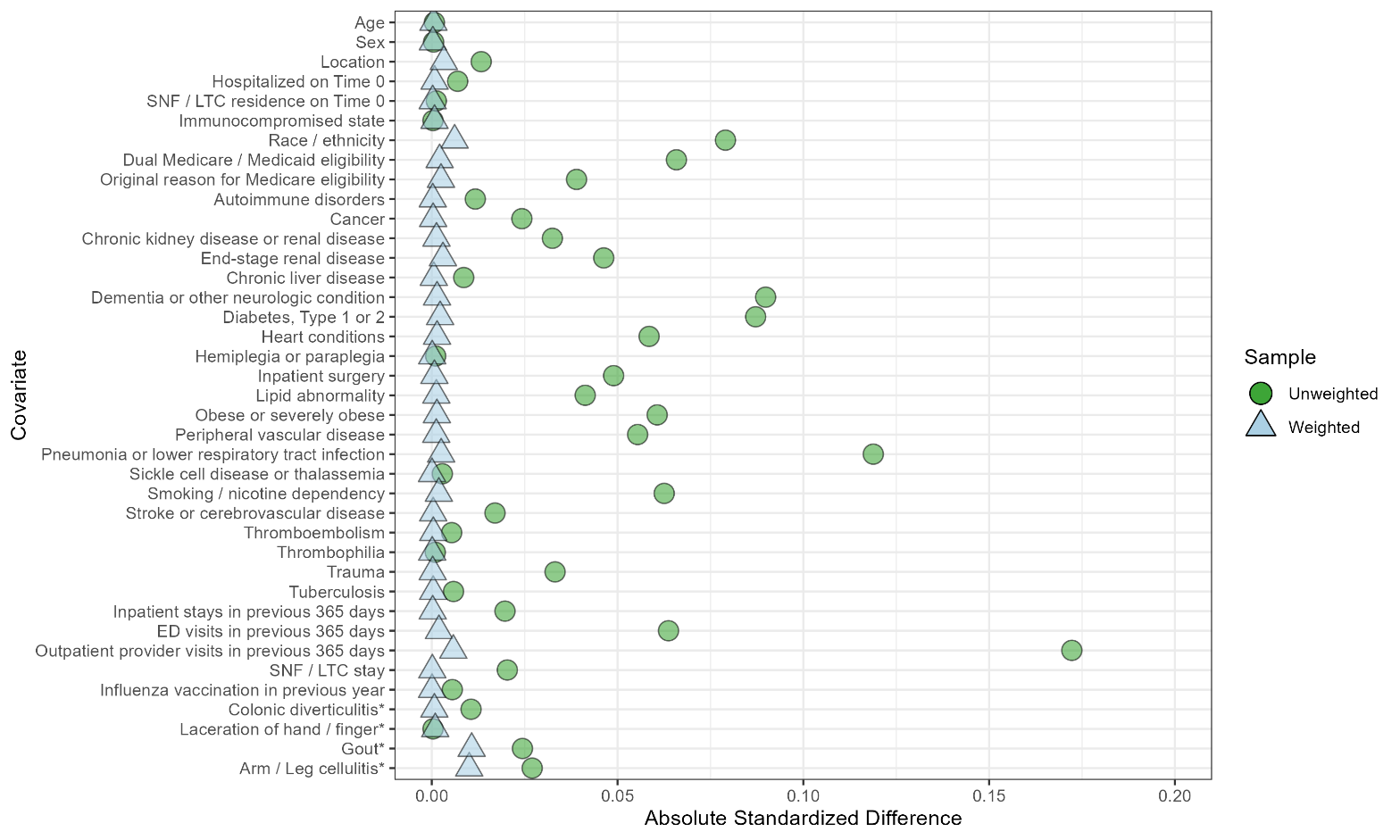

Q. Pulmonary Embolism, Medicare

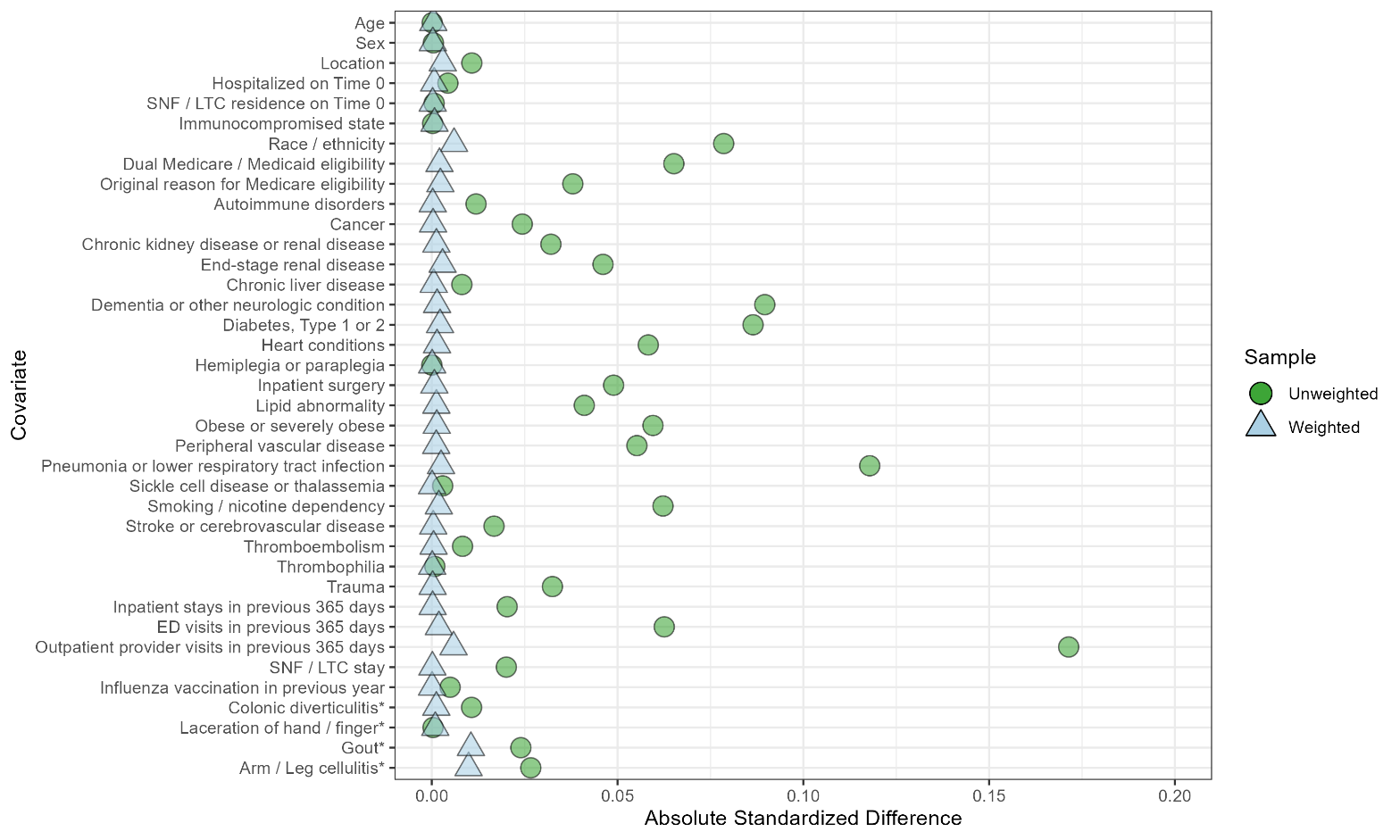

R. Disseminated Intravascular Coagulation, Medicare

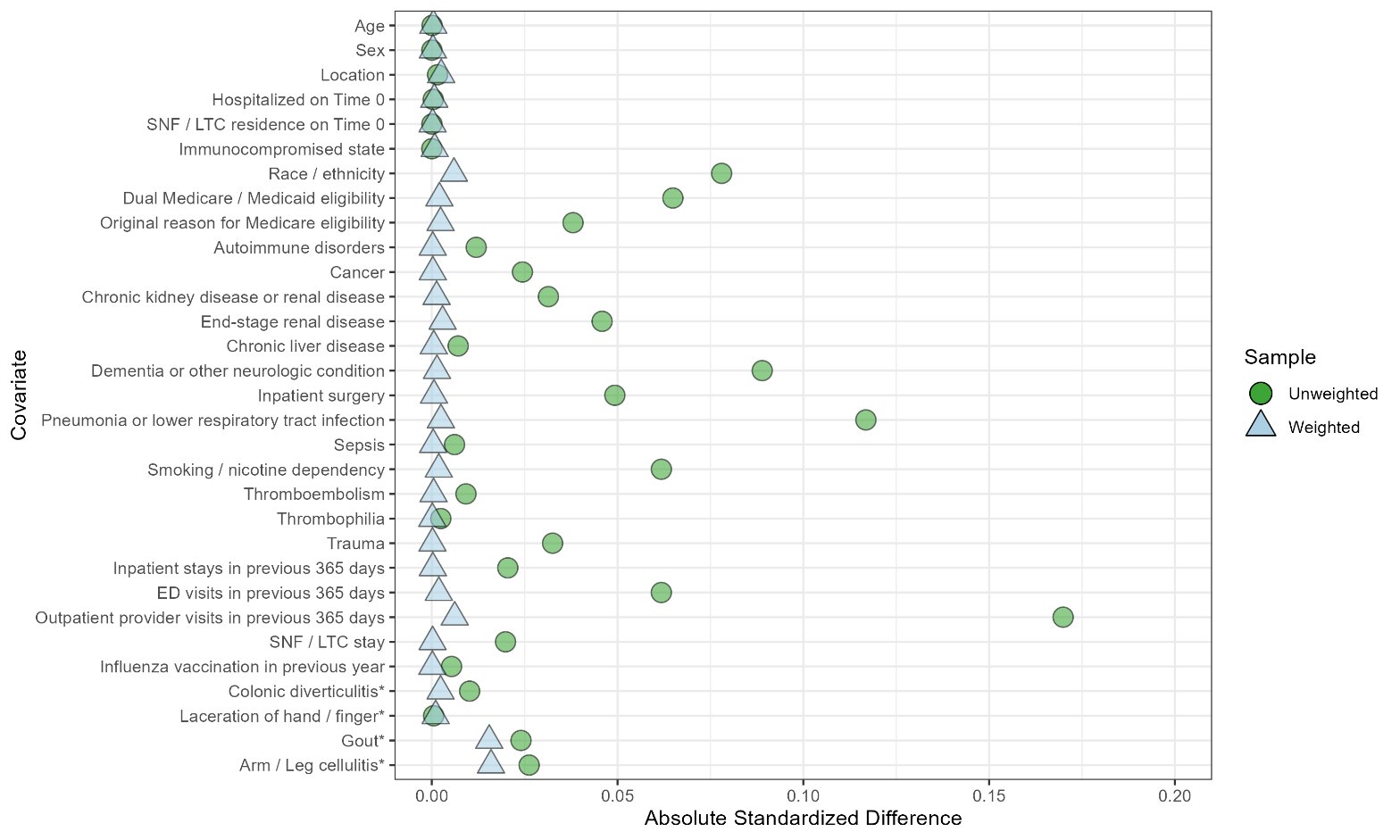

S. Unusual-Site Thrombosis With Thrombocytopenia Syndrome, Medicare

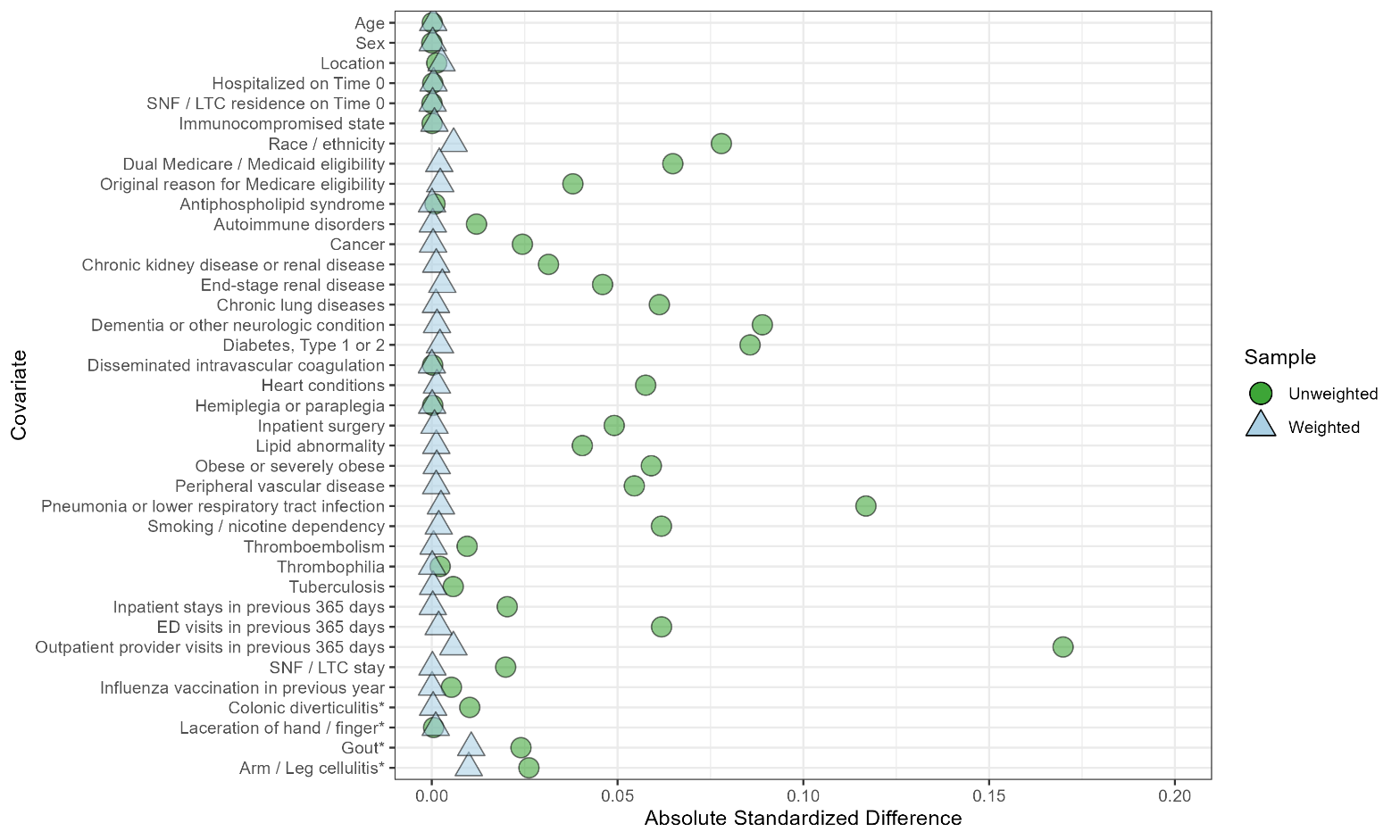

T. Common-Site Thrombosis With Thrombocytopenia Syndrome, Medicare

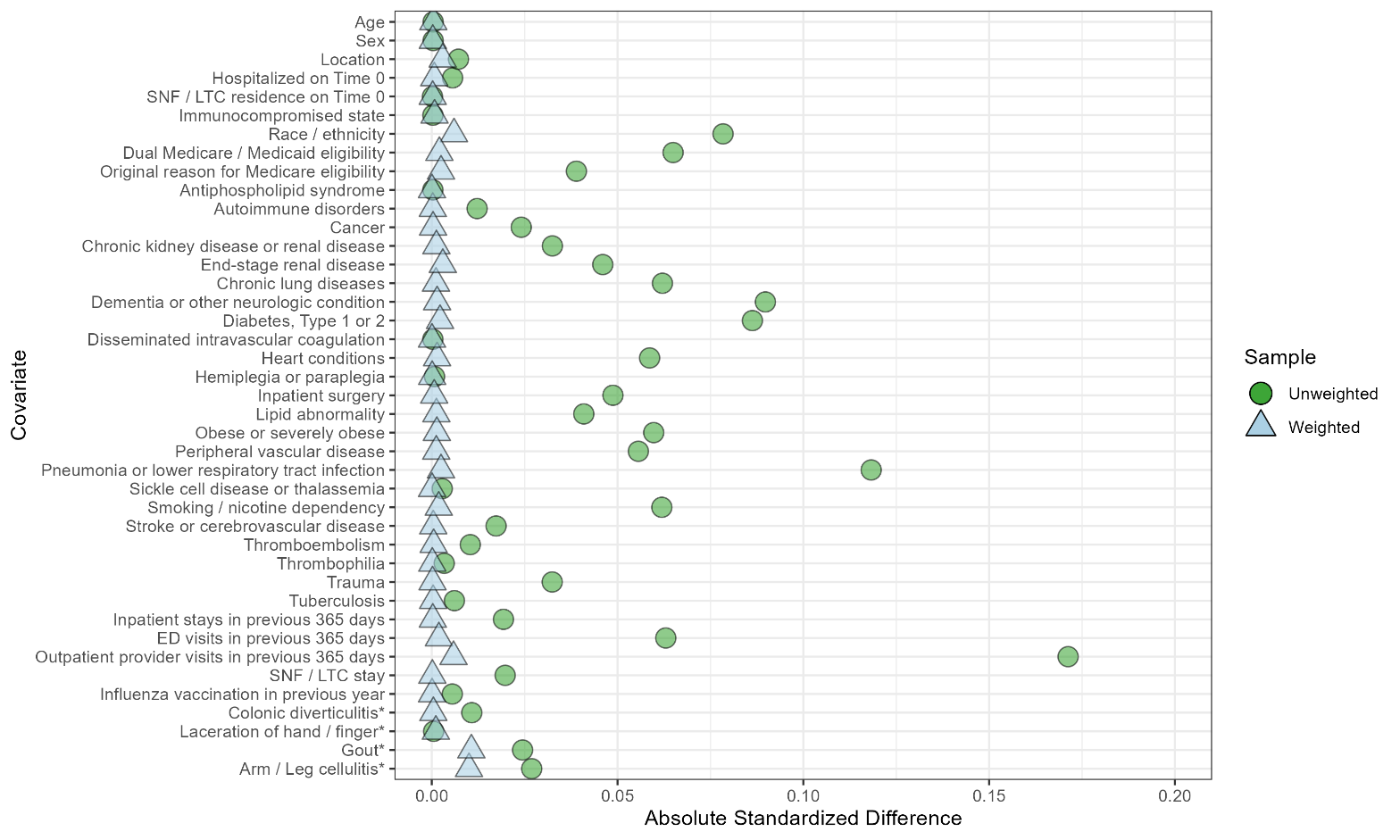

ASD = absolute standardized difference; COVID-19 = coronavirus disease 2019; ED = emergency department; LTC = long-term care; SNF = skilled nursing facility.

* Indicates the history of negative control conditions occurring before Time 0.
